# Trans-ancestry genome-wide analysis of atrial fibrillation provides new insights into disease biology and enables polygenic prediction of cardioembolic risk

**DOI:** 10.1101/2021.09.06.21263189

**Authors:** Kazuo Miyazawa, Kaoru Ito, Zhaonan Zou, Hiroshi Matsunaga, Satoshi Koyama, Hirotaka Ieki, Seitaro Nomura, Masato Akiyama, Ryo Kurosawa, Hiroki Yoshida, Kouichi Ozaki, Yoshihiro Onouchi, BioBank Japan Project, Atsushi Takahashi, Koichi Matsuda, Yoshinori Murakami, Hiroyuki Aburatani, Michiaki Kubo, Yukihide Momozawa, Chikashi Terao, Shinya Oki, Hiroshi Akazawa, Yoichiro Kamatani, Issei Komuro

**Affiliations:** Laboratory for Cardiovascular Genomics and Informatics, RIKEN Center for Integrative Medical Sciences, Yokohama, Japan; Department of Drug Discovery Medicine, Kyoto University Graduate School of Medicine, Kyoto, Japan; Department of Cardiovascular Medicine, Graduate School of Medicine, The University of Tokyo, Tokyo, Japan; Program in Medical and Population Genetics, Broad Institute of Harvard and MIT, Cambridge, MA, USA; Cardiovascular Research Center, Massachusetts General Hospital, Boston, MA, USA; Laboratory for Statistical and Translational Genetics, RIKEN Center for Integrative Medical Sciences, Kanagawa, Japan; Department of Ocular Pathology and Imaging Science, Kyushu University Graduate School of Medical Sciences, Fukuoka, Japan; Division for Genomic Medicine, Medical Genome Center, National Center for Geriatrics and Gerontology, Obu, Japan; Department of Public Health, Chiba University Graduate School of Medicine, Chiba, Japan; Institute of Medical Science, The University of Tokyo, Tokyo, Japan; Department of Genomic Medicine, Research Institute, National Cerebral and Cardiovascular Center, Suita, Japan; Department of Computational Biology and Medical Science, Graduate School of Frontier Sciences, The University of Tokyo, Tokyo, Japan; Division of Molecular Pathology, Institute of Medical Science, The University of Tokyo, Tokyo, Japan; Genome Science Division, Research Center for Advanced Science and Technology, The University of Tokyo, Tokyo, Japan; RIKEN Center for Integrative Medical Sciences, Kanagawa, Japan; Laboratory for Genotyping Development, RIKEN Center for Integrative Medical Sciences, Kanagawa, Japan

## Abstract

To understand the genetic underpinnings of atrial fibrillation (AF) in the Japanese population, we performed a large-scale genome-wide association study comprising 9,826 cases of AF among 150,272 individuals and identified five new susceptibility loci, including East Asian-specific rare variants. A trans-ancestry meta-analysis of >1 million individuals, including 77,690 cases, identified 35 novel loci. Leveraging gene expression and epigenomic datasets to prioritize putative causal genes and their transcription factors revealed the involvement of *IL6R* gene and transcription factor ERG besides the known ones. Further, we constructed a polygenic risk score (PRS) for AF, using the trans-ancestry meta-analysis. PRS was associated with an increased risk of long-term cardiovascular and stroke mortality, and segregated individuals with cardioembolic stroke in undiagnosed AF patients. Our results provide novel biological and clinical insights into AF genetics and suggest their potential for clinical applications.

---

Atrial fibrillation (AF) is the most common cardiac arrhythmia, affecting approximately 46.3 million individuals worldwide^1^. The global prevalence of AF is increasing due to the rapid aging of the general population and intensified search for subclinical AF^2^. Despite progress in diagnostic and therapeutic technologies, a substantial number of patients with AF are admitted with life-threatening complications such as stroke and heart failure^3^, causing a considerable burden on patients and public healthcare systems^4^. Besides conventional clinical risk factors such as ageing, obesity, hypertension, and heart failure, the genetic contribution to the development of AF is also widely recognized. Recent genome-wide association studies (GWASs) have identified more than 100 AF-associated loci, some of which are involved in cardiac developmental, electrophysiological, contractile, and structural pathways^5–8^. However, since the vast majority of AF-GWASs have been predominantly performed in European populations, the genetic pathophysiology of AF in non-European populations has not been comprehensively understood and it is difficult to apply polygenic risk scores (PRSs) derived from such GWASs to non-European populations.

Therefore, in the present study, we sought to explore the genetic architecture of AF in a non-European population and improve the statistical power of AF-GWASs by performing a large-scale Japanese GWAS, followed by conducting a trans-ancestry meta-analysis. Further, we investigated the biological role of the identified AF-associated loci by leveraging gene expression and epigenomic datasets. Additionally, we developed a PRS derived from the trans-ancestry meta-analysis and assessed the impact of the PRS on relevant phenotypes and long-term mortality, which may provide evidence for the clinical utility of AF-PRS and lay the foundation for the realization of precision medicine in AF.

## Results

### Five novel loci for AF identified in the Japanese GWAS

An overview of the study design is shown in Supplementary Fig. 1. We performed a GWAS on the case-control dataset from the BioBank Japan (BBJ) that comprised 9,826 AF cases and 140,446 controls, using 16,394,105 variants in the autosomes and 423,039 variants in the X chromosome with a minor allele frequency (MAF) > 0.1%. The GWAS identified 31 AF-associated loci with genome-wide significance, of which five were previously unreported (Table 1, Supplementary Table 1, Supplementary Fig. 2, and Supplementary Datasets 1 and 2). The proportion of the variation in AF (the single nucleotide polymorphism (SNP) heritability; *h*^2^) explained by the total genome-wide genetic variation detected in the current Japanese GWAS was estimated to be 6.1% (standard error of mean [s.e.m.] 1.4%), and the liability-scale *h*^2^ was estimated at 11.7% (s.e.m. 2.6%) using linkage disequilibrium (LD)-score regression.

**Table 1.** Novel association loci in the Japanese genome-wide association study (GWAS). Sentinel variants in novel loci with a genome-wide significance in the Japanese GWAS (9,826 cases and 140,446 controls). CHR, chromosome; rsID, reference SNP cluster ID; POS, position (hg19); REF, reference allele; ALT, alternate allele; BBJ, BioBank Japan; AAF, alternate allele frequency; EUR, European populations; SE, standard error.

| CHR | POS (hg19) | rsID | REF | ALT | AAF |  |  | Beta | SE | P | Gene | Annotation |
| --- | --- | --- | --- | --- | --- | --- | --- | --- | --- | --- | --- | --- |
|  |  |  |  |  | BBJ | gnomAD EUR<br>(non-Finnish) | genomAD EUR<br>(Finnish) |  |  |  |  |  |
| 6 | 152466619 | rs202030113 | T | C | 0.012 | 0 | 0 | 0.352 | 0.063 | 1.90E-08 | *SYNE1* | Intronic |
| 12 | 104471663 | rs2930856 | C | T | 0.579 | 0.861 | 0.854 | 0.090 | 0.015 | 5.80E-09 | *HCFC2* | Intronic |
| 16 | 30619745 | rs1055894680 | C | T | 0.001 | <0.001 | 0.001 | 1.215 | 0.158 | 1.58E-14 | *ZNF689* | Intronic |
| X | 23399501 | rs73205368 | T | C | 0.284 | 0.045 | 0.015 | 0.089 | 0.012 | 7.50E-13 | *PTCHD1* | Intronic |
| X | 137790580 | rs778479352 | T | C | 0.002 | 0 | 0 | 0.692 | 0.075 | 1.62E-20 | *FGF13* | Intronic |

We found two novel loci whose lead variants were observed only in East Asian according to the Genome Aggregation Database v2.1.1 (gnomAD)^9^: one low-frequency and one rare lead variant, rs202030113 (MAF = 1.2%) and rs778479352 (MAF = 0.25%), respectively. The lead variant on 6q25.1, rs202030113, was located in the intronic region three bases away from an exon-intron boundary of *SYNE1* and was predicted as a splice donor loss with the spliceAI delta score of 0.33^10^. *SYNE1* encodes nesprin-1 (spectrin repeat) protein, and together with Sad1p/UNC84-domain-containing proteins (SUN1/2), compose the nuclear envelope protein complex via its nucleoplasmic domains to lamin A/C. Mutations in *LMNA* and *SYNE1* genes have been identified in patients with severe muscle dystrophy and dilated cardiomyopathy^11, 12^. Mutations in the *SYNE1* gene cause defects in nuclear morphology, myoblast differentiation, and heart development^13^, altering the nuclear envelope protein complex that contributes to the structural substrate in atrial arrhythmogenesis. A strong signal (odds ratio [OR] for AF development = 2.00, 95% confidence interval [CI] = 1.73-2.31, *P* = 1.6 × 10^-20^) was displayed by rs778479352, the lead variant located in an intron of *FGF13*. *FGF13* encodes a member of the fibroblast growth factor family, which possesses broad mitogenic and cell survival activities. *FGF13* directly binds to the C-terminus of the main cardiac sodium channel (Na_V_1.5) in the sarcolemma; knockdown of *FGF13* in rat cardiomyocytes showed a loss of function of Na_V_1.5-reduced Na^+^ current density, decreased Na^+^ channel availability, and slowed Na_V_1.5-reduced Na^+^ current recovery from inactivation^14^. This evidence of conduction disturbance in cardiomyocytes indicates that *FGF13* is an important target gene associated with AF.

To identify AF-associated variants independent of the lead variant at each locus, we performed a stepwise conditional analysis, in which 18 independent variants (locus-wide *P* < 5.0×10^-6^) were additionally detected, increasing the total number of AF-associated signals to 49 (Supplementary Fig. 3 and Supplementary Table 2). We identified 10 loci that had multiple independent association signals, especially in the *PITX2-C4orf32* locus with 6 association signals (*N*_signal_ = 2: *GORAB-PRRX1*, *CAND2*, *HAND2-AS1*, *FANCC*, *NEBL*, *AKAP6*, *ZFHX3* loci, *N*_signal_ = 3: *LINC02459-TBX5* locus, *N*_signal_ = 4: *NEURL1* locus, and *N*_signal_ = 6: *PITX2-C4orf32* locus). Of these additional signals, three variants were observed only in East Asian populations in gnomAD (rs577463446 at the *FANCC* locus, MAF = 0.5%, OR = 1.58; rs965277670 at *NEBL* locus, MAF = 0.6%, OR = 1.65; rs201901902 at *NEURL1* locus MAF = 0.5%, OR = 1.68).

### Sex-stratified GWAS for AF

Sex-related differences in the epidemiology, clinical presentation, and prognosis of AF have been consistently reported^15, 16^. For example, the age-stratified prevalence of AF is lower in female than in male, despite a higher risk for AF-related outcomes such as stroke in female^17–19^. Thus, we first perfomed sex-stratified GWAS and calculated the genetic correlation between sexes using LD-score regression to explore the genetic heterogeneity. We then found a significant correlation (*r*_g_ = 1.095, s.e.m. = 0.112, *P* = 1.2 × 10^−22^). Next, we examined genetic loci with sex-dependent effects on AF development in the sex-stratified GWAS. We found 21 loci with genome-wide significance in the male GWAS, but only six loci in the female GWAS (Supplementary Fig. 4a and Supplementary Table 3). Furthermore, by comparing the effect sizes of the lead variants identified in the sex-stratified GWAS, we observed significant positive correlation and concordant allelic effects between sexes (Spearman’s ρ = 0.92, *P* = 7.4 × 10^-7^; Supplementary Fig. 4b). We found several variants with significant heterogeneity in the *PITX2-C4orf32* and *CUX2* loci (*P*_het_ < 1.0 × 10^-4^). Although the *PITX2-C4orf32* locus reached genome-wide significance in both sex-stratified GWAS (β_male_ (s.e.m._male_) = 0.569 (0.018), *P*_male_ = 1.9 × 10^−210^; β_female_ (s.e.m._female_) = 0.381 (0.027), *P*_female_ = 3.0 × 10^−46^, for rs12644625, the lead variant in the *PITX2-C4orf32* locus), the *CUX2* locus in the female GWAS did not achieve genome-wide significance level (β_male_ (s.e.m._male_) = −0.280 (0.024), *P*_male_ = 9.7 × 10^−33^; β_female_ (s.e.m._female_) = −0.105 (0.034), *P*_female_ = 2.1 × 10^−3^, for rs12644625, the lead variant in the *CUX2* locus). *CUX2* is a transcription factor regulating sex-biased gene expression in rat and mouse liver^20, 21^, and the variant in the *CUX2* locus is associated with a DNA methylation site near the *ALDH2* gene, encoding aldehyde dehydrogenase, which catalyzes the oxidation of aldehydes into carboxylic acids^22^. Moreover, according to Genotype-Tissue Expression (GTEx) data^23^, *CUX2* gene expression in liver was significantly different between male and female (*P* = 7.8 × 10^−4^, Wilcoxon rank-sum test; Supplementary Fig. 5). These results suggest that sex-biased gene expression related to the metabolism of aldehydes in the liver may partially explain the sex-related differences in the genetic basis of AF.

### Trans-ancestry meta-analysis identified 33 novel loci for AF

To improve the statistical power to detect further genetic associations with AF, we conducted a trans-ancestry meta-analysis by combining the current Japanese GWAS (BBJ) and two European GWASs: a large-scale meta-analysis of European populations (EUR)^7^ and a biobank data of FinnGen data release 2 (FIN). All three datasets yielded 77,690 cases (BBJ: 9,826; EUR: 60,620; FIN: 7,244) and 1,167,040 controls (BBJ: 140,446; EUR: 970,216; FIN: 56,378). We tested a total of 5,158,449 variants with MAF ≥ 1% and identified 150 AF-associated loci with genome-wide significance (log_10_ Bayes factor (BF) > 6; Fig. 1, Supplementary Table 4, and Supplementary Datasets 3 and 4). Of these loci, 33 have not been reported previously, including three novel loci detected in the current Japanese GWAS. In total, we identified 35 novel loci through the current Japanese GWAS and trans-ancestry meta-analysis (Table 2).

**Fig. 1.**
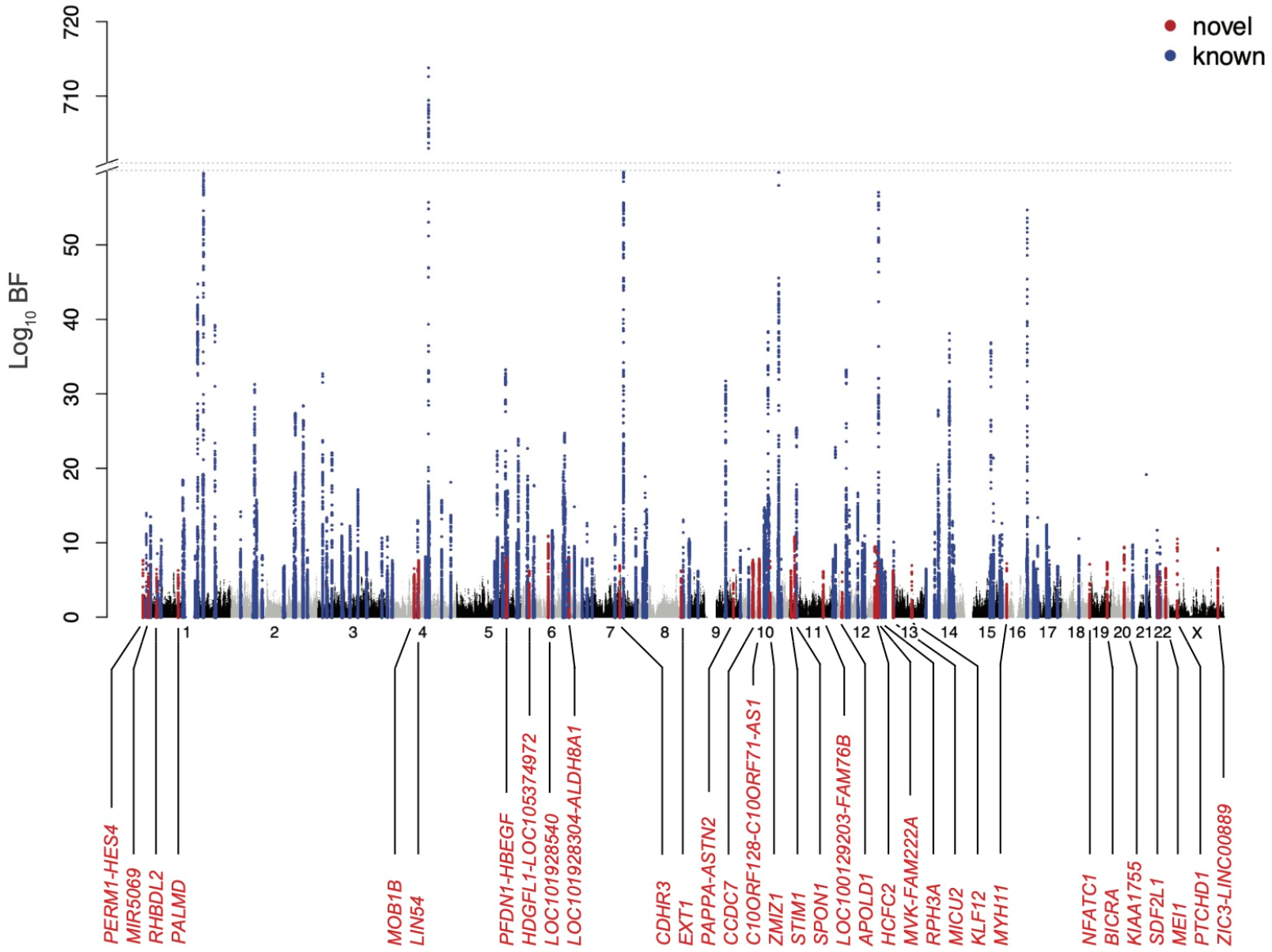
Manhattan plot for the trans-ancestry meta-analysis. The results of the trans-ancestry meta-analysis (77,690 AF cases and 1,167,040 controls) are shown. The log_10_ BFs on the *y* axis are shown against the genomic positions (hg19) on the *x* axis. Association signals that reached a genome-wide significance level (log_10_ BF > 6) are shown in blue if previously reported loci and in red if novel loci. AF, atrial fibrillation; BF, Bayes factor.

**Table 2.**
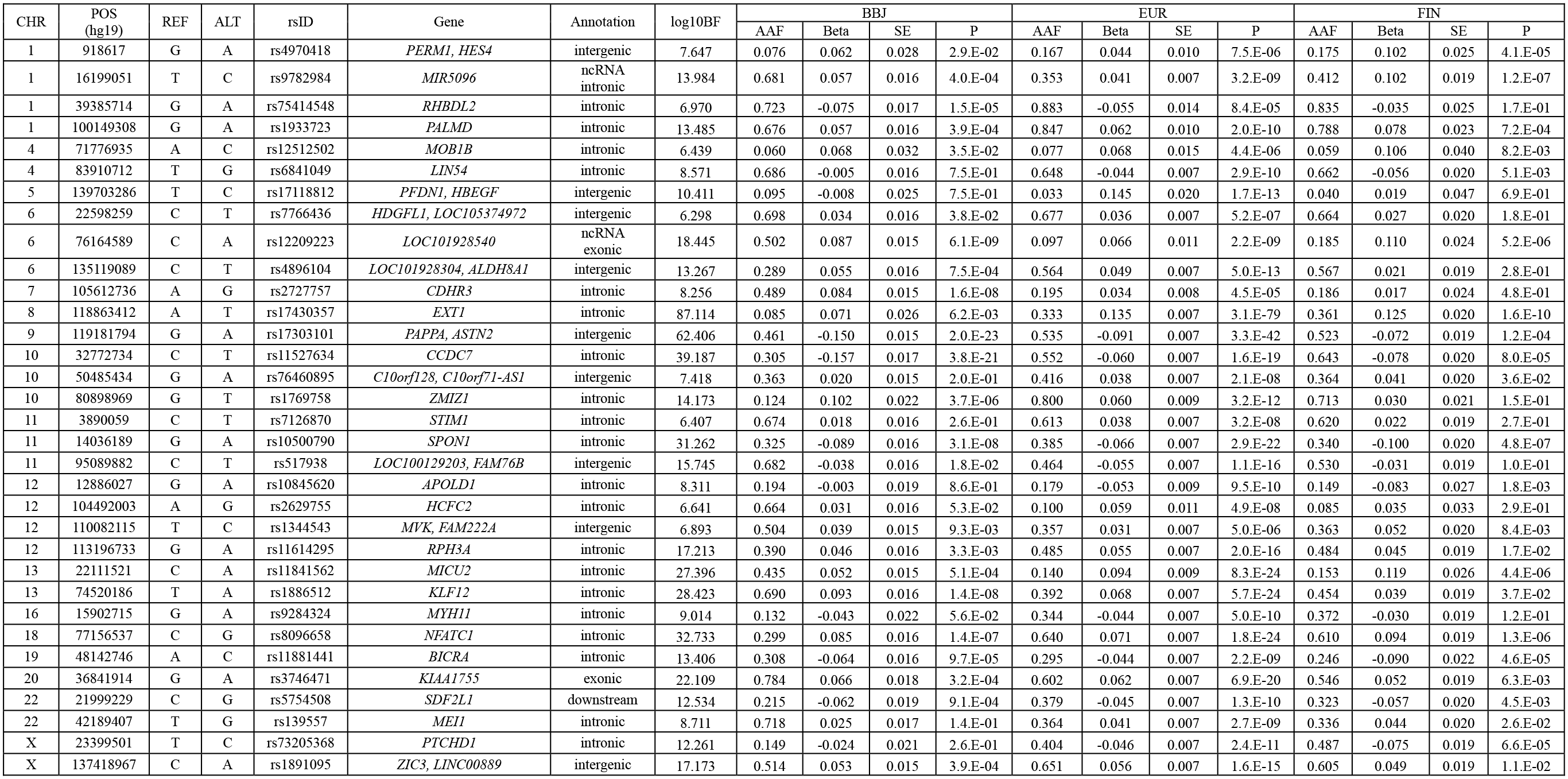
Novel association loci in the trans-ancestry meta-analysis. Sentinel variants in novel loci with a genome-wide significance in the Japanese GWAS (77,690 cases and 1,167,040 controls). CHR, chromosome; POS, position (hg19); REF, reference allele; ALT, alternate allele; rsID, reference SNP cluster ID; BF, Bayes factor; AAF, alternate allele frequency; BBJ, BioBank Japan; SE, standard error; ncRNA, non-coding RNA.

Of the 5,855 variants in LD (*r*^2^ > 0.6) with 150 lead variants, 27 missense variants were observed (Supplementary Table 5). Among novel loci, we found a missense variant, rs848208 (p.Ala970Val), in the *SPEN* gene, encoding a hormone-inducible transcriptional co-regulator that activates and represses downstream targets. *SPEN*-deficient zebrafish embryos developed bradycardia, atrioventricular block, and heart chamber fibrillation with downregulation of connexin 43 expression^24^, which is a well-known component of gap junctions and is associated with the cardiac conduction system^25^. Another missense variant in novel loci, rs3746471 (p.Arg1045Trp), was located on the *KIAA1755* gene, which is reported to be associated with heart rate^26^ and heart rate variability^27^. Given the robust relationship between autonomic nervous dysfunction and AF^28^, *KIAA1755* can be a potential target gene for neural modulation contributing to AF management; however, the biological association between the *KIAA1755* gene and AF has not been fully examined.

### Shared allelic effects between Japanese and European populations and fine mapping/credible set analyses in the trans-ancestry meta-analysis

We compared alternate allele frequencies and allelic effects for 150 lead variants between BBJ, EUR, and FIN. Compared to substantial concordance in allele frequencies between EUR and FIN (Spearman’s ρ = 0.974, *P* < 2.2 × 10^−16^), we observed a moderate correlation in allele frequencies between BBJ and EUR as well as BBJ and FIN (ρ = 0.592, *P* = 1.5 × 10^−15^ and ρ = 0.632, *P* < 2.2 × 10^−16^, respectively; Supplementary Fig. 6a-c). Additionally, we found a significant positive correlation between concordant allelic effects of these variants (ρ = 0.769, *P* < 2.2 × 10^−16^ for BBJ vs. EUR; ρ = 0.769, *P* < 2.2 × 10^−16^ for BBJ vs. FIN; Supplementary Fig. 6d-f). To further explore the relationship of allelic effects between populations, we performed a trans-ancestry genetic correlation analysis, which showed strong correlation of BBJ with EUR and FIN (BBJ and EUR: *r*_g_ = 0.990, s.e.m. = 0.097, BBJ and FIN *r*_g_ = 0.955, s.e.m. = 0.344).

To assess the contribution of trans-ancestry meta-analyses to the refinement of the putative causal variants, we constructed 99% credible sets for the 150 AF-associated loci detected by the current trans-ancestry meta-analysis, and compared the number of variants included in the 99% credible sets derived from three combinations of the meta-analysis (EUR + BBJ, EUR + FIN, BBJ + EUR + FIN; Supplementary Fig. 6g, h). The size of the 99% credible sets derived from EUR + BBJ significantly decreased compared with those from EUR + FIN (*P* = 0.004, paired Wilcoxon rank-sum test). In addition, trans-ancestry meta-analysis of three datasets (BBJ + EUR + FIN) yielded the most significant decrease in the number of variants among all the combinations of meta-analyses (median number of variants = 12; interquartile range [IQR] 5 – 36; Supplementary Table 6). In particular, a single variant, rs67329386, was identified on the *ZFHX3* locus with a high posterior probability of 0.995, only in the trans-ancestry meta-analysis of three datasets. In previous AF-GWAS, several variants were found in the *ZFHX3* locus^29, 30^, where a large transcription factor, *ZFHX3*, together with *PITX2*, facilitated DNA binding and transcription activity^31^. To corroborate the role of rs67329386 as a causal variant, we searched for transcription factors binding to this locus using the ChIP-seq dataset in ChIP-Atlas^32^. Compared to other AF-associated variants within this locus, rs67329386 was distinctly located on the binding site of CEBPB (Supplementary Fig. 7), aiding in the recognition and binding of target gene regulatory regions, leading to cell proliferation, differentiation, immune response, and tumor formation^33^.

### Pleiotropic effects and functional pathways of AF-associated loci

To characterize the AF-associated loci, we examined the pleiotropic effects of the identified lead variants and proxies using the NHGRI-EBI GWAS catalog database. Of the 150 AF-associated loci, 114 (76.0%) had at least one overlapping variant, and 739 variants overlapped with 200 diseases or traits (Supplementary Table 7). Among them, blood pressure-related traits were most frequently observed (13.0%), followed by traits related to electrocardiogram (10.1%) and heart rate (4.0%).

Given the heterogeneous study design and population in the GWAS catalog, we subsequently utilized the BBJ dataset^34–36^ with a consistent study design and population, to investigate the pleotropic effects. In particular, to explore the biological pathways related to AF development, we searched for AF-associated variants overlapping with quantitative trait loci (QTL) for clinical measurements such as anthropometric, metabolic, kidney-related, and blood pressure data (Supplementary Table 8). We found 160 loci-phenotype association pairs, where distinct functional clusters were observed (Supplementary Fig. 8); positive impact on blood pressure: *CASZ1*, *PRDM8-FGF5*, *CUX2*, and *RPH3A*, and negative impact on renal function: *MIR5096*, *MBD5*, *DNAJC12-SIRT1*, *IGF1R*, and *NFATC1*. Moreover, we observed associations on chromosome 6p21.31 harboring *HMGA1*, in which AF-associated variants overlapped with QTLs for height (rs1759628) and body mass index (BMI) (rs6913361). Both rs1759628 and rs6913361 were located 7 kilobases (kb) and 25 kb upstream of *HMGA1*, respectively, and the risk alleles of these variants were significantly associated with increased height and BMI (β_height_ (s.e.m._height_) = 0.090 (0.005), *P*_height_ = 8.7 × 10^−93^; β_BMI_ (s.e.m._BMI_) = 0.037 (0.005), *P*_BMI_ = 1.7 × 10^−12^, respectively). *HMGA1* encoded a chromatin-associated protein that attach to AT-rich regions of DNA and recruit transcription factors, resulting in upregulation of the insulin receptor gene expression^37^ and the variants in *HMGA1* were associated with increased risk of metabolic syndrome^38^.

To enhance the biological understanding of the AF-associated loci, we performed in silico tissue and gene-set enrichment analysis using DEPICT software^39^, which prioritizes candidate genes and tissues based on gene expression data from different tissues and cell types. Consistent with a previous study^7^, we found significant enrichment of AF-associated loci predominantly in heart tissues (*P* < 1.2 × 10^−5^ for heart and *P* < 2.3 × 10^−5^ for heart atria; Supplementary Fig. 9 and Supplementary Table 9). Additionally, pathway analysis demonstrated that AF-associated loci were significantly enriched in 34 out of 1,157 gene sets, most of which were involved in heart development and morphogenesis (Supplementary Table 10). Among them, the regulation of cell adhesion was a previously unreported pathway (*P* = 1.6 × 10^−10^), in which *ZMIZ1*, the nearest gene for rs1769758, was reported to regulate the activity of various transcription factors related to vascular development, androgen receptor coregulation, SMAD3 regulation, and coactivation of p53^40–42^. Knockdown of *ZMIZ1* in mice resulted in severe defects in the reorganization of the yolk sac vascular plexus and cell proliferation^43^.

### Prioritization of AF-associated genes and transcription factors using gene expression and epigenetic features

We performed a transcriptome-wide association study (TWAS) using the identified loci in the trans-ancestry meta-analysis and GTEx data^23^ to identify candidate genes associated with AF. Given the enrichment of AF-associated loci in the heart tissue, we used the gene expression data of GTEx in the atrial appendage and left ventricle as a reference. TWAS prioritized 86 and 74 candidate causal genes significantly associated with AF in the atrial appendage and left ventricle, respectively (Fig. 2a and Supplementary Table 11). We found only 14 and 15 overlapping genes between nearby genes located on the AF-associated loci and the candidate genes in the atrial appendage and left ventricle, respectively (Fig. 2b). Moreover, the median physical distances from the lead variants to the candidate genes were greater than 100 kb. Notably, 17.4% and 25.6% of the candidate genes were located 500 kb away from the lead variants (Fig. 2c). Further, to test whether the candidate genes were enriched in unique pathways, we used ingenuity pathway analysis and found several canonical pathways, including fatty acid metabolism-related pathways, in which the candidate genes prioritized in both atrial appendage and left ventricle were enriched (Fig. 2d). Intriguingly, *IL6R* was identified as a candidate gene associated with AF in the atrial appendage and was also involved in the canonical associated pathway. Despite increasing evidence for the role of inflammation in the AF pathophysiology^44^, including a report on the suggestive association of the *IL6R* locus with AF (*P* = 5.0 × 10^−4^)^45^, the genetic contribution of inflammatory process to AF development has not been fully elucidated. Our transcriptome-wide analysis revealed a significant association between *IL6R* and AF development, providing supportive evidence for the contribution of inflammation through *IL6R* to the pathophysiological mechanism underlying AF.

**Fig. 2.**
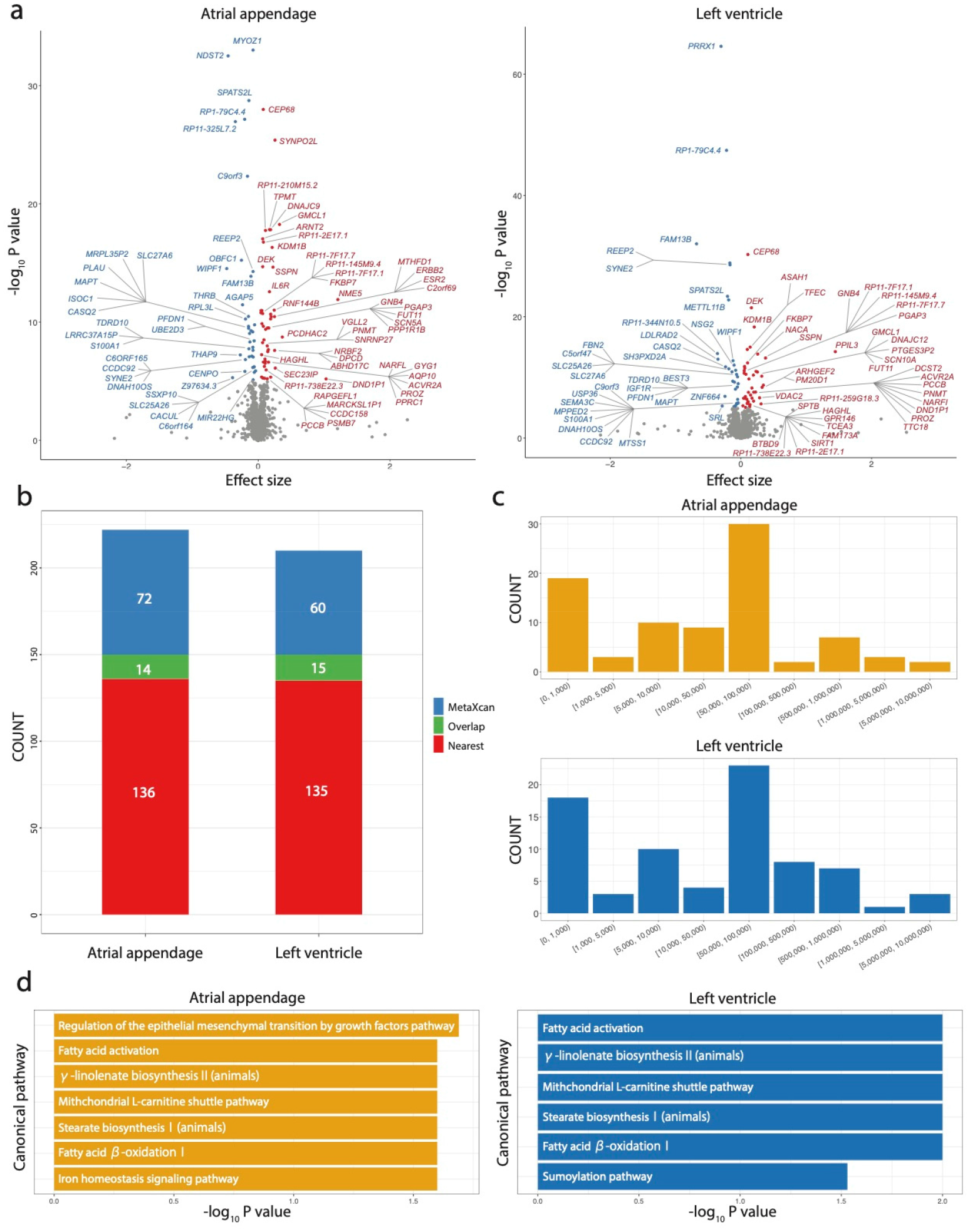
Transcriptome-wide association analysis. **a**, Volcano plot showing individual genes with the effect size and the -log_10_ *P* value for TWAS based on atrial appendage and left ventricular tissues from GTEx project. Significant genes adjusted Bonferroni correction for all tested genes in each tissue (*P* value < 0.05/5,988 for atrial appendage and < 0.05/5,378 for left ventricle) are highlighted with red (positive effect of predicted gene expression on AF) and blue (negative effect of predicted gene expression on AF). **b**, Number of genes located close to the lead variants (red), identified by TWAS, (blue) and overlapped between them (green) in atrial appendage and left ventricular tissues. **c,** Distribution of genes identified by TWAS based on physical distance to the lead variants in atrial appendage and left ventricular tissues. **d,** Canonical pathways enriched in gene sets identified by transcriptome-wide association study in atrial appendage (yellow) and left ventricular tissues (blue). TWAS, transcriptome-wide association study; GTEx, Genotype-Tissue Expression; AF, atrial fibrillation.

Next, we sought to identify transcription factors that bind to AF-associated loci which orchestrate the expression of causative genes involved in AF development. We performed enrichment analysis using the ChIP-Atlas^32^ dataset, which comprised several high-throughput ChIP-seq experiments (15,109 experiments, 1,028 transcription factors). As a result, AF-associated loci were characterized by enriched binding of 33 distinct transcription factors (*P* < 0.05) in cardiovascular cells, pluripotent stem cells-derived cardiac cells, and muscle cells (Fig. 3a and Supplementary Table 12). Of these, ERG, TAL1, and TBX5 were among the top enriched transcription factors and are known to be involved in endothelial gene expression and differentiation^46, 47^. In particular, ERG binding was significantly enriched with Bonferroni-corrected significance level of *P* = 3.3 × 10^-6^ (0.05/15,109): ERG ChIP-seq peaks overlapped with AF-associated loci around genes encoding cardiac ion channels (*SCN10*, *KCNH2*, *HCN4*, and *CAMK2D*), where active histone markers such as H3K27ac and H3K4me3 in induced pluripotent stem cells (iPSC)-derived cardiac cells were also observed (Fig. 3b and Supplementary Fig. 10). These results suggest that ERG is a transcription factor that regulates the expression of AF-associated genes.

**Fig. 3.**
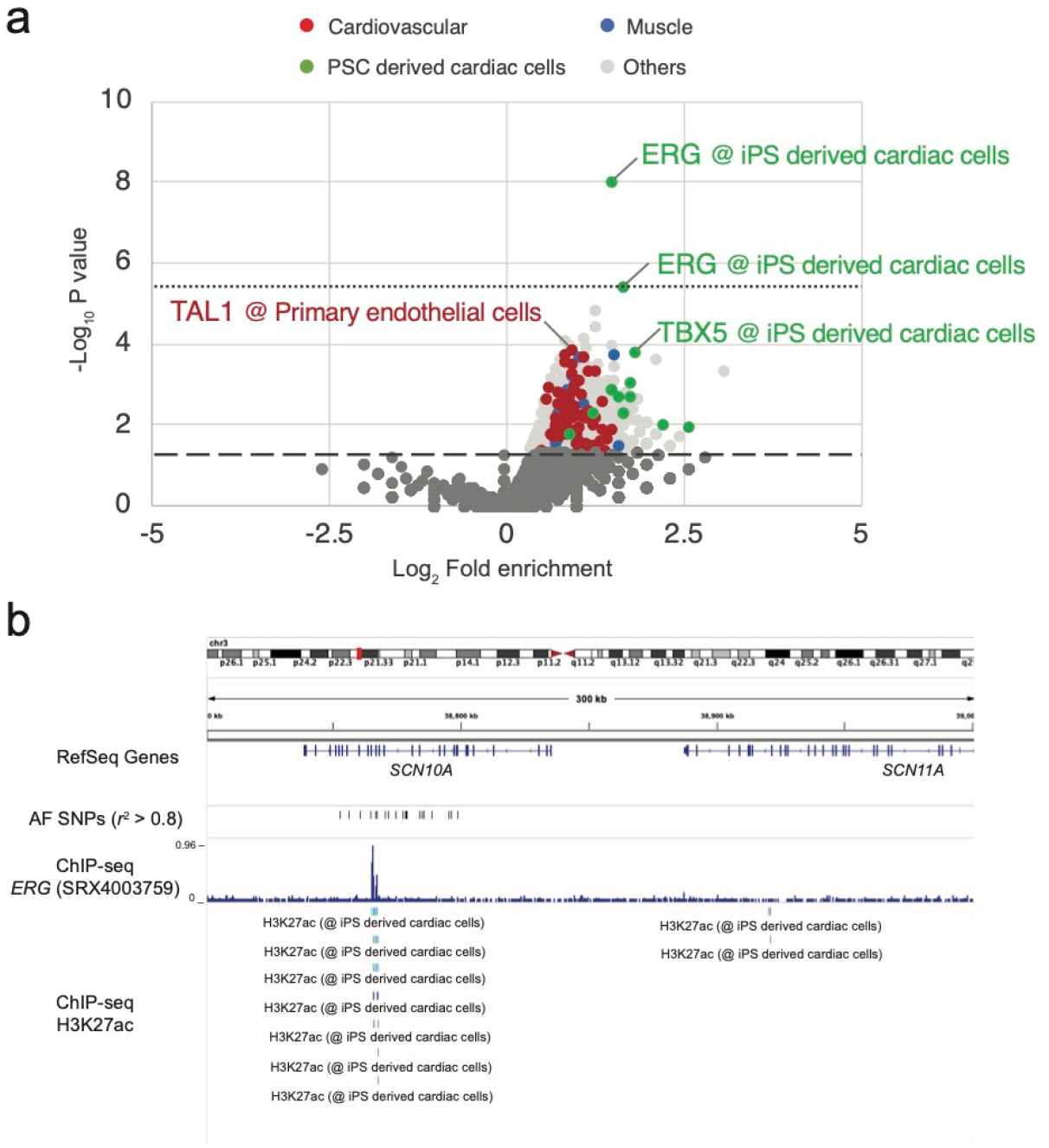
Enrichment analysis of transcription factor for AF-associated loci. **a**, Volcano plot analysis. Each point represents ChIP-seq experiment and is highlighted as red for cardiovascular cells, green for induced pluripotent stem cell-derived cardiac cells, blue for muscle cells, and grey for other cell types. The *x* axis shows log_2_-transformed hold enrichment of transcription factor in 150 AF-associated loci, compared to randomly selected 150 genome regions. The *y* axis shows log_10_-transformed *P* value for enrichment. The dashed line indicated significance threshold level of *P* = 0.05, and the dotted line indicates *P* = 0.05/15,109. **b**, *SCN10A* locus showing AF-associated variants with *r^2^* > 0.8 in European samples of 1KG and ChIP-seq track of ERG experiment (*P* = 1.0 × 10^−8^) and H3K27ac in iPSC-derived cardiac cells. AF, atrial fibrillation; iPSC, induced pluripotent stem cell.

### Predictive ability of PRS derived from single-population GWAS and trans-ancestry meta-GWAS

PRS has the potential for risk stratification of complex traits and diseases based on genetic data. However, the transferability of PRS from diverse populations to a population of another ancestry remains challenging. Therefore, we examined the performance of a PRS derived from various combinations of summary statistics in the Japanese population. We split our case-control samples into derivation, validation, and test datasets, and constructed 255 combinations of the summary statistics of three GWAS studies (BBJ, EUR, and FIN) and parameters for PRS derivation. Based on the PRS performance in the validation cohort, we determined the parameters that showed the best performance for each combination of summary statistics (BBJ, FIN, EUR, BBJ + FIN, BBJ + EUR, EUR + FIN, and BBJ + EUR + FIN) (Supplementary Table 13), and assessed the performance of the best model in the test cohort (Fig. 4, Supplementary Fig. 11, and Supplementary Table 14). Concordant with the population specificity, the PRS derived from BBJ showed a significantly better performance than those from European studies (*P* < 2.2 × 10^−16^ for both BBJ vs. EUR and BBJ vs. FIN). Furthermore, the performance of the PRS derived from meta-analysis that included different ancestry groups (EUR + BBJ and FIN + BBJ) significantly exceeded that from a single study (*P* < 2.2 × 10^−16^ for all cases) as well as that from a meta-analysis of two European studies (*P* < 2.2 × 10^−16^ for both EUR + BBJ vs. EUR + FIN and BBJ vs. FIN + EUR). Among all models, the PRS derived from three studies with multi-ancestry and the largest sample size (BBJ + EUR + FIN) showed the highest performance (pseudo *R*^2^ = 0.144, 95% CI = 0.130–0.154, area under the curve of receiver operating characteristic = 0.737, 95% CI = 0.726–0.748).

**Fig. 4.**
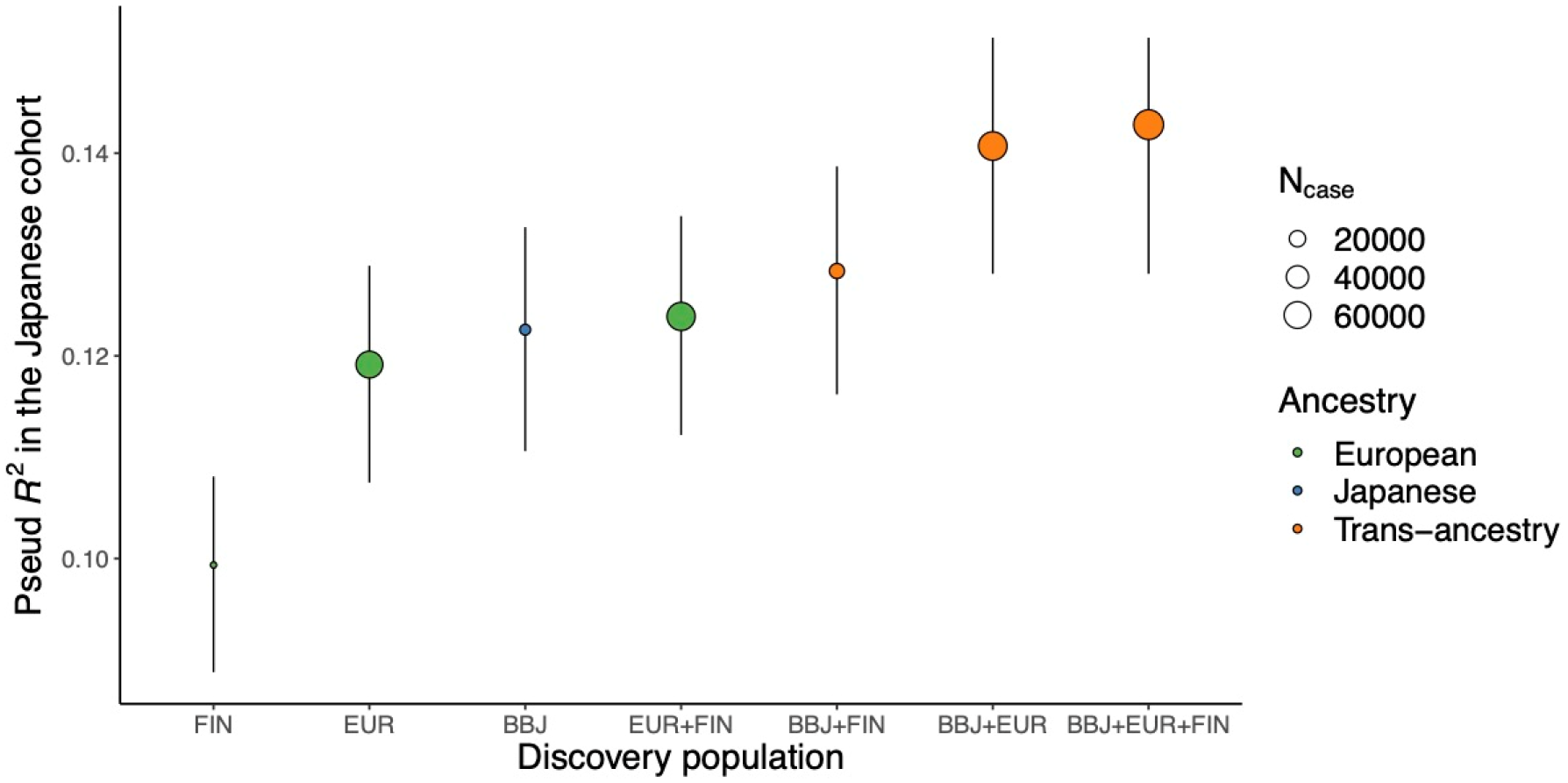
Performance of polygenic risk score (PRS) derived from the transethnic meta-analysis. Each point indicates the median Nagelkerke’s pseudo *R^2^* for PRS in the Japanese test cohort (2,953 cases and 21,194 controls). Error bars represent the 95% CI. Median pseudo *R^2^* and CI were estimated from 5 × 10^4^ times bootstrapping. The color of each point indicates the ancestry that was included in the discovery population. The size of each point indicates the number of cases in the discovery population. CI, confidence interval.

### Impact of AF-PRS on AF-relevant phenotypes and cardiovascular outcomes

To assess the potential for clinical applications of the PRS, we investigated the association between PRS and the onset age of AF in individuals from our BBJ case samples (n = 7,459). We observed that the onset age decreased as the PRS increased, and individuals with the top 1% PRS were estimated to be approximately 4 years younger in AF onset compared to the remaining individuals (Fig. 5a, 5b, and Supplementary Fig. 12a). Next, we examined whether AF-PRS could explain the phenotypic variability of stroke in individuals without a diagnosis of AF. We performed logistic regression analysis in 140,446 control samples in our dataset, and found significant associations of the PRS with increased risks of cerebral infarction (OR [95% CI] = 1.055 [1.032 – 1.078], *P* = 1.2 × 10^−6^), cardioembolic stroke (OR [95% CI] = 1.391 [1.155 – 1.676], *P* = 5.2 × 10^−4^), atherothrombotic stroke (OR [95% CI] = 1.054 [1.001 – 1.111], *P* = 0.045), and cerebral hemorrhage (OR = 1.068, 95% CI = 1.008 – 1.133, *P* = 0.027; Fig. 5c). Importantly, we observed the largest impact of the PRS on cardioembolic stroke among those with other stroke phenotypes, indicating that AF-PRS may reveal clinically undetectable AF (i.e., subclinical AF) or AF-related conditions, such as prothrombotic or hypercoagulable state, in individuals without AF.

**Fig. 5.**
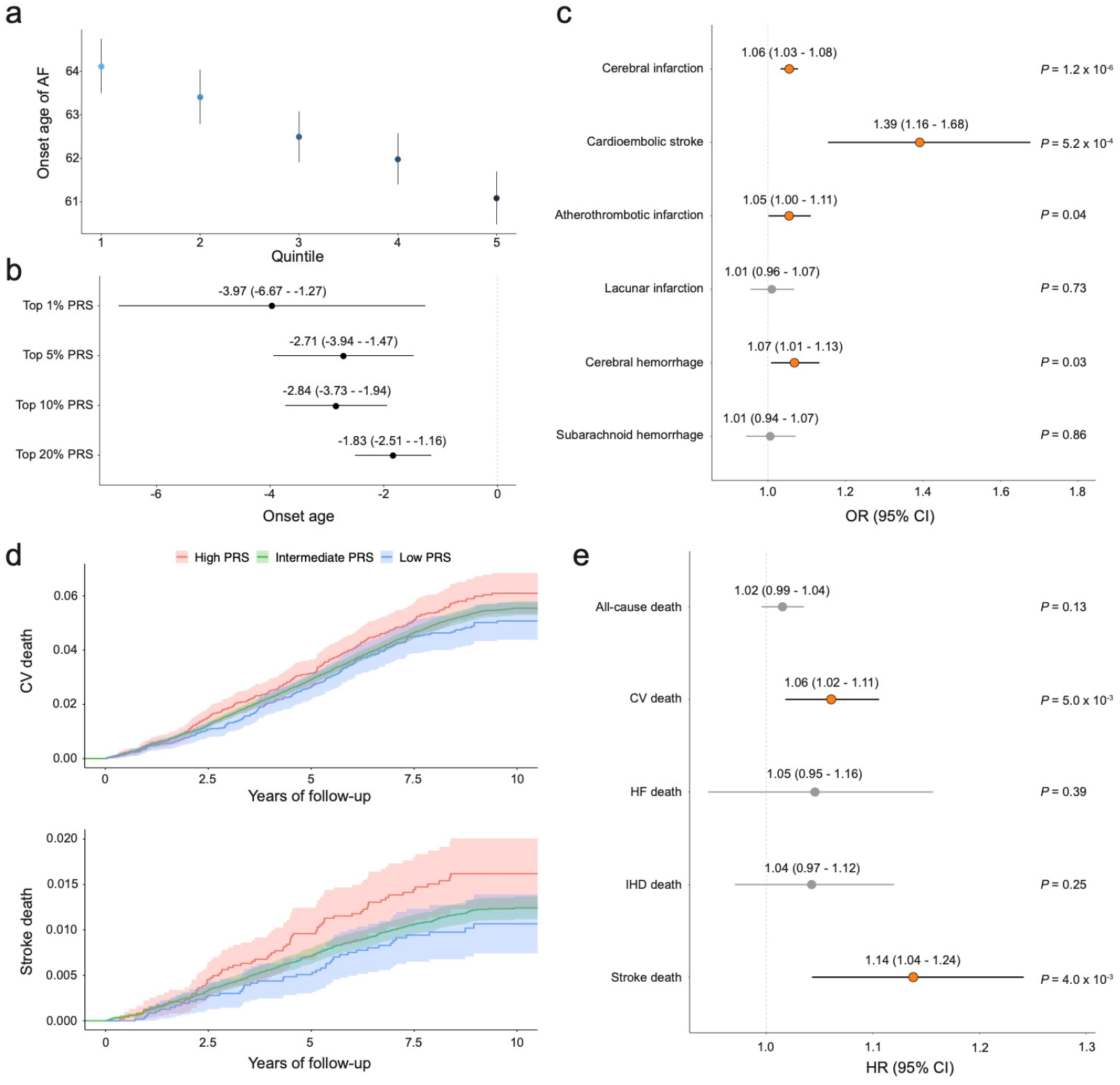
Impact of AF-PRS on long-term cardiovascular mortality. **A**, **b**, Association analysis between AF-PRS and age of AF onset. **A**, Each point represents median age of onset according to the PRS quintile. **B**, Each point and error bar represent estimated β and 95% CI obtained from the linear regression model. **C**, Association between AF-PRS and stroke subtypes in individuals without AF. Data are presented as estimated ORs and 95% CI for 1 s.d. increase in AF-PRS. **c**, **d**, Kaplan-Meier estimates of cumulative events from cardiovascular mortality (**a**) and stroke death (**b**) with 95% CI. Individuals are classified into high PRS (top 10 percentile, red), low PRS (bottom 10 percentile, blue), and intermediate (others, green). **e**, Effects of AF-PRS on long-term mortality. Data are presented as estimated HRs and 95% CIs for a 1 s.d. increase in AF-PRS. AF, atrial fibrillation; PRS, polygenic risk score; CI, confidence interval; OR, odds ratio; HR, hazard ratio; s.d., standard deviation; CV, cardiovascular death; HF, congestive heart failure death; IHD, ischemic heart disease death.

To further explore the clinical utility of AF-PRS, we assessed the impact of PRS on mortality using long-term follow-up data in the BBJ. The Kaplan-Meier estimates of cumulative mortality rate were increased in individuals with a high PRS, especially in cardiovascular- and stroke-related mortality (Fig. 5d and Supplementary Fig. 12b). Cox regression analysis demonstrated that PRS was significantly associated with an increased risk of cardiovascular death (hazard ratio [HR] per 1 standard deviation [s.d.] of PRS = 1.06, 95% CI = 1.02–1.11, *P* = 5.0 × 10^−3^) and stroke death (HR [95% CI] = 1.14 [1.04–1.24], *P* = 4.0 × 10^−3^; Fig. 5e). In contrast to evidence from clinical studies, the association between PRS and heart failure death did not reach statistical significance in the present study (HR [95% CI] = 1.05 [0.95–1.16], *P* = 0.39), because the statistical power might be hampered by a small fraction of heart failure deaths with relatively wide dispersion in our cohort.

### Genetic liability of AF to various cardiovascular diseases

AF is frequently associated with various cardiovascular diseases, such as valvular heart disease, heart failure, and stroke. The causal relationship between AF and cardiovascular diseases is clinically evident, but genetic causality is not comprehensively understood. Therefore, we estimated the directional effect of AF on a wide range of cardiovascular diseases using two-sample Mendelian randomization (MR), where the exposure was AF and all the distinct AF-associated variants from the trans-ancestry meta-analysis were used as instrumental variables. Consistent with clinical evidence, we observed significant causal effects on heart failure, cardiomyopathy, ischemic heart disease, valvular heart disease, and stroke (Fig. 6 and Supplementary Table 15). Notably, we found that AF was a significant predictor for sinus node dysfunction and atrioventricular block (OR [95% CI] = 1.480 [1.255–1.745], *P* = 3.0 × 10^-6^; OR [95% CI] = 1.253 [1.167–1.346], *P* = 6.1 × 10^-10^, respectively), indicating that genetic risk for AF contributes to the potential disturbance in the cardiac conduction system.

**Fig. 6.**
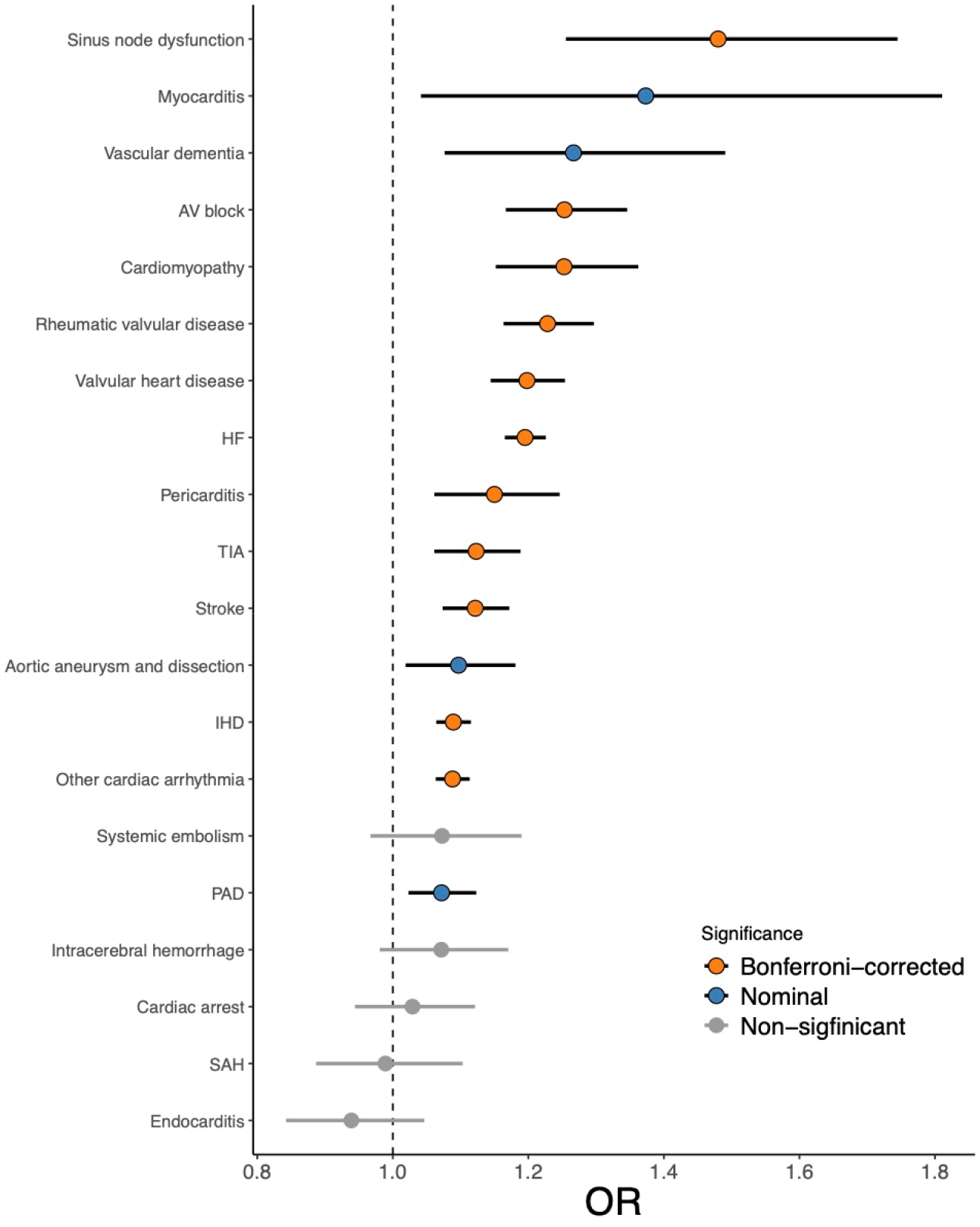
Causal effect of AF on cardiovascular and cerebrovascular diseases. Each point and error bar represent the causal estimates on the OR scale and the 95% CI for these ORs respectively. We analyzed MR analysis for the estimation of the causal effects using variants after excluding horizontal pleiotropic outliers. *P* values were determined from the IVW two-sample MR method. AF, atrial fibrillation; OR, odds ratio; CI, confidence interval; MR, sample Mendelian randomization; IVW, inverse-variance weighting; AV block, atrioventricular block; HF, heart failure; TIA, transient ischemic attack; IHD, ischemic heart disease; PAD, peripheral artery disease; SAH, subarachnoid hemorrhage.

In contrast, AF is also known as the phenotype accumulated by multiple atherosclerotic- and metabolic-related traits. Therefore, we investigated whether genetic risks for such traits could affect the development of AF using two-sample MR, where the exposures represented quantitative traits, and selected the distinct variants associated with each trait as instrumental variables. As expected^48^, height and BMI were significant predictors for AF (OR [95% CI] = 1.381 [1.166–1.635], *P* = 1.8 × 10^−4^; OR [95% CI] = 1.160 [1.099–1.225], *P* = 7.6 × 10^−8^, respectively). Furthermore, blood pressure showed a significant causal effect on AF (OR [95% CI] = 1.383 [1.292–1.480], *P* = 9.4 × 10^−21^ for systolic blood pressure; OR (95% CI) = 1.393 (1.298–1.494), *P* = 2.9 × 10^−20^ for diastolic blood pressure; OR (95% CI) = 1.220 (1.131–1.317), *P* = 3.0 × 10^−7^ for pulse pressure; Supplementary Fig. 13 and Supplementary Table 15).

## Discussion

We conducted a large-scale AF-GWAS with approximately 10,000 AF cases in the Japanese population and identified 31 genome-wide significant loci associated with AF. This includes five novel loci, where disease-relevant rare and highly East Asian-specific variants were found in the *SYNE1* and *FGF13* loci, suggesting the involvement of the functional alteration in the nuclear envelope and the ion channel as a mechanism underlying AF.

Furthermore, we performed the largest trans-ancestry meta-analysis for AF to date, where 150 genome-wide significant loci were identified, resulting in the discovery of 35 novel loci. We further analyzed AF-associated genes and transcription factors using gene expression and epigenetic datasets. Transcriptome-wide analysis linked AF-associated loci to target genes, including *IL6R*, which indicated that inflammation plays a key role in the pathogenesis of AF. Additionally, our approach based on the ChIP-seq dataset clearly implicated ERG as a candidate transcription factor involved in the regulation of AF-associated gene expression. ERG is highly expressed in endothelial cells and regulates a wide range of targets and pathways involved in vascular homeostasis and angiogenesis^49^. Endothelial dysfunction is observed in patients with AF^50, 51^, but the association between AF and ERG had not been previously examined. This may be attributable to mutational constraints on the ERG gene according to the relatively low LOEUF score of 0.33 in gnomAD and embryonic lethality in ERG knockout mice with severe defects in vascular development^52, 53^. Although the contribution of ERG signaling to AF development needs further confirmation by functional experiments, the results from this study suggest that ERG may play a critical role in the pathophysiology of AF through vascular development at early developmental stages.

We found shared allelic effects of lead variants in AF-associated loci and genetic correlations between Japanese and European populations. We then derived AF-PRS from the trans-ancestry meta-analysis, which outperformed that derived from a population-specific GWAS. In addition to the predictive ability for AF itself, AF-PRS segregated individuals with AF-related phenotypes, such as early onset of AF and cardioembolic stroke, as well as those with increased risks of long-term cardiovascular and stroke mortalities. These results indicated that the cumulative genetic risk for AF may be an indicator for early therapeutic intervention, including anticoagulation in at-risk individuals as a primary prevention of stroke. Taken together, our findings suggest the clinical utility of AF-PRS for individual risk stratification, leading to the realization of future precision medicine.

In conclusion, our large-scale Japanese and trans-ancestry genetic analyses identified 35 novel loci and provided insights into the distinct and shared genetic architecture of AF between Japanese and Europeans. Integrative analysis of transcriptome and epigenome data revealed the candidate genes and transcription factors involved in the mechanism of disease development. Furthermore, analyses of disease prediction and long-term survival demonstrated the clinical utility of the AF-PRS. These data highlight the importance of AF genetics in clinical settings and provide useful evidence for the implementation of genomic medicine.

## Methods

### Study populations

All subjects were Japanese who were registered in the BioBank Japan (BBJ) project (https://biobankjp.org/). The BBJ is a hospital-based national biobank project that collects DNA and serum samples and clinical information from 12 cooperative medical institutes throughout Japan (Osaka Medical Center for Cancer and Cardiovascular Diseases, Cancer Institute Hospital of Japanese Foundation for Cancer Research, Juntendo University, Tokyo Metropolitan Geriatric Hospital, Nippon Medical School, Nihon University School of Medicine, Iwate Medical University, Tokushukai Hospitals, Shiga University of Medical Science, Fukujuji Hospital, National Hospital Organization Osaka National Hospital, and Iizuka Hospital). Approximately 200,000 patients with any of the 47 target diseases were enrolled between 2003 and 2007. All samples were at least 18 years old. Informed consent was obtained from all participants, and our study was approved by the relevant ethical committees at each facility.

For the quality control of GWAS, we excluded samples with a call rate < 0.98 and related individuals with PI_HAT > 0.2, which is an index of relatedness based on pairwise identity-by-descent implemented in PLINK 2.0 (20 Aug 2018 ver.). We then excluded samples with a heterozygosity rate > +4 s.d. To identify population stratification, we performed principal component analysis using PLINK 2.0. and excluded outliers from the Japanese cluster. For the case samples in GWAS, we selected individuals with atrial fibrillation or atrial flutter diagnosed by a physician based on general medical practices or documented on a 12-lead electrocardiogram. The demographic features of the case–control cohort are shown in Supplementary Table 16.

### Genotyping, imputation, and quality control

GWAS subjects were genotyped using the Illumina Human OmniExpress Genotyping BeadChip or a combination of Illumina HumanOmniExpress and HumanExome BeadChips (Illumina, San Diego, CA). For genotype quality control, we excluded variants with (i) SNP call rate < 0.99, (ii) MAF < 0.01, and (iii) Hardy–Weinberg equilibrium *P* value ≤ 1.0 × 10^−6^. We pre-phased the genotypes using EAGLE and imputed dosages with the 1,000 Genome Project Phase 3 (1KG Phase 3; May 2012)^54^ reference panel with 1,037 Japanese in-house reference panel from BBJ using minimac3. For the X chromosome, pre-phasing was performed in both males and females, and imputation was performed separately for males and females. Dosages of variants in X chromosomes for males were assigned between 0 and 2.

### GWAS

In the Japanese GWAS, association was performed by logistic regression analysis assuming an additive model with adjustment for age, age^2^, sex, and top 20 principal components using PLINK 2.0. We selected variants with minimac3 imputation quality score of > 0.3 and MAF ≥ 0.001. For the X chromosome, we conducted association analyses in males and females separately, and integrated the results using an inverse-variance weighted fixed-effects model implemented in METASOFT (v2.0.1). Heterogeneity between studies was calculated using the Cochran’s Q test. We filtered variants with strong heterogeneity (*P_het_* < 1.0 × 10^-4^). The genome-wide significance threshold was defined at *P* < 5.0 × 10^-8^ for variants with MAF ≥ 1% and *P* < 5.71 × 10^-9^ for those with MAF < 1% (0.05/8,753,038 variants). Although the genomic inflation factor (*λ*_GC_) was 1.12, LD-score regression indicated that the inflation was primarily due to polygenic effects (LD-score regression intercept = 1.02; Supplementary Fig. 14a). Adjacent genome-wide significant SNPs were grouped into one locus if they were within 1 Mb from each other. We defined a locus as the candidate region that was located within 500 Mb upstream and downstream of the genome-wide significant variants. When a locus was not in LD (*r*^2^ < 0.10) with reported loci, we defined it as a novel locus. In sex-stratified association analysis of 81,050 males (6,825 cases and 74,225 controls) and 69,222 females (3,001 cases and 66,221 controls), we also performed logistic regression analysis in males and females separately using the same pipeline as described above. To investigate the difference in genetic susceptibility to AF between males and females, we compared effect sizes of the variants identified in the sex-stratified GWAS. Genotyped or imputed variants were annotated using ANNOVAR (7 July 2017 build) (http://annovar.openbioinformatics.org).

### Stepwise conditional analysis

To identify independent association signals in the loci, we conducted a stepwise conditional analysis for genome-wide significant loci defined as described in the GWAS. First, we performed logistic regression conditioning on the lead variants of each locus. We set a locus-wide significance at *P* < 1.0 × 10^-5^ and repeated this procedure until none of the variants reached locus-wide significance for each locus.

### LD score regression and heritability

To estimate confounding biases such as cryptic relatedness and population stratification, we performed an LD score regression (Version 1.0.0). We selected SNPs with MAF ≥ 0.01, and variants within the major histocompatibility complex region were excluded. For the regression, we used the East Asian LD scores provided by the authors (https://github.com/bulik/ldsc/).

### Trans-ancestry meta-analysis

Summary results from two European AF GWASs (EUR and FIN) were obtained from a previously published website (http://csg.sph.umich.edu/willer/public/afib2018)7 and from the FinnGen research project website (https://www.finngen.fi/en), respectively. EUR is a meta-analysis of six contributing AF data sources (The Nord-Trøndelag Health Study, deCODE, the Michigan Genomics Initiative, DiscovEHR, UK Biobank, and the AFGen Consortium). FIN is composed of 102,739 Finnish participants, and the phenotypes were derived from ICD codes in Finnish national hospital registries and cause-of-death registry as a part of the FinnGen project. To inspect the biases of these GWAS, we calculated the LD-score regression intercept for each study and confirmed that these two studies were well calibrated (LD score regression intercept for EUR = 1.052 (s.e.m. = 0.012) and FIN = 1.033 (s.e.m. = 0.010); Supplementary Fig. 14b, c). We also calculated the genetic correlation and found a significant genetic correlation between EUR and FIN (*r*_g_ = 0.918, s.e.m. = 0.035, *P* = 3.9 × 10^−1^^55^).

To account for the ancestral heterogeneity among the three studies, we applied the MANTRA algorithm in the trans-ancestry meta-analysis^55^, which allows for heterogeneity between diverse ancestry groups and improves performance compared to fixed-effects meta-analysis and random-effects meta-analysis. Variants with MAF ≥ 1% in both the Japanese and European populations were selected for association. We considered SNPs with log_10_ BF > 6 to be genome-wide significant.

### Trans-ancestry genetic correlation

To estimate the trans-ancestry genetic correlation, we applied Popcorn software (v.1.0). We used East Asia or European samples of 1KG for the LD reference and excluded the variants within the major histocompatibility complex region according to the instructions of the software. To estimate the liability-scale genetic correlation, we assumed that the AF prevalence was 0.58% in the Japanese population, 1.38% in the United Kingdom population, and 0.97% in the Finnish population (https://vizhub.healthdata.org/gbd-compare/).

### Credible set analysis

To construct sets of variants that likely include causal variants, we constructed a 99% credible set in each AF-associated locus using BF obtained from MANTRA results in the trans-ancestry meta-analysis. For each locus, we calculated the posterior probability (PP) for the *j*^th^ SNP using the following formula: 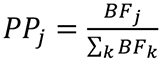, where *BF_j_* denotes the BF for the *j*^th^ SNP and *BF_k_* denotes all of the variants included in the locus. We then constructed the 99% credible set by adding the variants in order of decreasing PP until the cumulative PP reached 0.99. To assess whether the 99% credible set derived from the trans-ancestry meta-analysis narrowed down the causal variants, we performed two additional meta-analyses (EUR + BBJ and EUR + FIN) using the MANTRA algorithm and constructed 99% credible sets for 150 AF-associated loci identified by the trans-ancestry meta-analysis. We then compared the number of variants in the 99% credible set for each locus. Among them, the loci where at least one variant surpassed genome-wide significance (log_10_ BF > 6) in all meta-analyses were included in the test. The differences in the sizes of the credible sets were tested using the paired Wilcoxon rank-sum test.

### Pleiotropy

We searched for AF-associated variants that overlapped with other diseases or traits using the NHGRI-EBI GWAS catalog on February 8, 2019. As reported previously^34^, we selected the AF-associated variants in the trans-ancestry meta-analysis according to the following criteria: (i) variants with *r*^2^ > 0.5 in both East Asian and European samples of 1KG if the publication in GWAS catalog included East Asians, (ii) variants with *r*^2^ > 0.5 in European samples of 1KG if the original report did not study East Asian populations; and (iii) the physical distance from AF-associated lead variants in the trans-ancestry meta-analysis to variants reported in the GWAS catalog was within 25 kb.

For the association analysis between AF-associated loci and QTL for clinical parameters, we used the summary statistics previously reported in the BBJ dataset^34–36^. We selected AF-associated variants (*r*^2^ > 0.6 in East Asian samples of 1KG and clinical parameters of nine distinct categories: anthropometric (n=2), metabolic (n=6), serum protein (n=2), kidney-related (n=4), electrolyte (n=3), liver-related (n=5), other biochemical (n=5), hematological (n=5), and blood pressure (n=4). In QTL analysis, clinical measurements were corrected by taking into account the effect of medications as follows: for individuals taking cholesterol-lowering drugs, total cholesterol and low-density-lipoprotein cholesterol levels were corrected by dividing by 0.8 and 0.7, respectively^35^; for individuals taking antihypertensive drugs, 15 mmHg was added to the systolic blood pressure and 10 mmHg to the diastolic blood pressure readings^35^. Furthermore, linear regression models for these quantitative traits were adjusted for sex, age, age^2^, the top 10 principal components, and disease status. Smoking status was also introduced into the model for white blood cell count and C-reactive protein levels. We chose QTL showing a genome-wide significant association (*P* < 5.0 × 10^-8^).

### Tissue and gene set enrichment analysis

For tissue and gene set enrichment analysis, we used data-driven expression-prioritized integration for complex traits (DEPICT) software (https://data.broadinstitute.org/mpg/depict)39. To reduce the redundancy of the pathways, we applied affinity propagation clustering implemented in the apcluster package v.1.4.8 in R ^56,57^. Using this algorithm, we obtained distinct clusters that contained ‘exemplar’ pathway and member pathways. We extracted 48 exemplar tissues and 1,157 gene sets for enrichment analysis and selected independent AF-associated variants with log_10_ BF > 5 as applied in a previous study^58^. The significance level was set at *P* = 1 × 10^−3^ (=0.05/48) for tissue enrichment analysis and *P* = 4.3 × 10^−5^ (=0.05/1,157) for gene set enrichment analysis.

### Transcriptome-wide association study

We performed a transcriptome-wide association study to assess gene-AF associations using MetaXcan v0.3.512^59^, which estimates the association between predicted gene expression levels and a phenotype of interest using summary statistics and gene expression prediction models. We used precomputed prediction models with LD reference data in GTEx v7 for atrial appendage and left ventricular tissues and the summary statistic of the trans-ancestry AF-GWAS as input. Bonferroni significance level was set at *P* = 8.4 × 10^-6^ (=0.05/5,988) for atrial appendage and *P* = 9.3 × 10^-6^ (=0.05/5,378) for left ventricle to account for the number of genes tested in each tissue.

### Enrichment analysis of transcription factor

To assess the enrichment of transcription factor in AF-associated loci, we defined AF-associated loci as regions within 500 Mb upstream and 500 Mb downstream of the AF-associated lead variants or proxies with *r*^2^ > 0.8 in European samples of 1KG. We then searched for overlaps of peak-call data archived in the ChIP-Atlas dataset with AF-associated loci and control regions selected from all the genome regions by permutation test. *P*-values were calculated with the two-tailed Fisher’s exact probability test (the null hypothesis is that the two regions overlap with the ChIP-Atlas peak-call data in the same proportion). The epigenetic landscapes around cardiac ion channel-related genes were visualized using ChIP-Atlas Peak Browser and Integrative Genomics Viewer^60^.

### PRS derivation and performance

We derived PRS using the pruning and thresholding methods. PRSs were created over a range of *P* value threshold (0.5, 5.0 × 10^-2^, 5.0 × 10^-4^, 5.0 × 10^-6^, and 5.0 × 10^-8^) and *r*^2^ threshold (0.2, 0.5, and 0.8). For the survival analysis, we divided our dataset into three groups: (i) a discovery group to derive and validate PRS (6,890 cases and 49,451 controls), (ii) a test group to assess PRS performance (2,953 cases and 21,194 controls), and (iii) a group for the survival analysis (70,645 controls) (Supplementary Fig. 15). To secure independence between the PRS derivation and validation, we used a ten-fold cross-validation approach. First, we randomly split a discovery group into ten subgroups and used nine of these subgroups for PRS derivation and the remaining one for PRS validation. To determine the weight of PRS, we performed GWAS in combination with two European GWAS: (i) Japanese subgroup GWAS, (ii) two European GWAS (EUR or FIN), (iii) Japanese subgroup GWAS + EUR (fixed- or random-effect model), (iv) Japanese subgroup GWAS + FIN (fixed- or random-effect model), (v) EUR + FIN (fixed- or random-effect model, and (vi) Japanese subgroup GWAS + EUR + FIN (fixed- or random-effect model). Meta-analyses were performed using the METASOFT software. In the meta-analysis with the Japanese subgroup GWAS, LD was calculated separately based on East Asian or European samples of 1KG. We then calculated the PRSs for the withheld validation cohort using PLINK 2.0 and validated its performance. Changing the withheld validation cohort, we repeated these procedures ten times and obtained the performance measurements for each combination. Notably, the validation cohort was excluded from the derivation of GWAS. The performance of the PRS was measured as (1) Nagelkerke’s pseudo *R^2^* obtained by modeling age, sex, and normalized PRS, and (2) the area under the curve of the receiver operator curve in the same model as Nagelkerke’s pseudo *R^2^*. The best model/parameter set for each combination model (BBJ, EUR, FIN, BBJ + EUR, BBJ + FIN, EUR + FIN, BBJ + EUR + FIN) was determined by averaging Nagelkerke’s pseudo *R^2^* (Supplementary Table 13). Using the best models and parameters determined in the derivation and validation cohorts, we calculated the PRSs and assessed their performance for the independent test cohort. We applied bootstrapping to assess the distribution of performance of the PRS; randomly extracted the individuals in the test cohort with replacement and calculated the performance for the samples. By repeating this procedure 5.0 × 10^4^ times, we estimated the distribution of the performance measures.

### Association of AF-PRS with age of AF onset and stroke subtypes

To assess the association between AF-PRS and age at AF onset, we extracted AF case samples with the data (n = 7,458, median age of AF onset was 63 years of age (IQR 56 – 71)), and estimated the difference in the effect of AF onset age between individuals with top PRS to those with the remaining PRS using a linear regression model adjusted for sex and top 10 principal components. For the association analysis between AF-PRS and stroke subtype, we calculated ORs and associated *P* values using a logistic regression model adjusted for age, sex, and the top 10 principal components. Among the control samples (without AF diagnosis) in our dataset (n = 140,447), we found 14,120 stroke phenotypes: 9,177 for cerebral infarction, 111 for cardioembolic stroke, 1,465 for atherothrombotic infarction, 1,266 for lacunar infarction, 1,143 for cerebral hemorrhage, and 967 for subarachnoid hemorrhage.

### Survival analysis

For survival analysis, the Cox proportional hazards model was used to assess the association between AF-PRS and long-term mortality. We obtained survival follow-up data with the ICD-10 code for 132,737 individuals from the BBJ dataset^61, 62^. The causes of death were classified according to ICD-10 codes: (i) cardiovascular death (I00–I99), (ii) heart failure death (I50), (iii) ischemic heart disease death (I20–I25), and (iv) stroke death (I60, I61, I63, and I64). The median follow-up period was 8.4 years (IQR 6.8 – 9.9). The Cox proportional hazards model was adjusted for sex, age, top 10 principal components, and disease status. Analyses were performed with the R package survival v.2.44, and survival curves were estimated using the R package survminer v.0.4.6, with modifications.

### Mendelian randomization

We performed two-sample MR to test for a causal relationship between AF and relevant diseases or traits. Although the statistical power to detect the associations with AF decreased, we selected AF-associated variants from the trans-ancestry meta-analysis in combination with BBJ and FIN to avoid sample overlap, and extracted summary statistics from a large database of genetic associations in the UK biobank (http://www.nealelab.is/uk-biobank/). To identify independent variants for exposure, genome-wide significant variants (*P* < 5 × 10^−8^) were pruned (*r^2^* < 0.05; LD window of 10,000 kb; using European samples of 1KG for LD reference) as applied previously^63^. First, we assessed whether AF is causally associated with cardiovascular diseases using AF-associated variants as instrument variables and variants associated with cardiovascular diseases as outcome variables. Next, we assessed the causal effects of quantitative traits related to anthropometry, metabolites, serum protein, kidney function, liver function, hematocyte count, and blood pressure on AF, where variants associated with quantitative traits were used as instrument variables and AF-associated variants as outcome variables. We used the inverse-variance-weighting (IVW) method. To exclude horizontal pleiotropic outliers, we performed MR-PRESSO for instrument variables^64^. We also calculated Cochran’s Q statistics for heterogeneity between the causal effects of variants using IVW and the MR-Egger intercept for directional pleiotropy. All analyses were performed using R v4.0.3, with R packages TwoSampleMR and MRPRESSO.

## Data Availability

Summary statistics of Japanese GWAS and polygenic risk scores derived in this study will be publicly available at the National Bioscience Database Center (https://humandbs.biosciencedbc.jp/).

https://humandbs.biosciencedbc.jp/

## Code Availability

Code used in this study is available on reasonable request to the corresponding authors.

## Acknowledgements

We thank the staff of BBJ for their assistance in collecting samples and clinical information. We also thank A. P. Morris for providing us with MANTRA software and valuable advice. This research was funded by the GRIFIN project of the Japan Agency for Medical Research and Development (AMED) (nos. JP20km0405209 and JP20ek0109487 to K. Miyazawa, K. I, S. K., S. N., H. Akazawa, and I. K.), MSD Life Science Foundation (to K. Miyazawa), the Japan Society for the Promotion of Science (to K. Miyazawa, K. I. and H. I.), and Sakakibara Memorial Research Grant from The Japan Research Promotion Society for Cardiovascular Diseases (to H. M.). BBJ was supported by the Tailor-Made Medical Treatment Program of the Ministry of Education, Culture, Sports, Science, and Technology (MEXT) and AMED under grant numbers JP17km0305002 and JP17km0305001.

## Author Contributions

K. Miyazawa, K. I., C. T., Y. K., H. Akazawa, and I. K. conceived and designed the study. C. T., K. Matsuda, Y. M., M. K., and Y. K. collected and managed the BBJ sample. Y. M., A. T., M. K., and Y. K. performed the genotyping. K. Miyazawa, K. I., Z. Z., H. M., C. T., S. O., and Y. K. performed the statistical analyses. K. Miyazawa, K. I., Z. Z., H. M., H. I., M. A., K. O., Y. O., C. T., and S. O. contributed to data processing, analysis, and interpretation. K. I., H. Aburatani, H. Akazawa, Y.K. and I. K. supervised the study. K. Miyazawa, K. I., and M. H. wrote the manuscript, and many authors have provided valuable edits.

---

**Supplementary Fig. 1.**
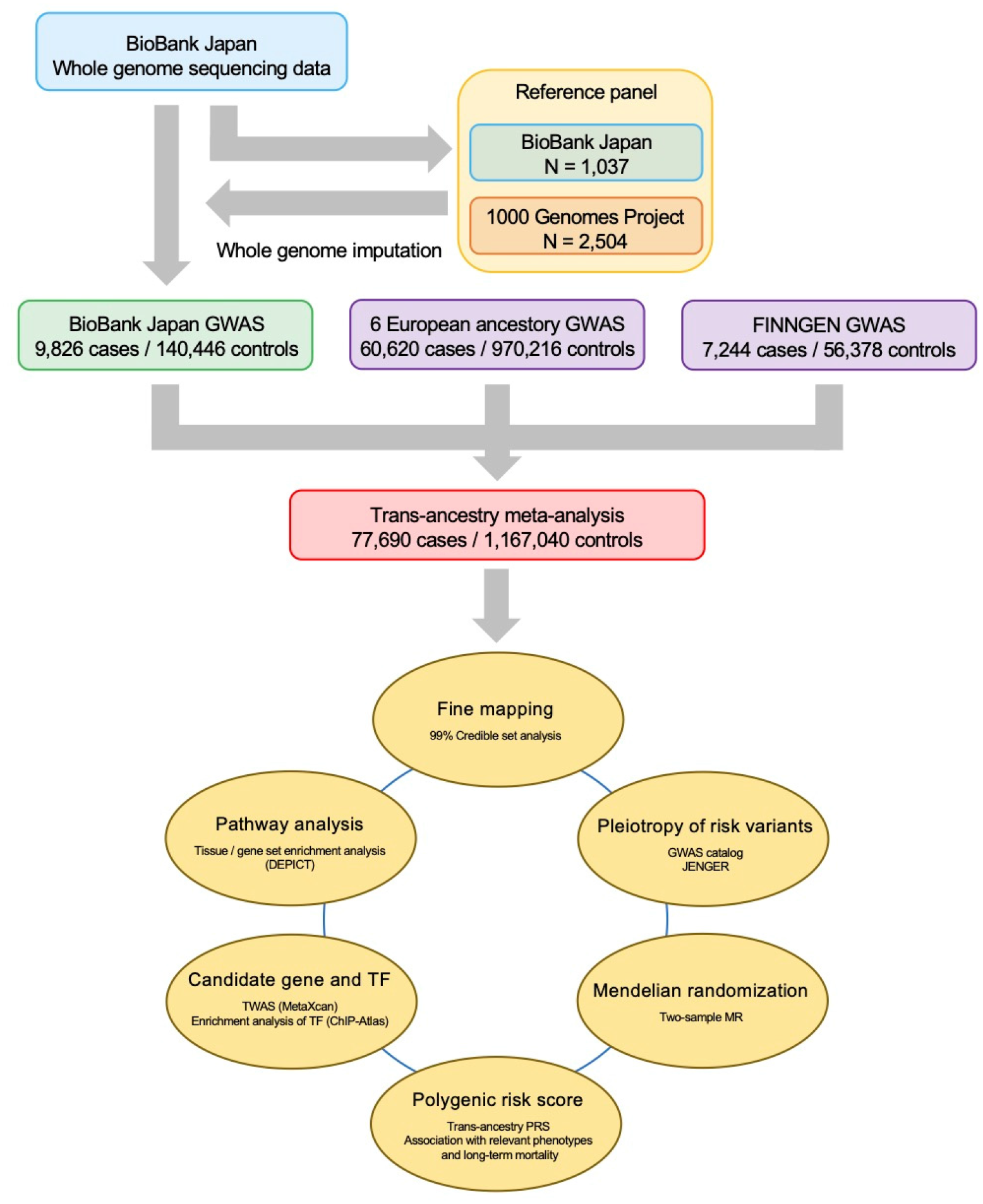
Overview of the study design. Top, overview of the Japanese cohort of 9,826 AF cases and 140,446 controls with reference panel. Middle, GWAS summary of each study and the trans-ancestry meta-analysis with the number of cases and controls. Bottom, overview of the downstream analyses for AF-associated variants in the trans-ancestry meta-analysis. GWAS, genome-wide association study; JENGER, Japanese ENcyclopedia GEnetic associations by Riken; MR, Mendelian randomization; PRS, polygenic risk score; TWAS, transcriptome-wide association study.

**Supplementary Fig. 2.**
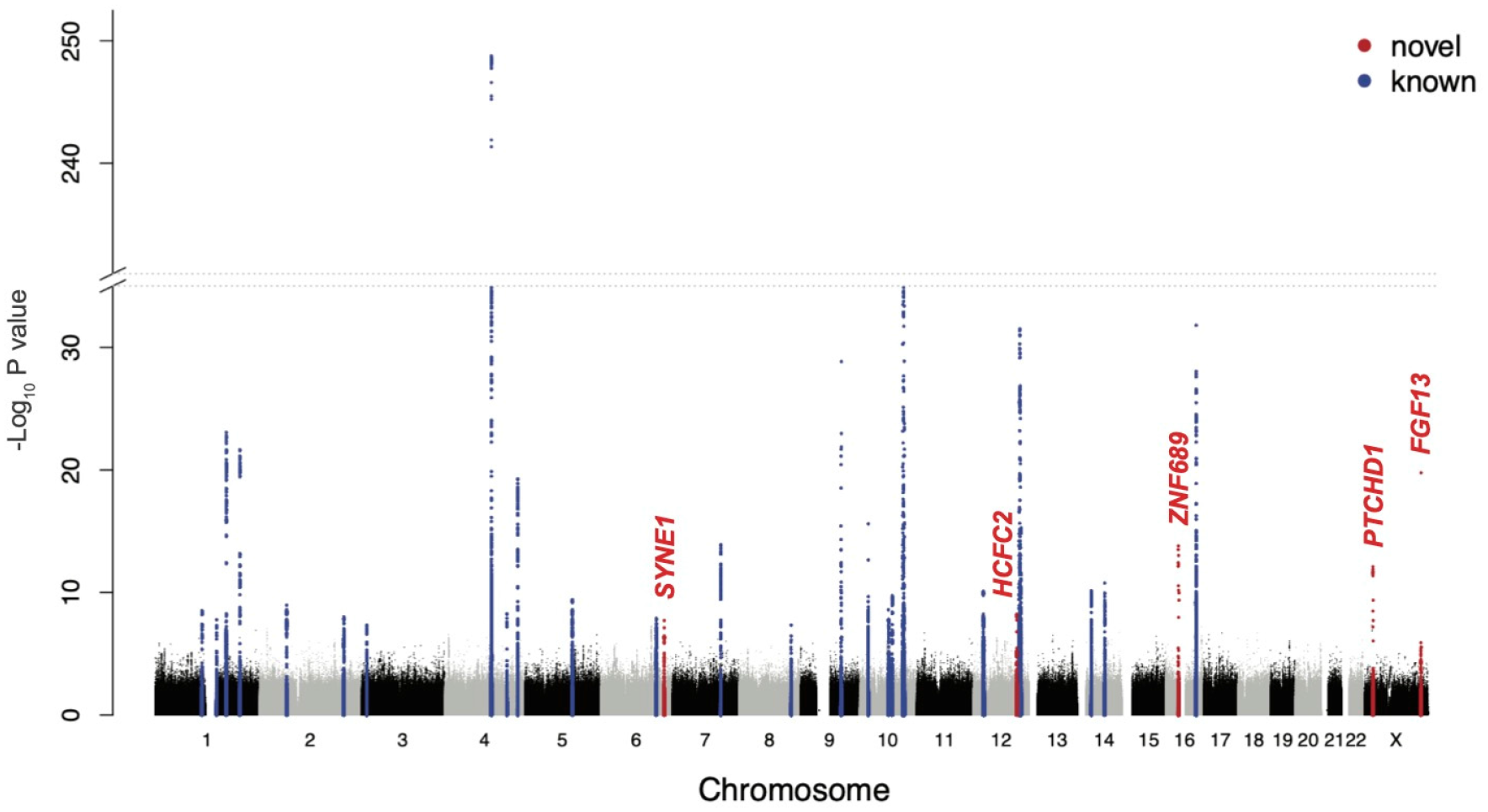
Manhattan plot for the Japanese GWAS. The results of the Japanese GWAS (9,826 AF cases and 140,446 controls) are shown. The negative log_10_ *P* values on the *y-axis* are shown against the genomic positions (hg19) on the *x-axis*. Association signals that reached a genome-wide significance level (*P* < 5.0 × 10^−8^) are shown in blue if previously reported loci and in red if novel loci. GWAS, genome-wide association study; AF, atrial fibrillation.

**Supplementary Fig. 3.**
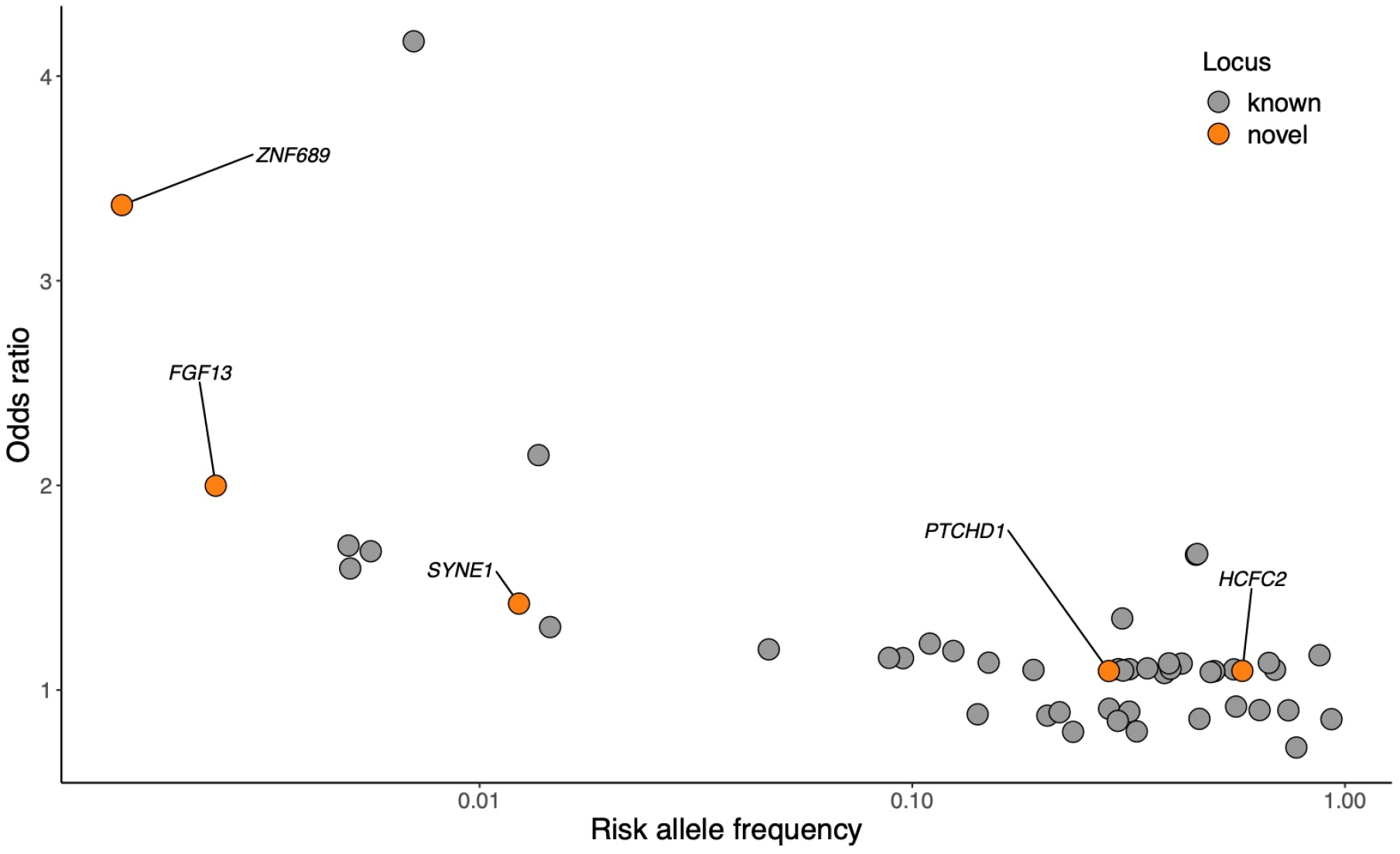
Independent association signals in AF development. The ORs for AF development of AF 49 independent signals in the Japanese GWAS (31 lead variants and 18 independent variants) were plotted against the risk allele frequencies. Novel variants are highlighted in orange with annotated genes. AF, atrial fibrillation; OR, odds ratio; GWAS, genome-wide association study.

**Supplementary Fig. 4.**
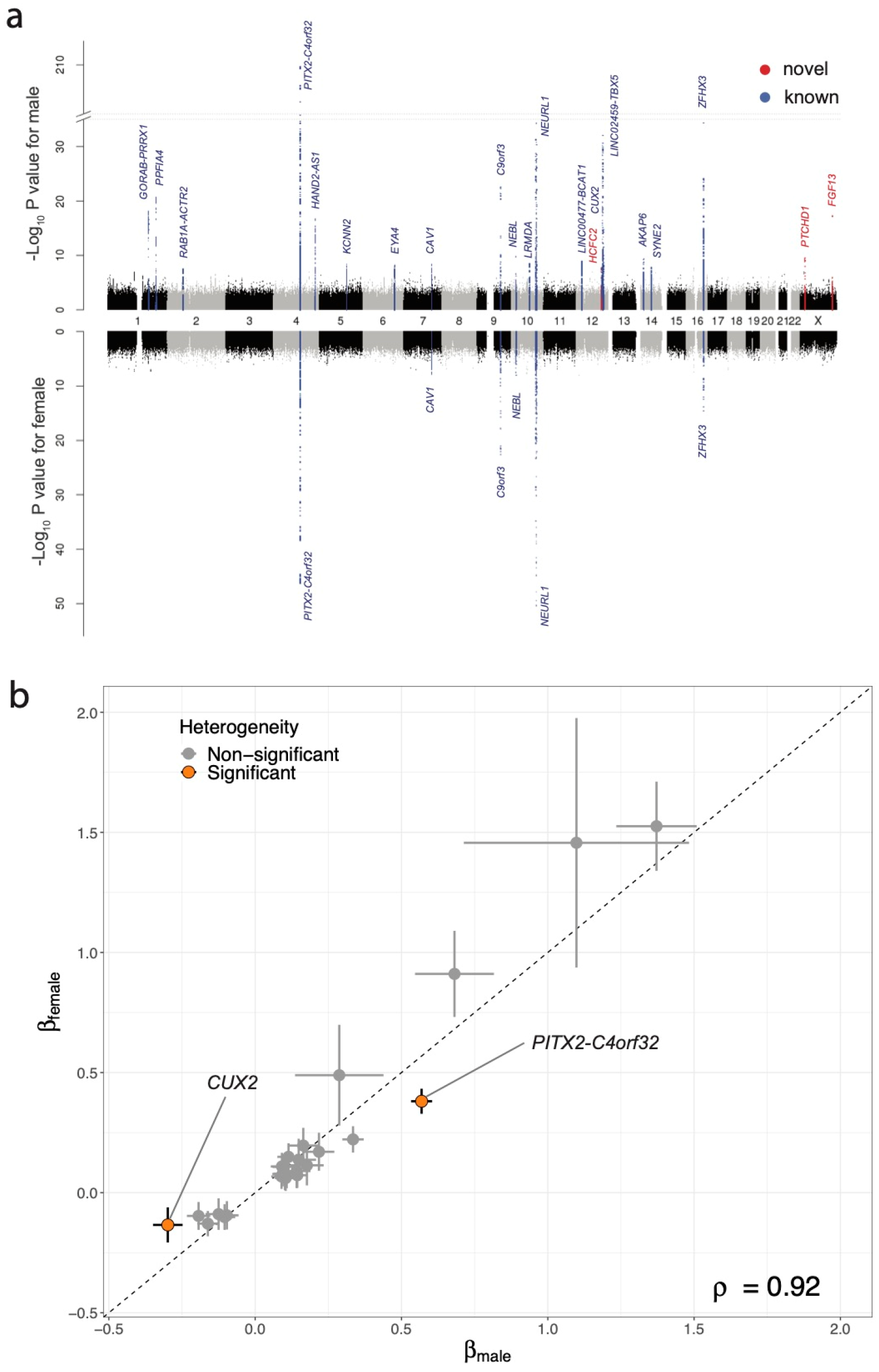
Sex-stratified GWAS. **a**, Miami plot for sex-stratified GWAS. Upper panel for male GWAS (N = 81,050: 6,825 cases and 74,225 controls) and lower panel for female GWAS (N = 69,222: 3,001 cases and 66,221 controls). Association signals that reached a genome-wide significance level (*P* < 5.0 × 10^−8^) are shown in blue if previously reported loci and in red if novel loci. **b**, Comparison of estimated effect sizes of the lead variants identified in both male and female GWAS. Data are presented as estimated β and 95% confidence. The lead variants with significant heterogeneity (*P_het_* < 0.0001) are highlighted in orange, and ρ indicates Spearman’s correlation coefficient. GWAS, genome-wide association study.

**Supplementary Fig. 5.**
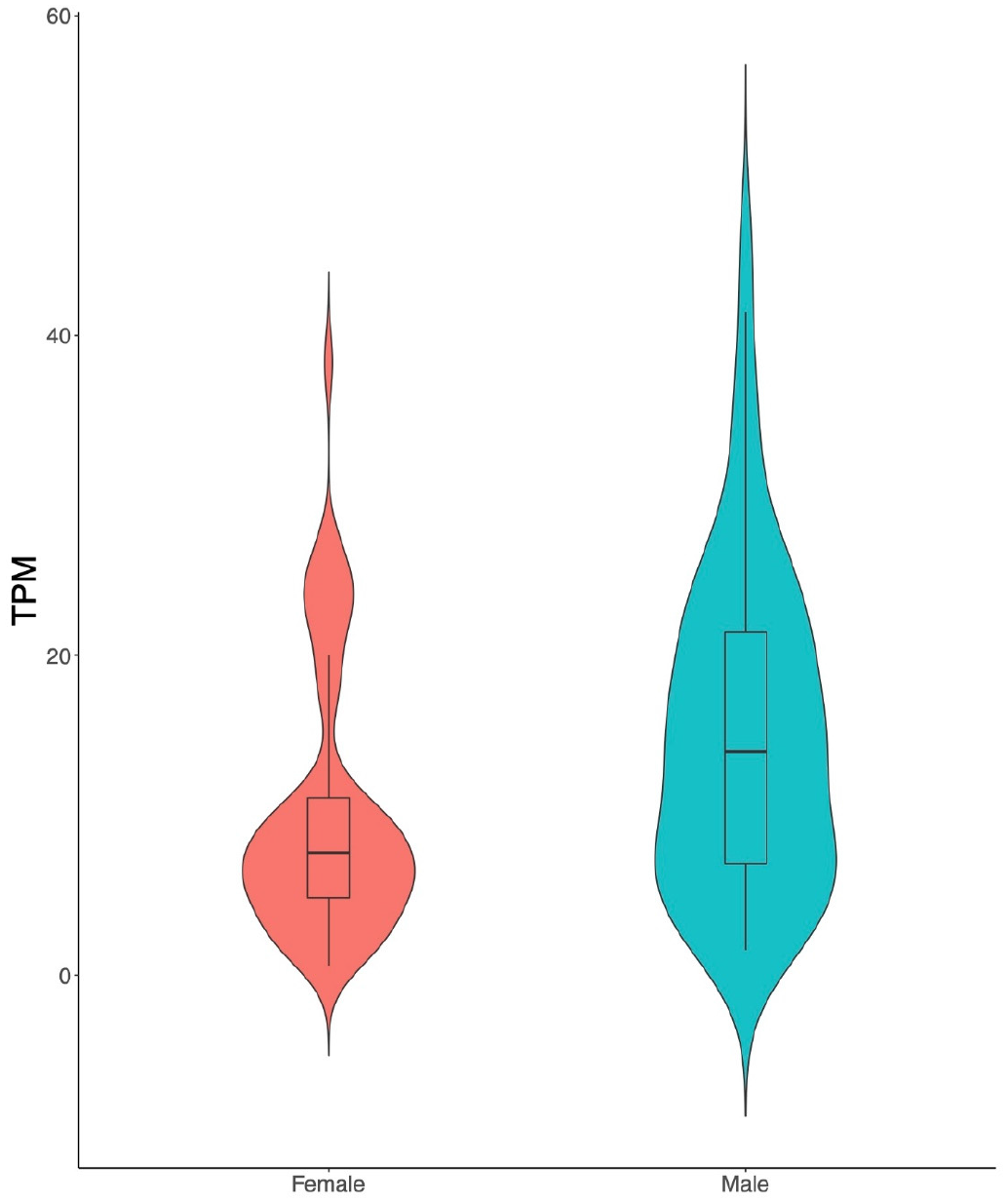
*CUX2* gene expression in liver. Violin plots showing the distribution of *CUX2* gene expression in the liver based on GTEx data.

**Supplementary Fig. 6.**
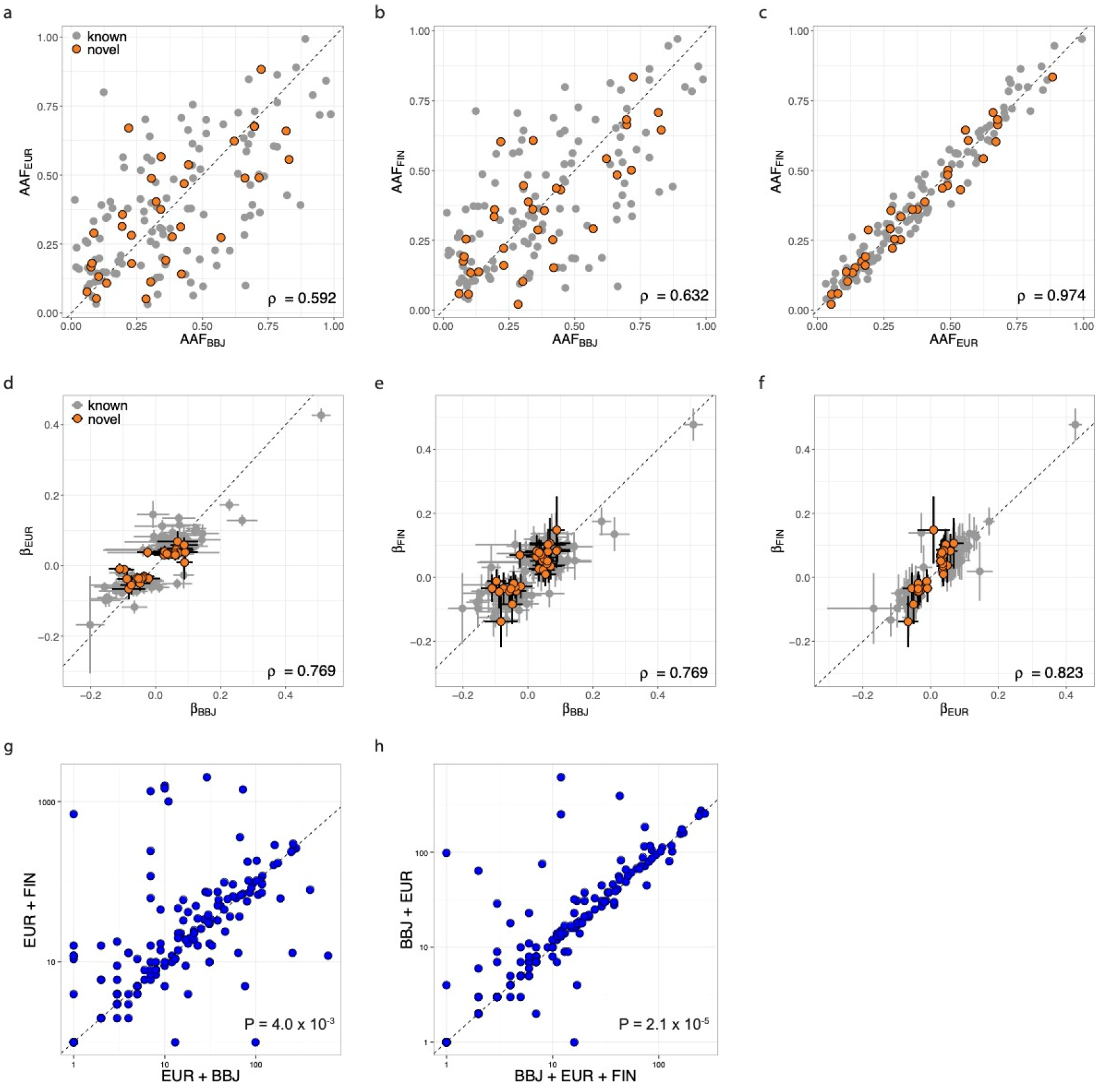
Comparison of allele frequencies and allelic effects among ancestries and fine mapping derived from credible set analyses. **a-c**, Comparisons of alternate allele frequencies of the 150 lead variants identified in the trans-ancestry meta-analysis (**a**, BBJ versus EUR; **b**, BBJ versus FIN; **c**, EUR versus FIN). **d-f**, Comparison of estimated effect sizes of 150 lead variants. Data are presented as estimated β and 95% CI in each study (**d**, BBJ versus EUR; **e**, BBJ versus FIN; **f**, EUR versus FIN). Grey points indicate previously reported loci, and orange points indicate newly identified loci in this study. ρ indicates Spearman’s correlation coefficient. **g**, **h**, Comparisons of the number of variants included in the 99% credible sets derived from each combination of the meta-analysis (**g**, EUR + BBJ versus EUR + FIN; **h**, BBJ + EUR + FIN versus BBJ + EUR). Both *the x* and *y* axes represent the log-transformed number of variants in the 99% credible sets. BBJ, BioBank Japan; EUR, European population; FIN, FinnGen data release 2; CI, confidence interval.

**Supplementary Fig. 7.**
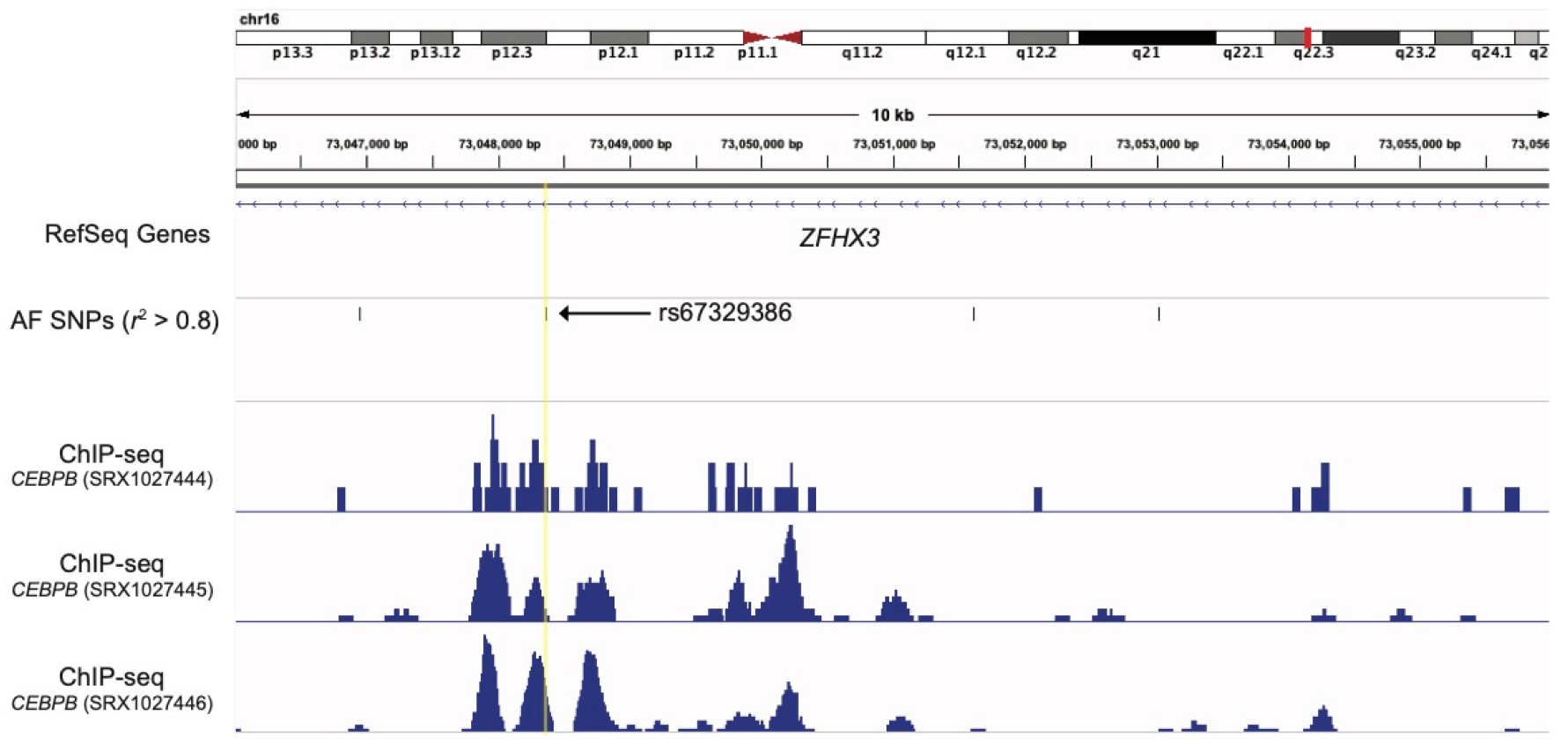
rs67329386 and transcription factor. *ZFHX3* locus showing rs67329386 (highlighted in yellow) and proxies with *r*^2^ > 0.8 in European samples in 1KG along with ChIP-seq track of CEBPB experiments in mesenchymal stem cells.

**Supplementary Fig. 8.**
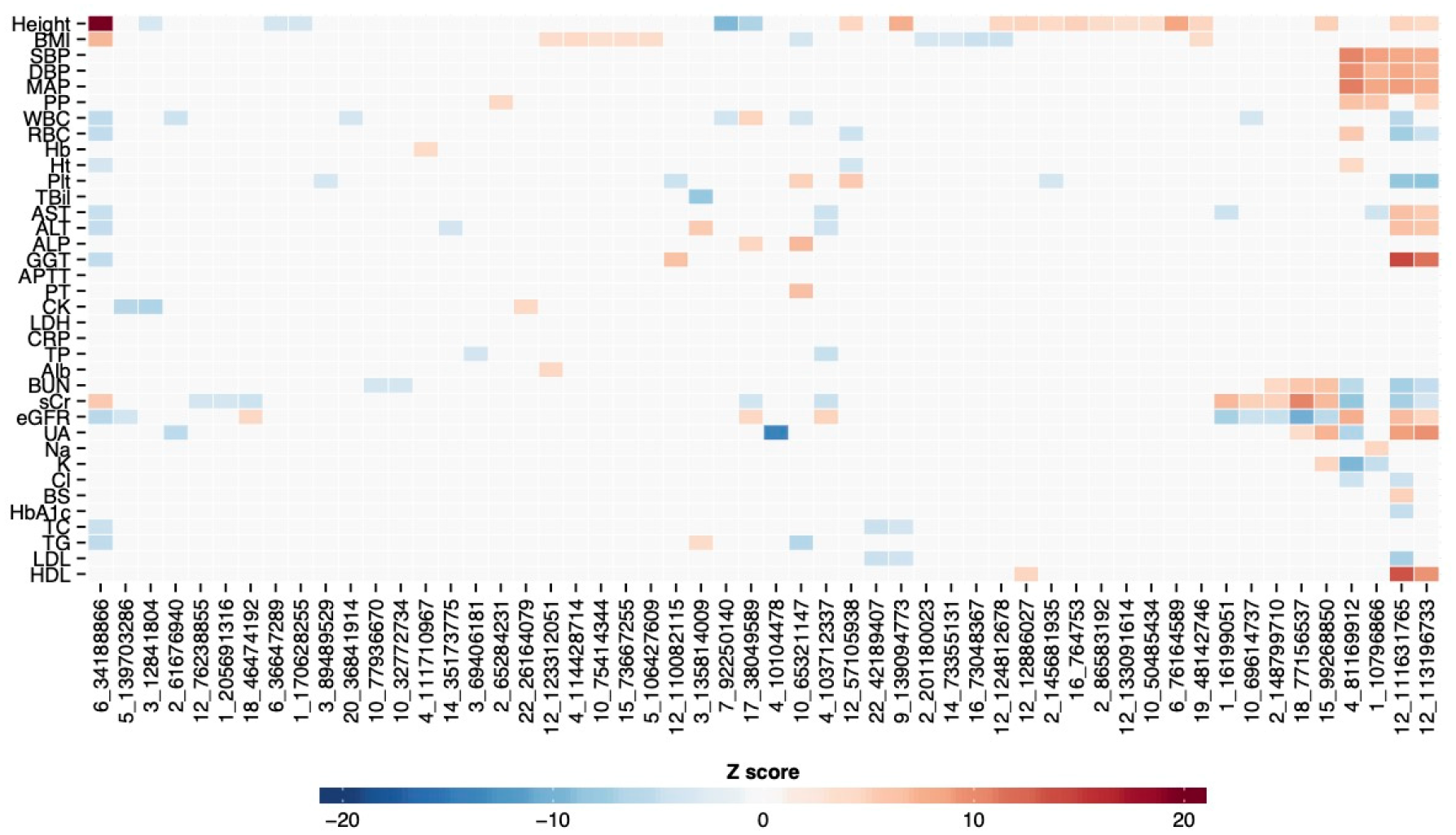
Overlap between AF-associated signals and variants associated with quantitative traits. Heat-map representation of the AF-associated loci, in which AF-associated variants in LD with *r^2^* > 0.6 in East Asian samples of 1KG overlapped with variants associated with at least one of the quantitative traits. The Z score of QTL alleles that were in accordance with the AF risk allele are shown. Positive values are shown in red, and negative values are shown in blue. The rows show quantitative traits, and the column shows the chromosome and base position of AF-associated signals. AF, atrial fibrillation; LD, linkage disequilibrium; QTL, quantitative trait loci.

**Supplementary Fig. 9.**
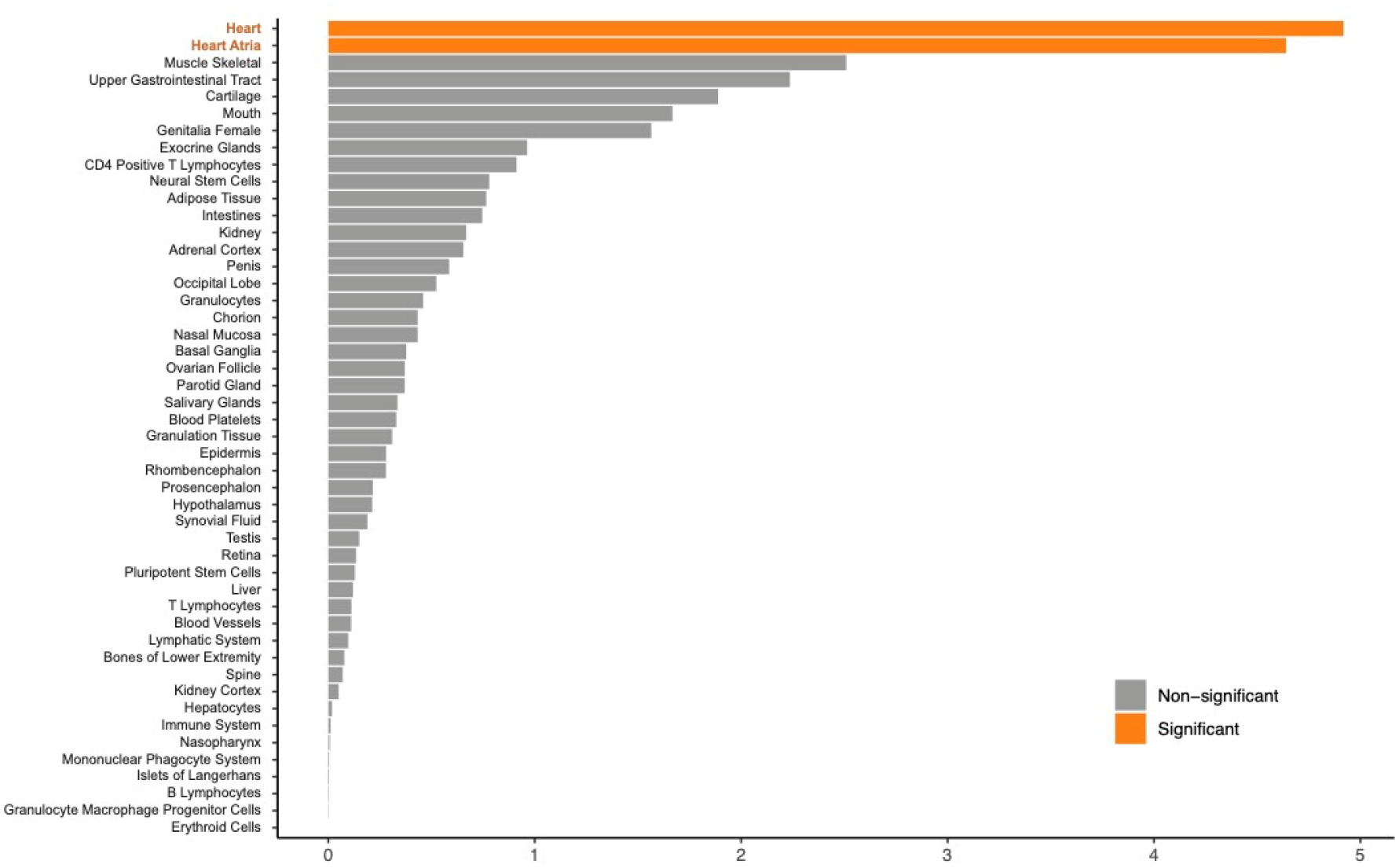
Tissue enrichment and eQTL analysis. The results of tissue enrichment analysis (log_10_ BF > 5 and 16,181 variants). The *x-axis* indicates the -log_10_ *P* value. Forty-eight exemplar tissues (Methods) were tested, and the significance level was set at *P* = 1 × 10^−3^ (0.05/48). eQTL, expression quantitative trait locus; BF, Bayes factor.

**Supplementary Fig. 10.**
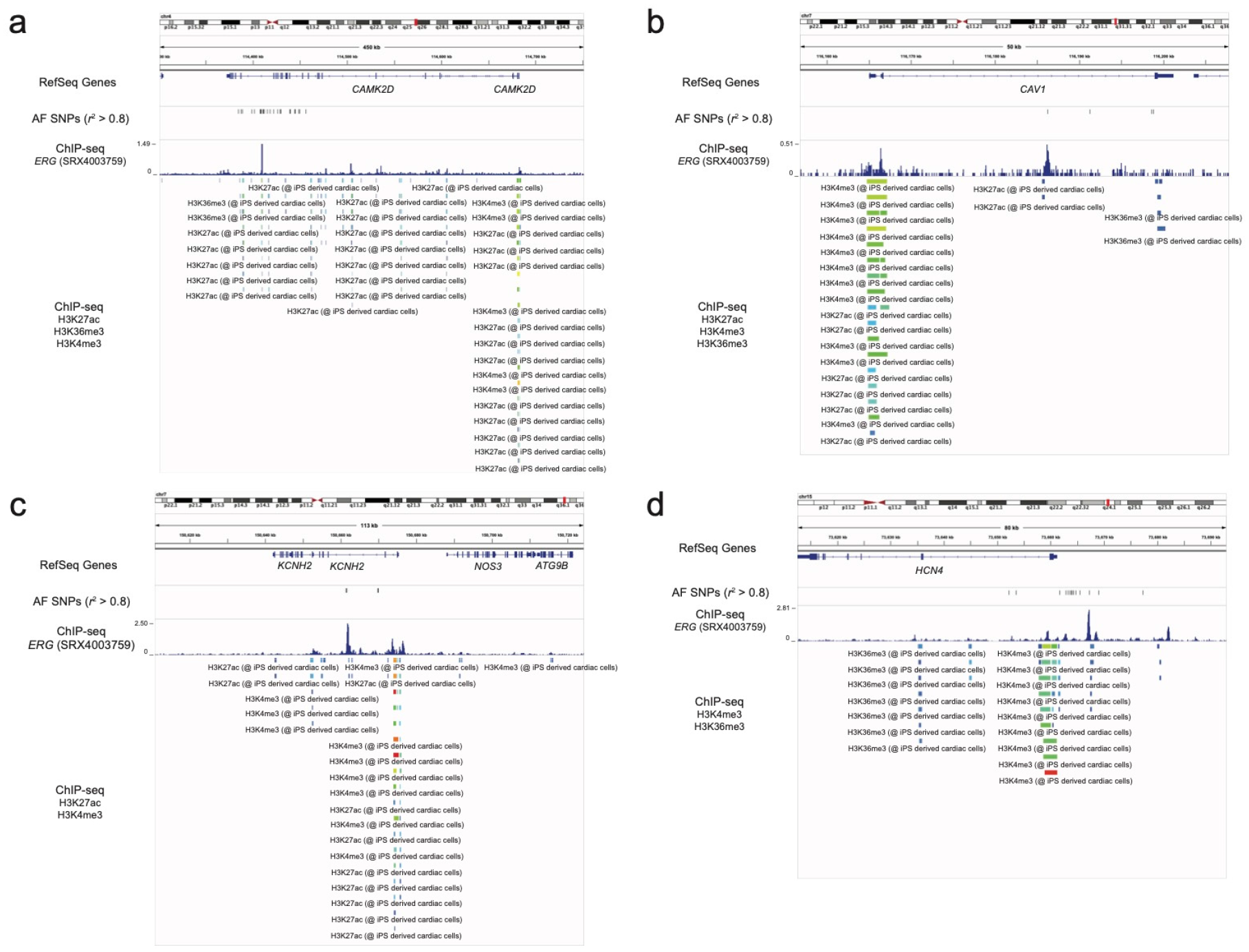
Ion channel-related gene and ERG. **a**, *CAMK2D* locus **b**, *CAV1* locus, **c**, *KCNH2* locus **d**, *HCN4* locus showing AF-associated variants (*r*^2^ > 0.8 in European samples in 1KG) and ChIP-seq track of ERG experiment (*P* = 1.0 × 10^−8^) and histone modification markers in iPSC-derived cardiac cells. iPSC, induced pluripotent stem cells.

**Supplementary Fig. 11.**
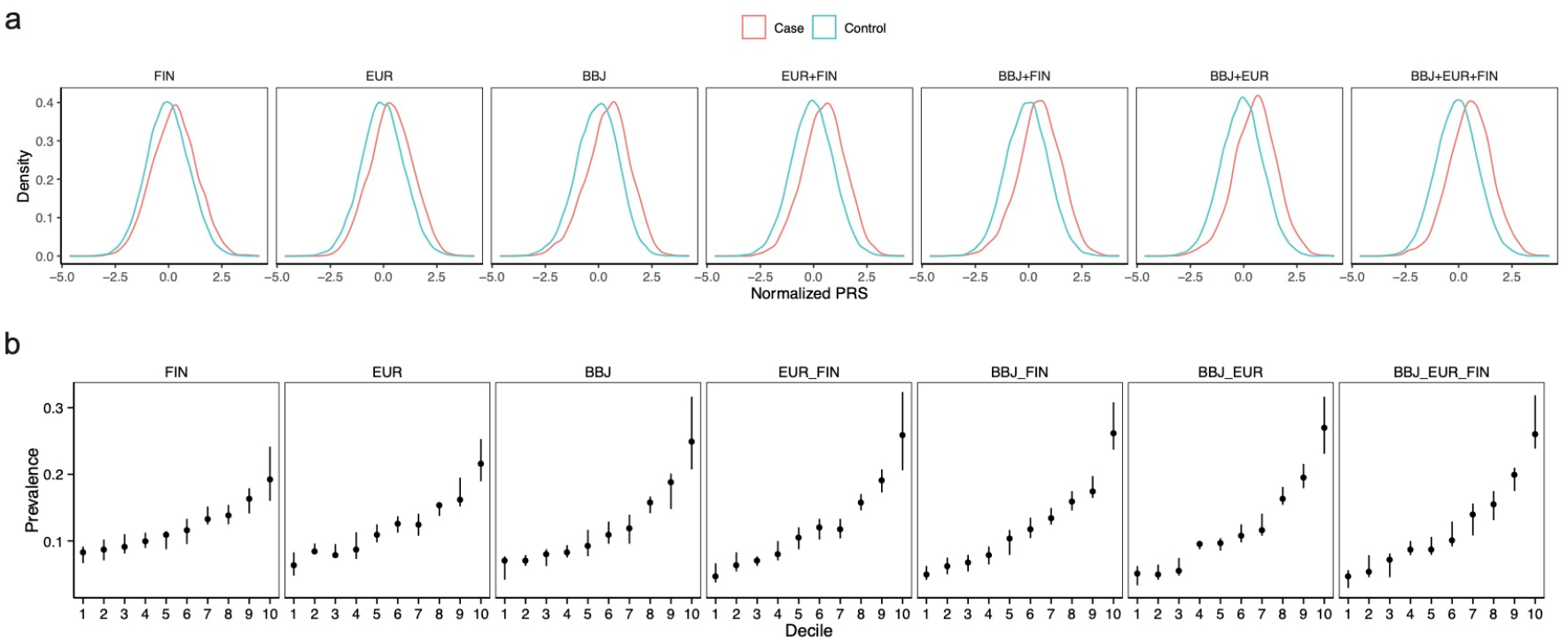
PRS distribution and AF prevalence. **a**, Distribution of PRS in the case and control samples for each combination of GWAS. **b** Prevalence of AF based on the AF-PRS deciles in each combination of GWAS. Data are presented as medians and 95% CI. PRS, polygenic risk score; AF, atrial fibrillation; GWAS, genome-wide association study; CI, confidence interval.

**Supplementary Fig. 12.**
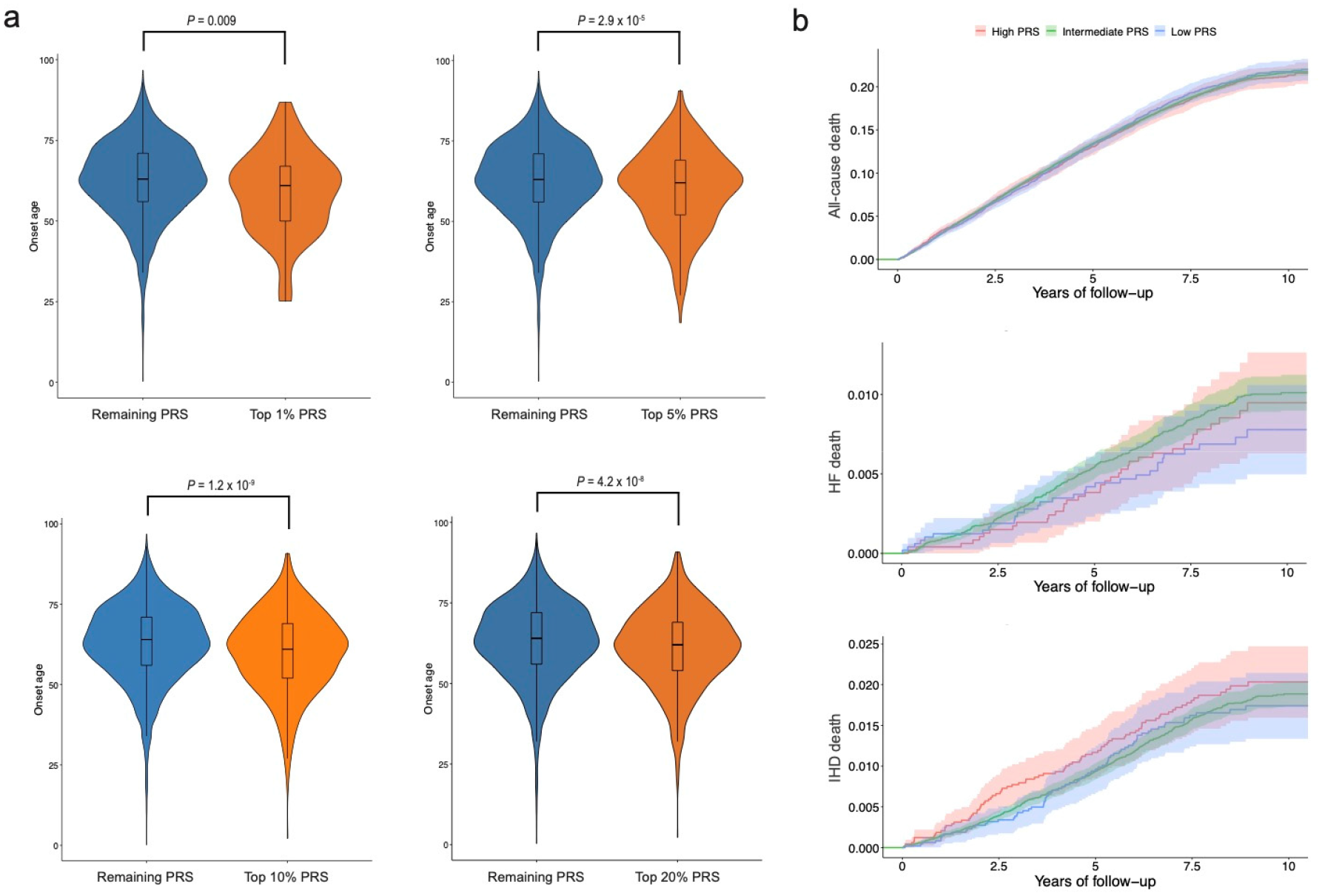
Impact of AF-PRS on the age of AF onset and long-term mortality. **a**, Comparison of the age of AF onset between samples with high PRS and those with remaining PRS. **b**, Kaplan-Meier estimates of cumulative events from all-cause death, heart failure death, and ischemic heart disease death with 95% CI. Individuals are classified into high PRS (top 10 percentile, red), low PRS (bottom 10 percentile, blue), and intermediate (others, green). AF, atrial fibrillation; PRS, polygenic risk score; CI, confidence interval.

**Supplementary Fig. 13.**
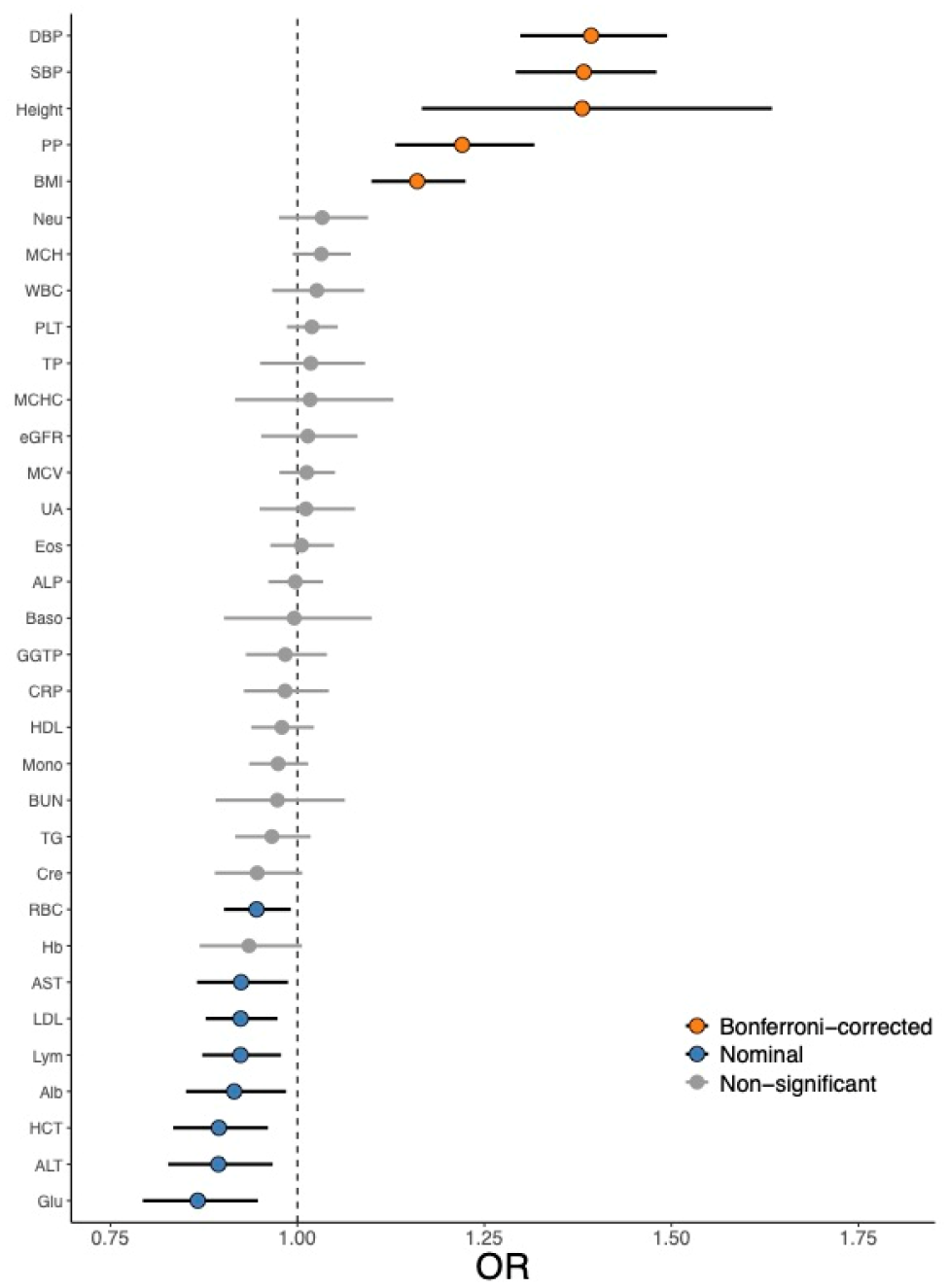
Causal effect of quantitative traits on AF development. Each point and error bar represents the causal estimates on the OR scale and the 95% CI for these ORs, respectively. We analyzed the MR analysis to estimate the causal effects using variants after excluding horizontal pleiotropic outliers. The *P* values were determined using the IVW two-sample MR method. OR, odds ratio; MR, Mendelian randomization; IVW, inverse-variance-weighting; DBP, diastolic blood pressure; SBP, systolic blood pressure; PP, pulse pressure; BMI, body mass index; Neu, neutrophil count; MCH, mean corpuscular hemoglobin; WBC, white blood cell count; PLT, platelet count; TP, total protein; MCHC, mean corpuscular hemoglobin concentration; eGFR, estimated glomerular filtration rate; MCV, mean corpuscular volume; UA uric acid; Eos, eosinophil count; ALP, alkaline phosphatase; Baso, basophil count; GGT, gamma-glutamyl transferase; CRP, C-reactive protein; HDL, high-density lipoprotein cholesterol; Mono, monocyte count; BUN, blood urea nitrogen; TG, triglyceride; Cre, serum creatinine; RBC, red blood cell; Hb, hemoglobin; AST, aspartate aminotransferase; LDL, low-density lipoprotein cholesterol; Lym, lymphocyte count; Alb, albumin; HCT, hematocrit; ALT, alanine aminotransferase; Glu, blood sugar

**Supplementary Fig. 14.**
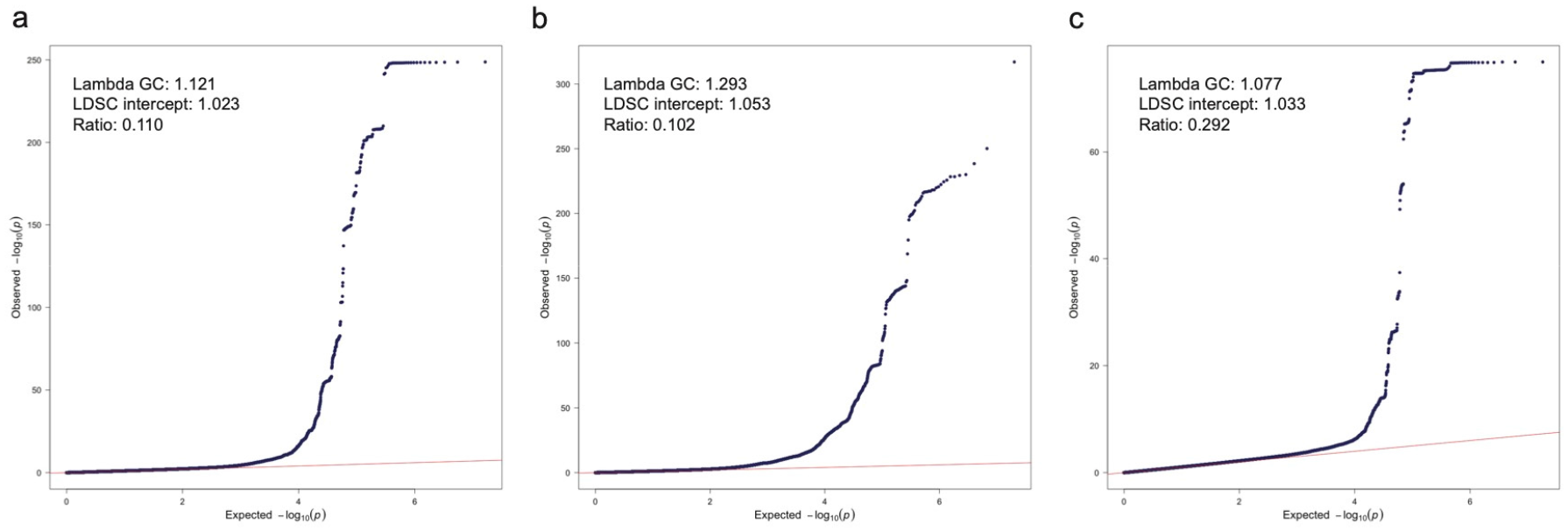
Quantile-quantile plot for GWAS of three studies. **a**, the Japanese GWAS. **b**, GWAS of a meta-analysis in European population (EUR). **c**, GWAS in data from FinnGen project (FIN). GWAS, genome-wide association study.

**Supplementary Fig. 15.**
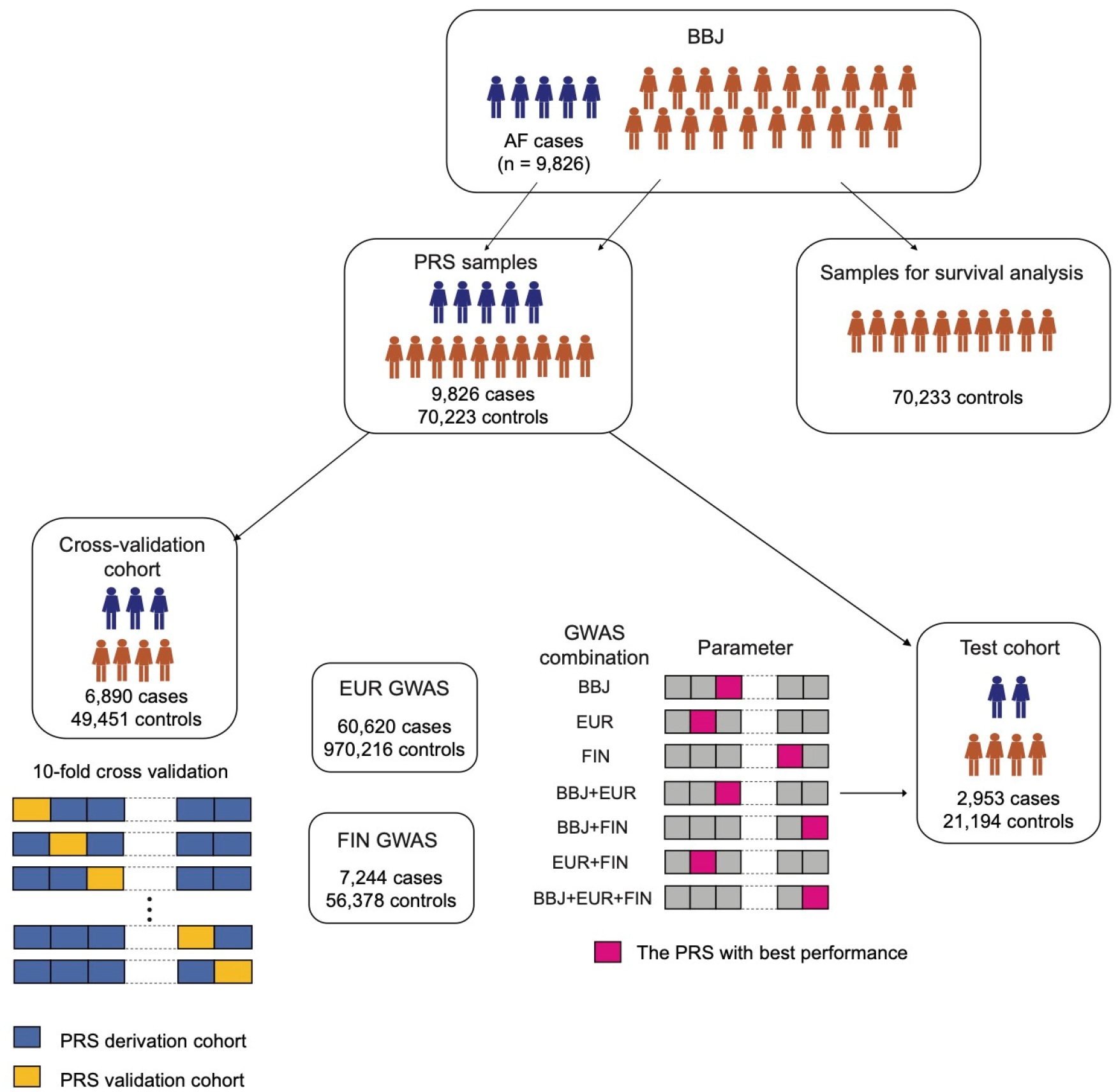
Analytical scheme for PRS derivation, validation, and test cohorts and survival analysis. Schematic representation for cross-validation, performance testing of the AF-PRS in the independent test cohort, and survival analysis for AF-PRS. PRS, polygenic risk score; AF, atrial fibrillation.

## Supplementary Dataset

### 1. Novel 5 loci in the Japanese GWAS

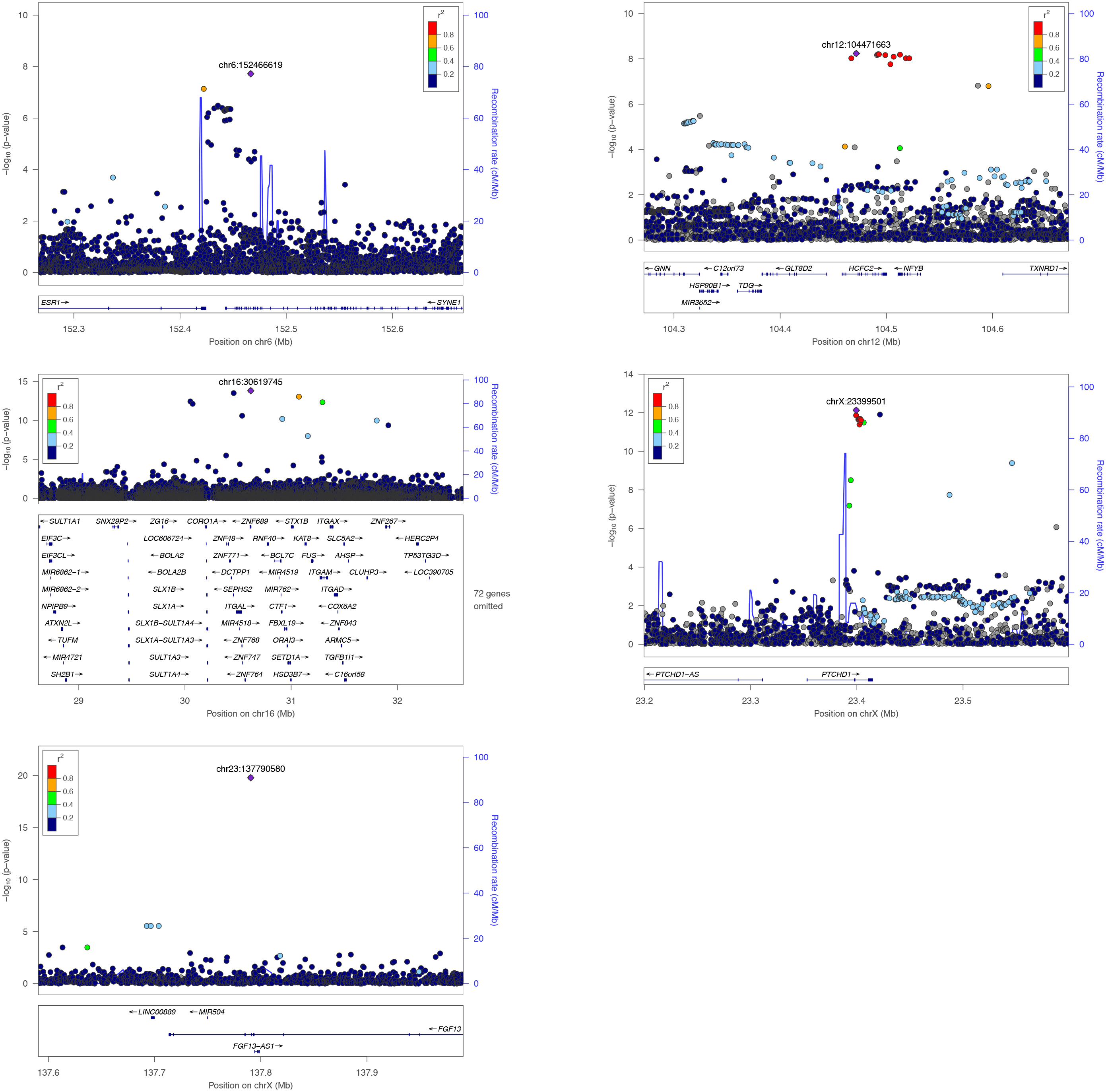

### 2. Previously reported 26 loci in the Japanese GWAS

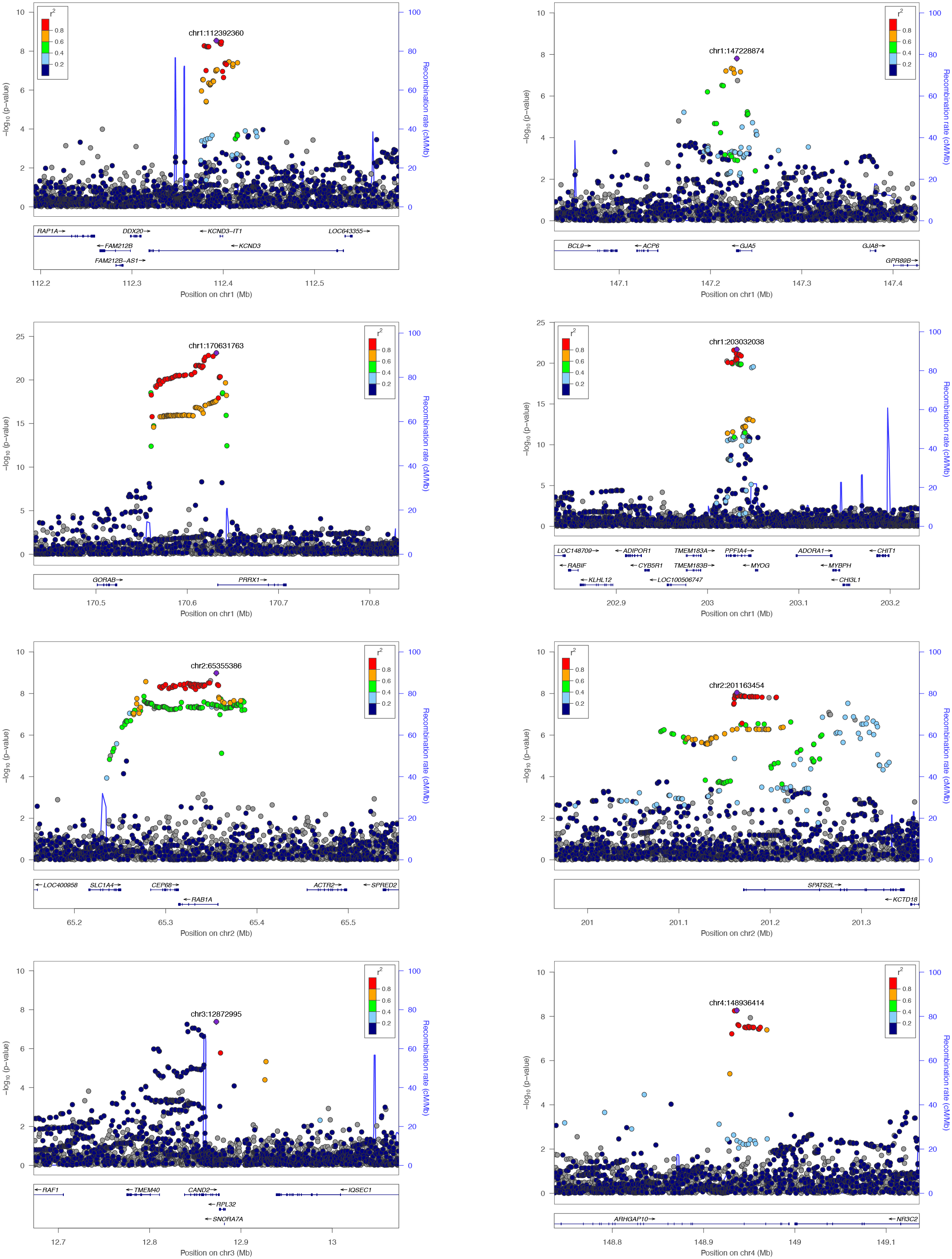
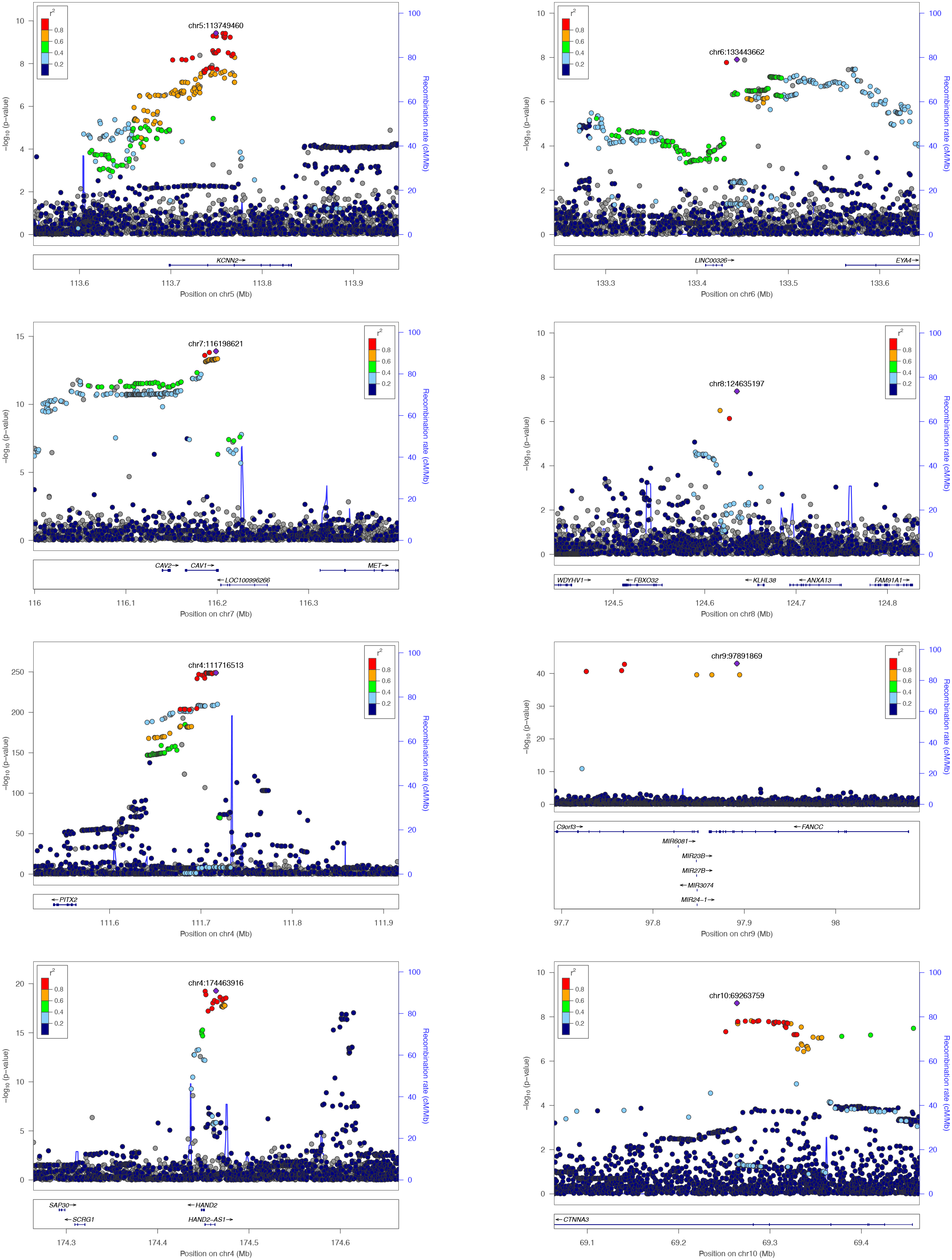
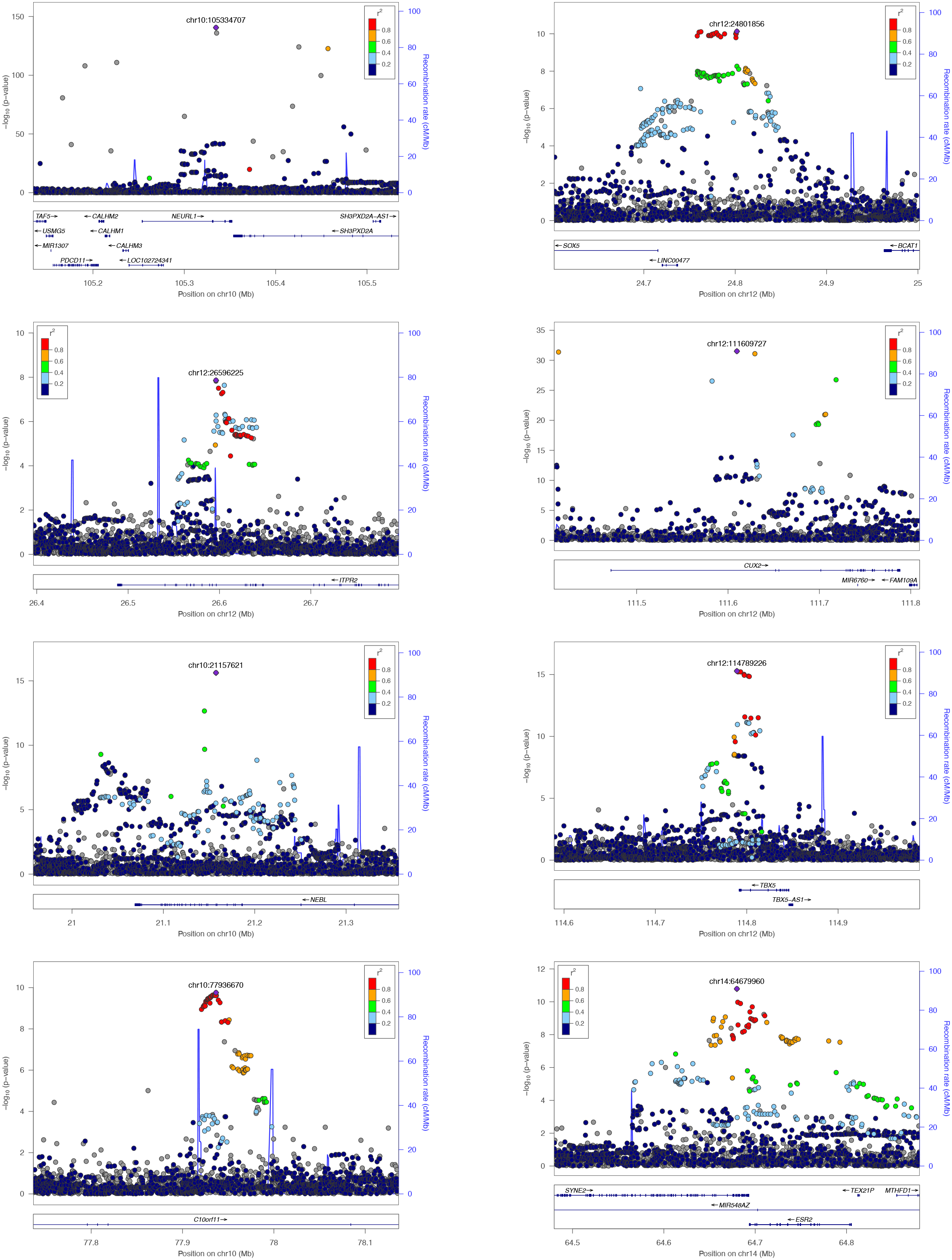
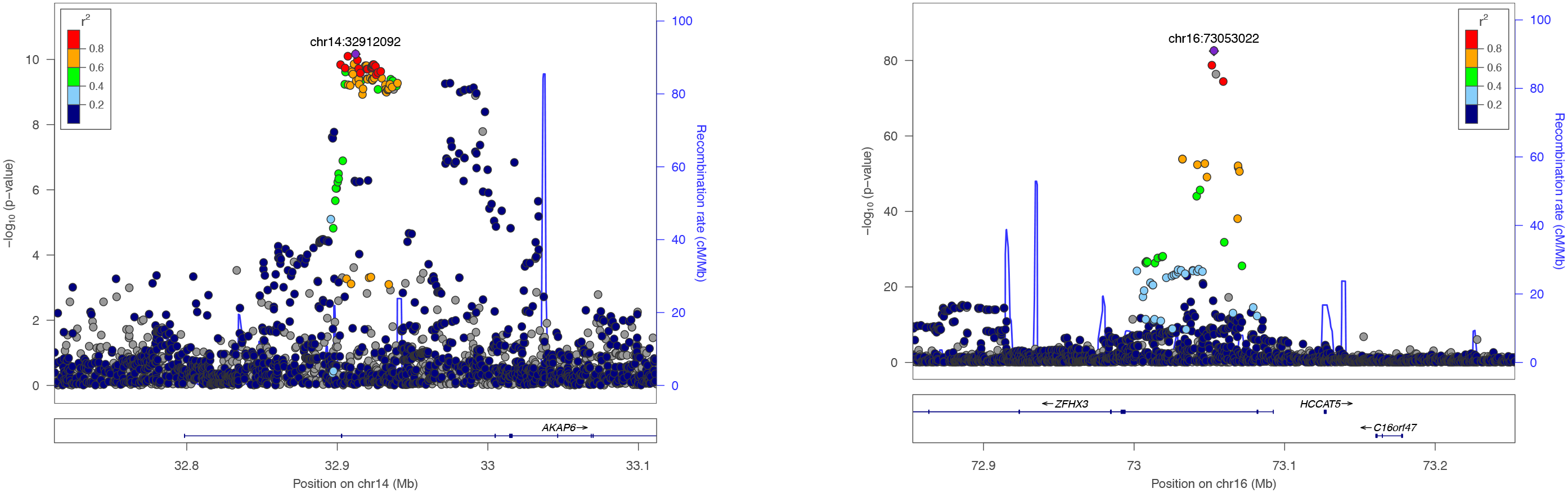

### 3. Novel 33 loci in the trans-ancestry meta-GWAS

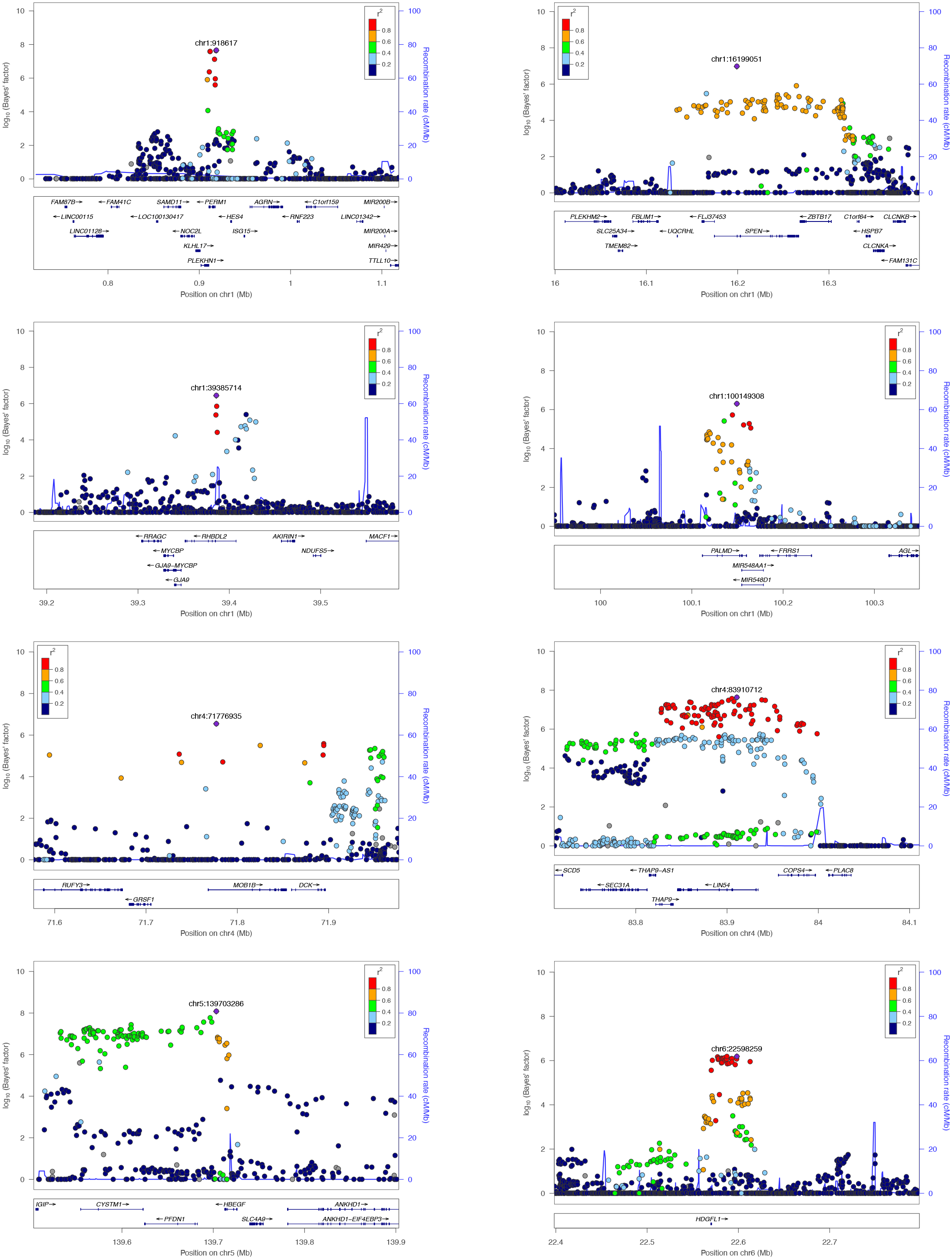
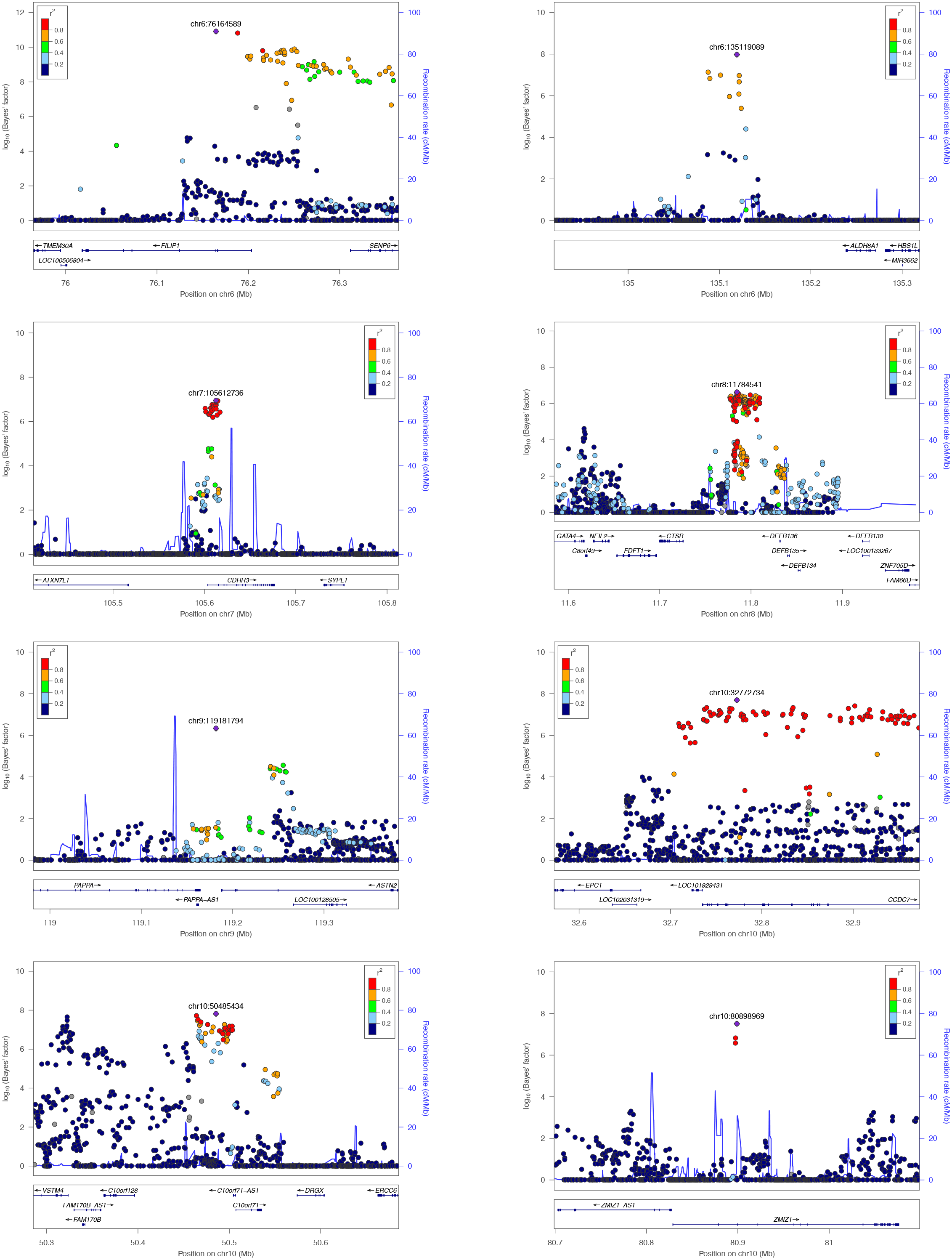
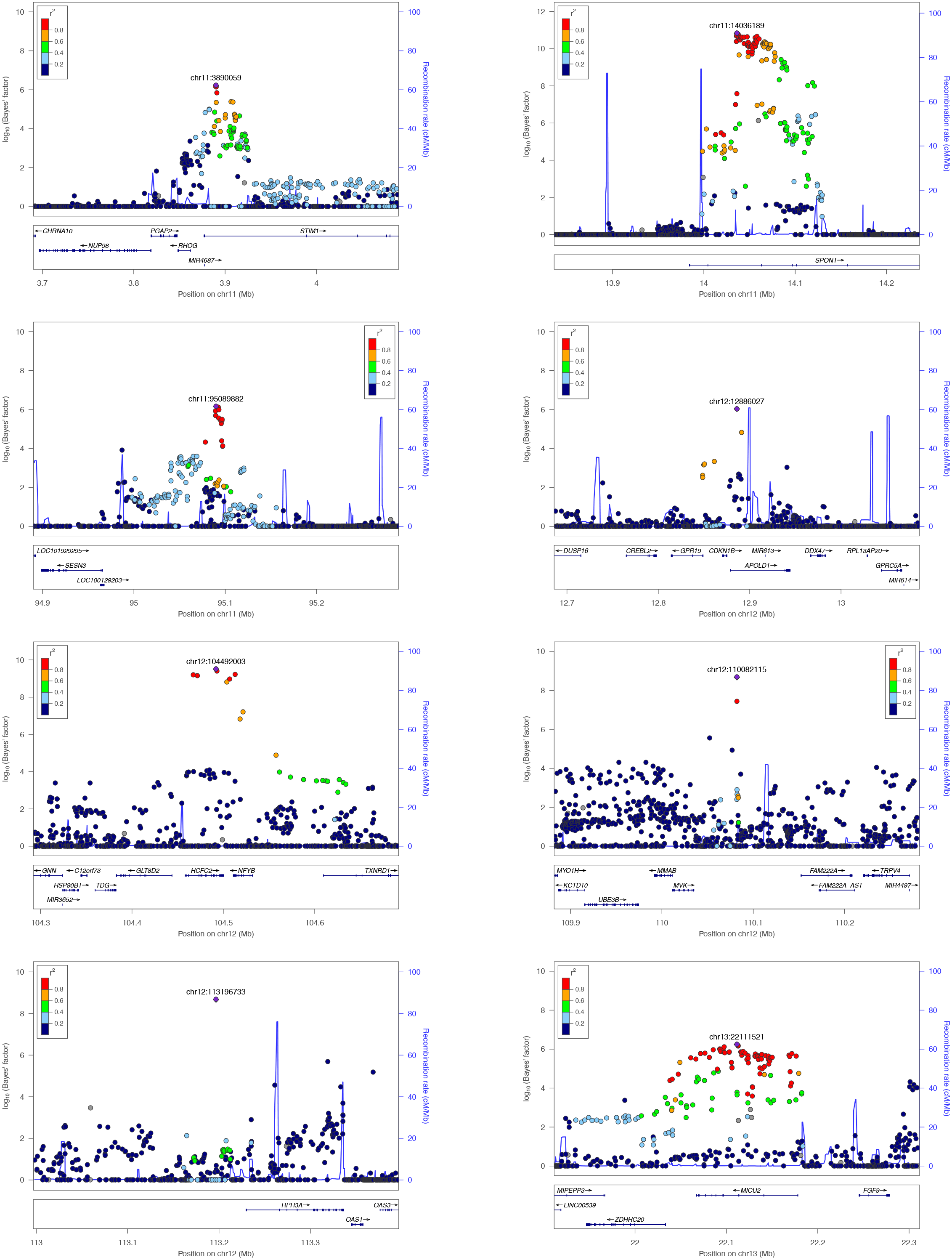
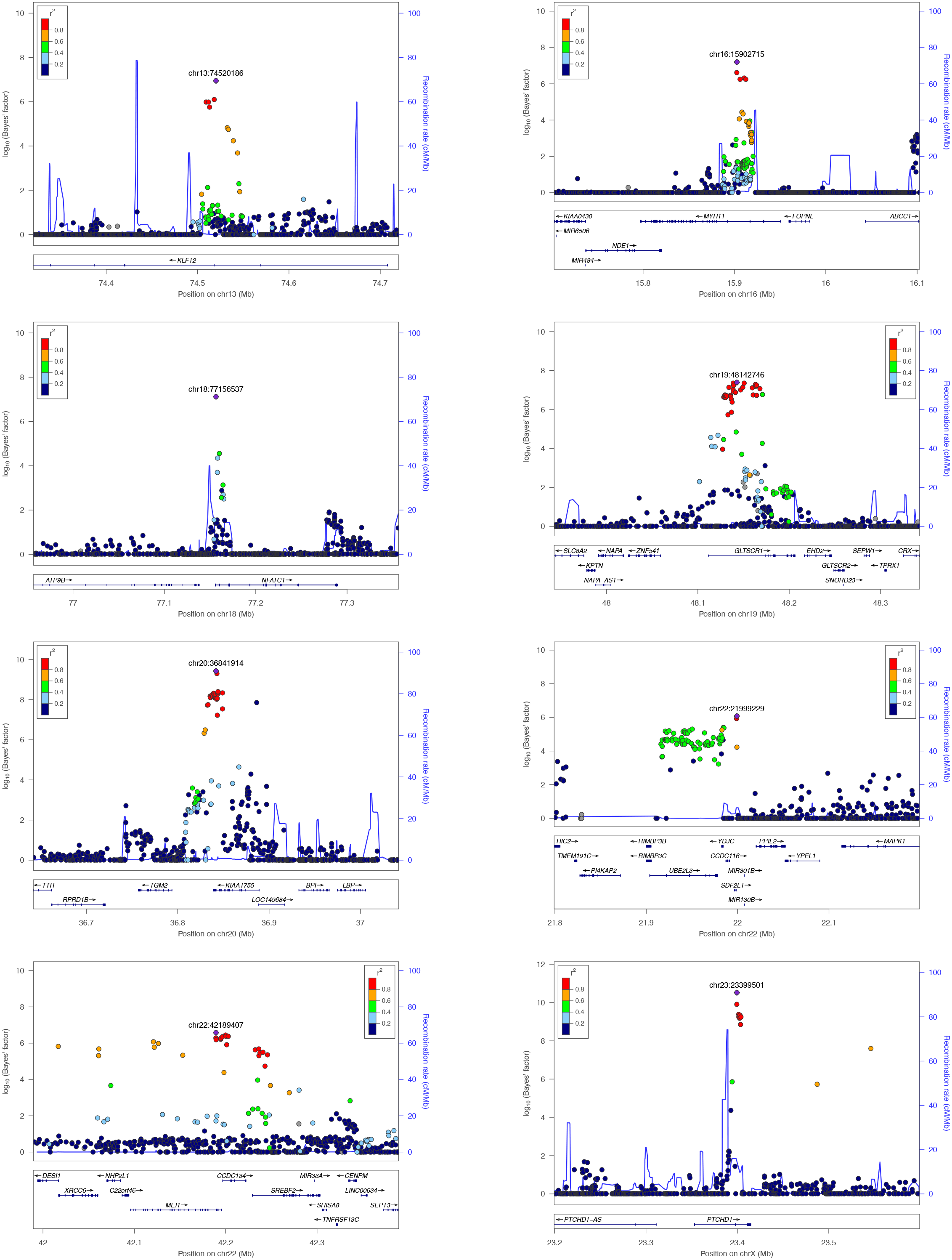
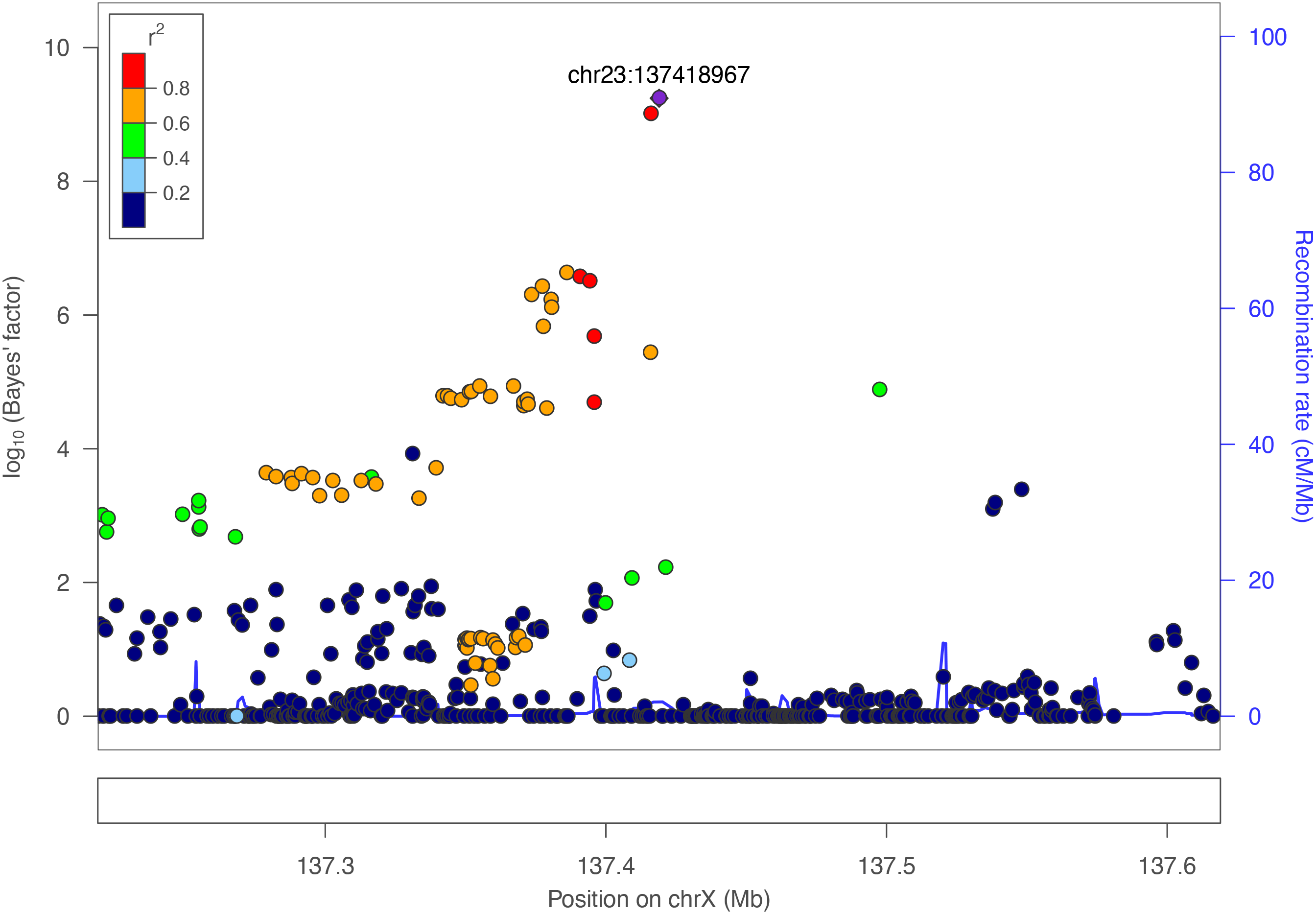

### 4. Previously reported 127 loci in the trans-ancestry meta-GWAS

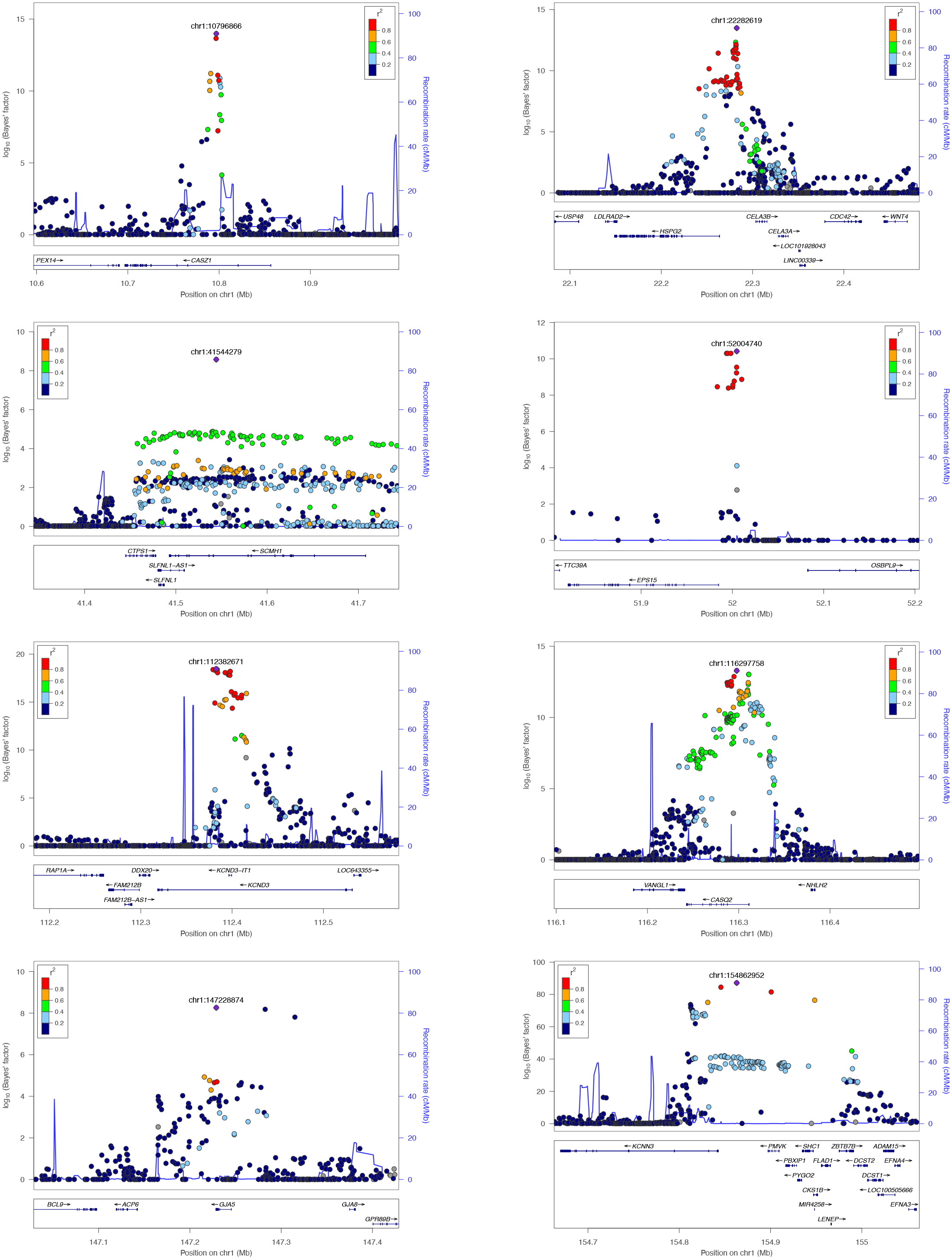
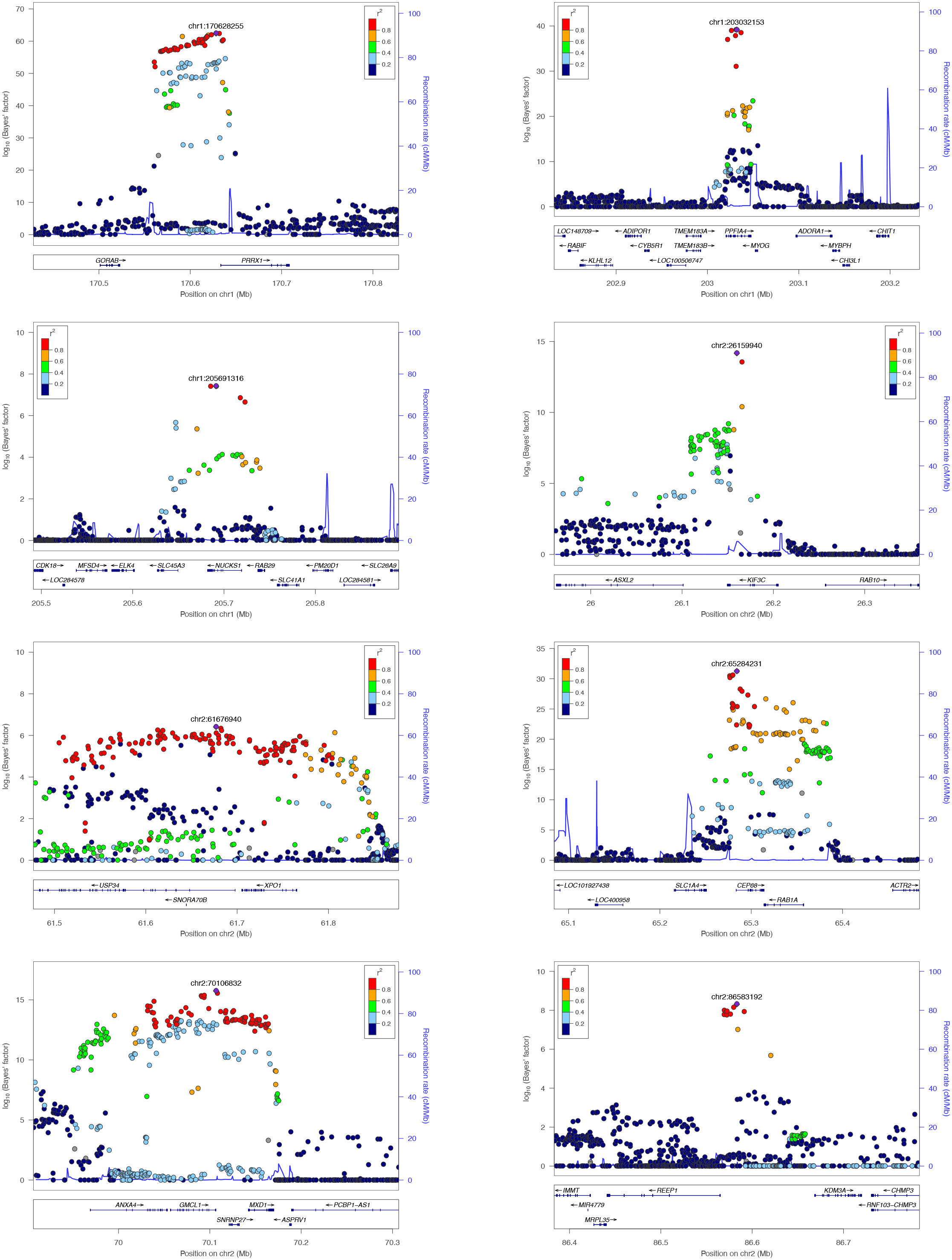
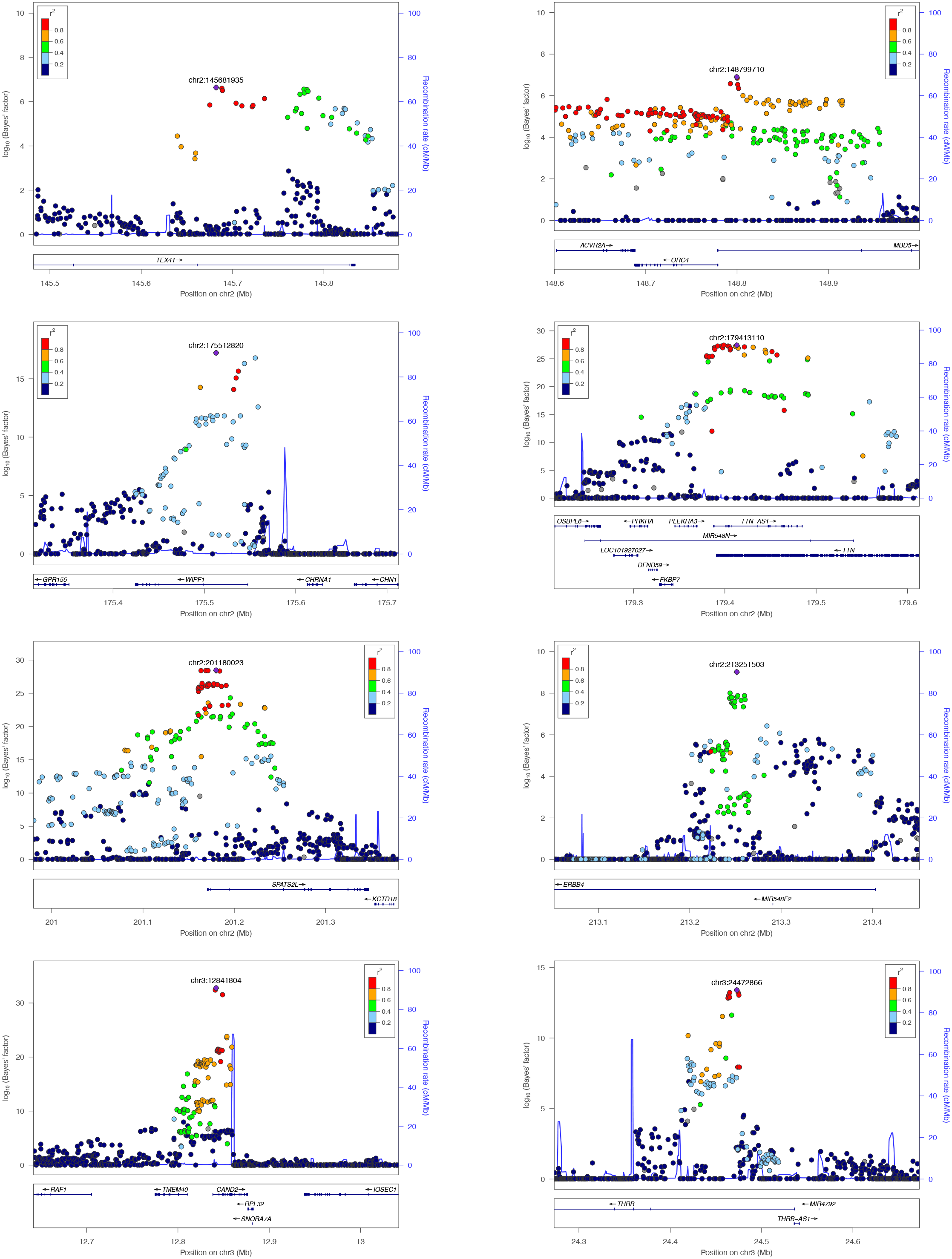
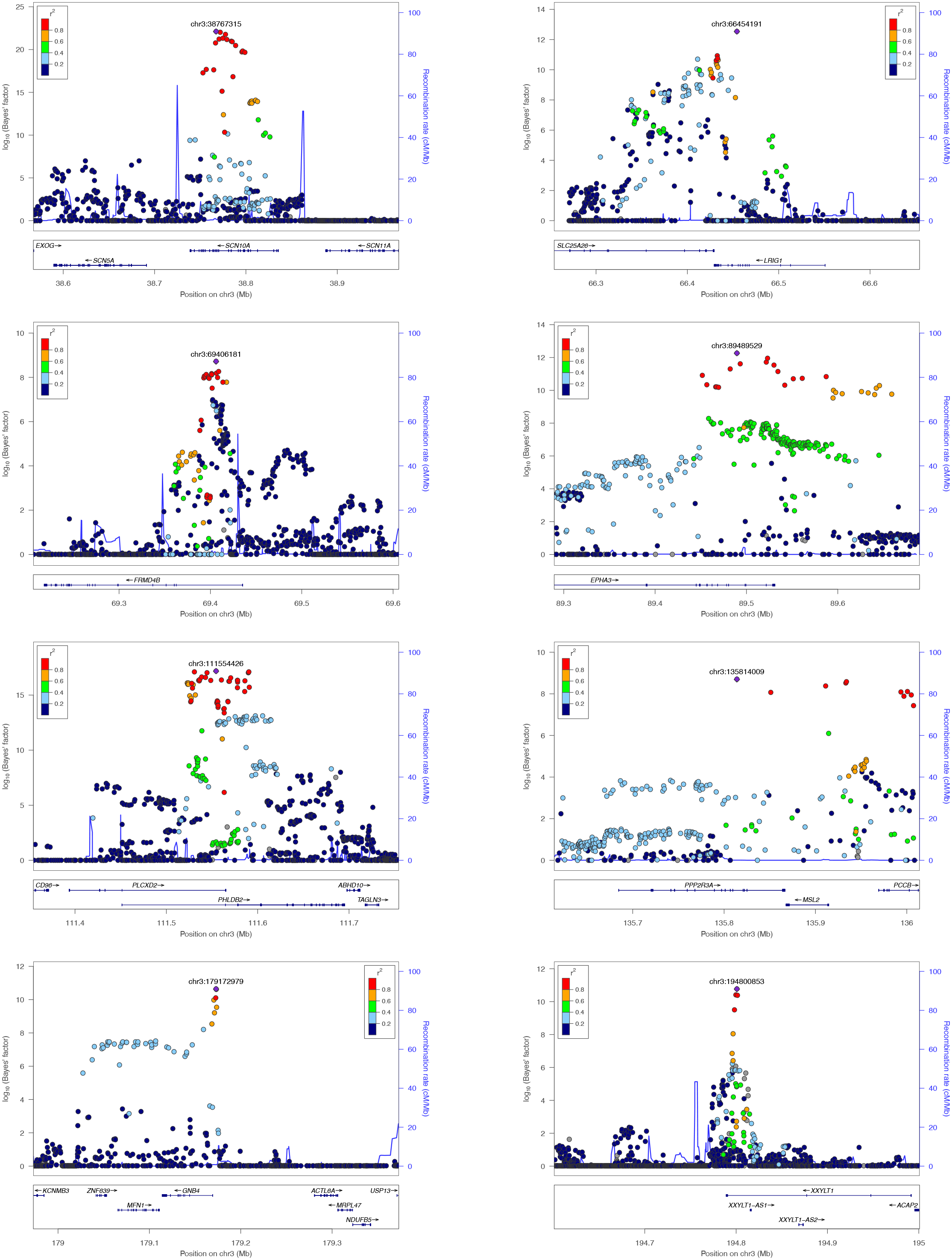
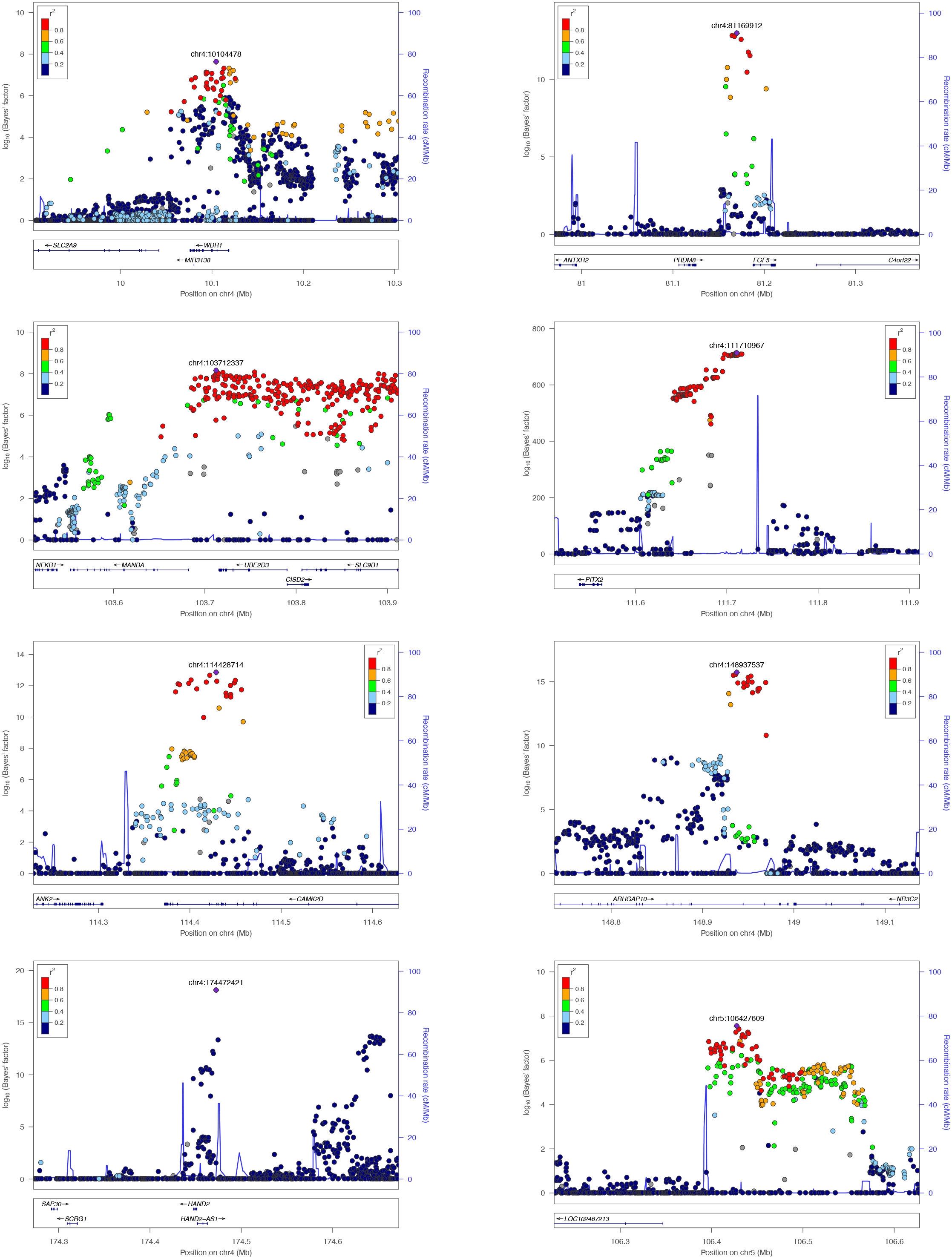
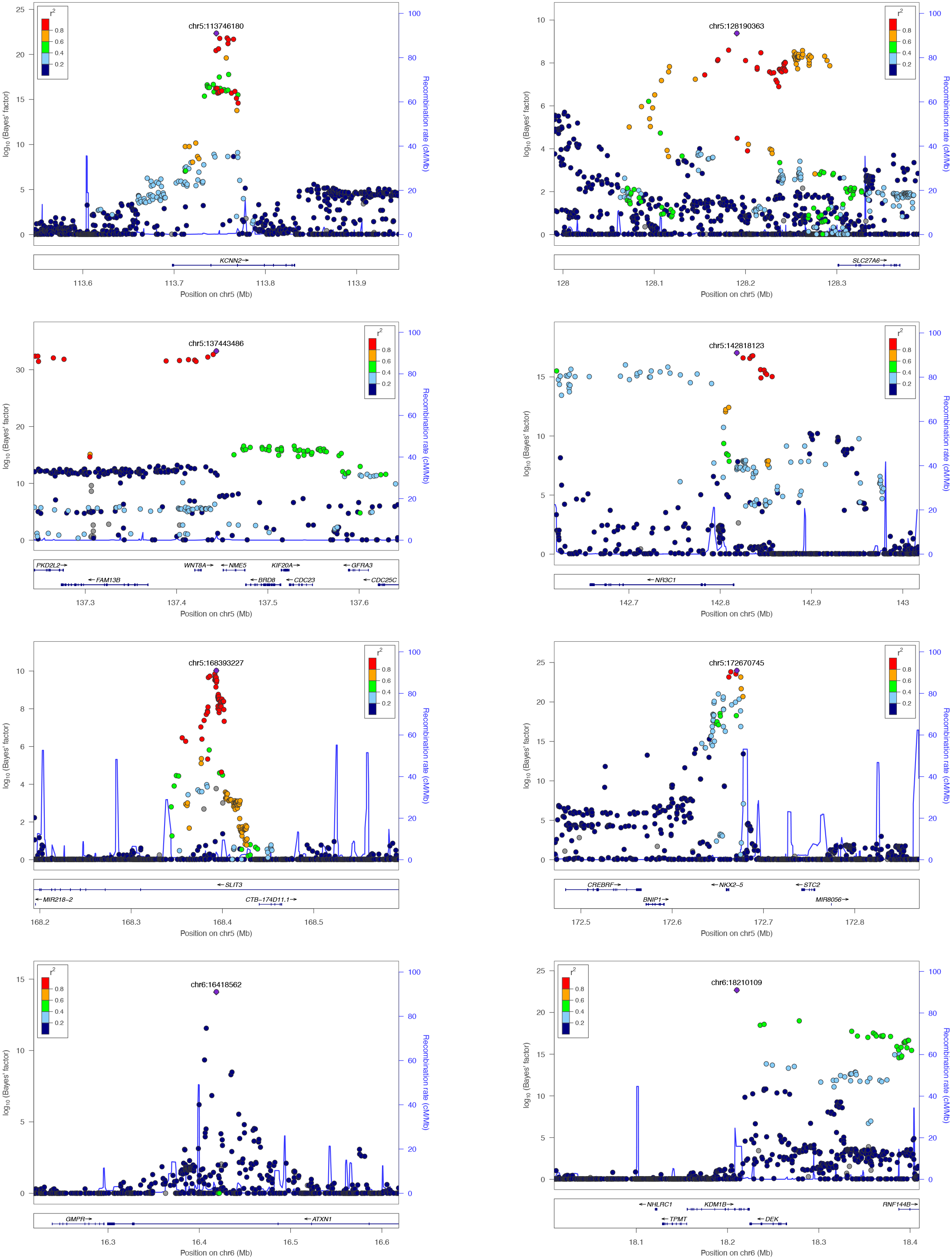
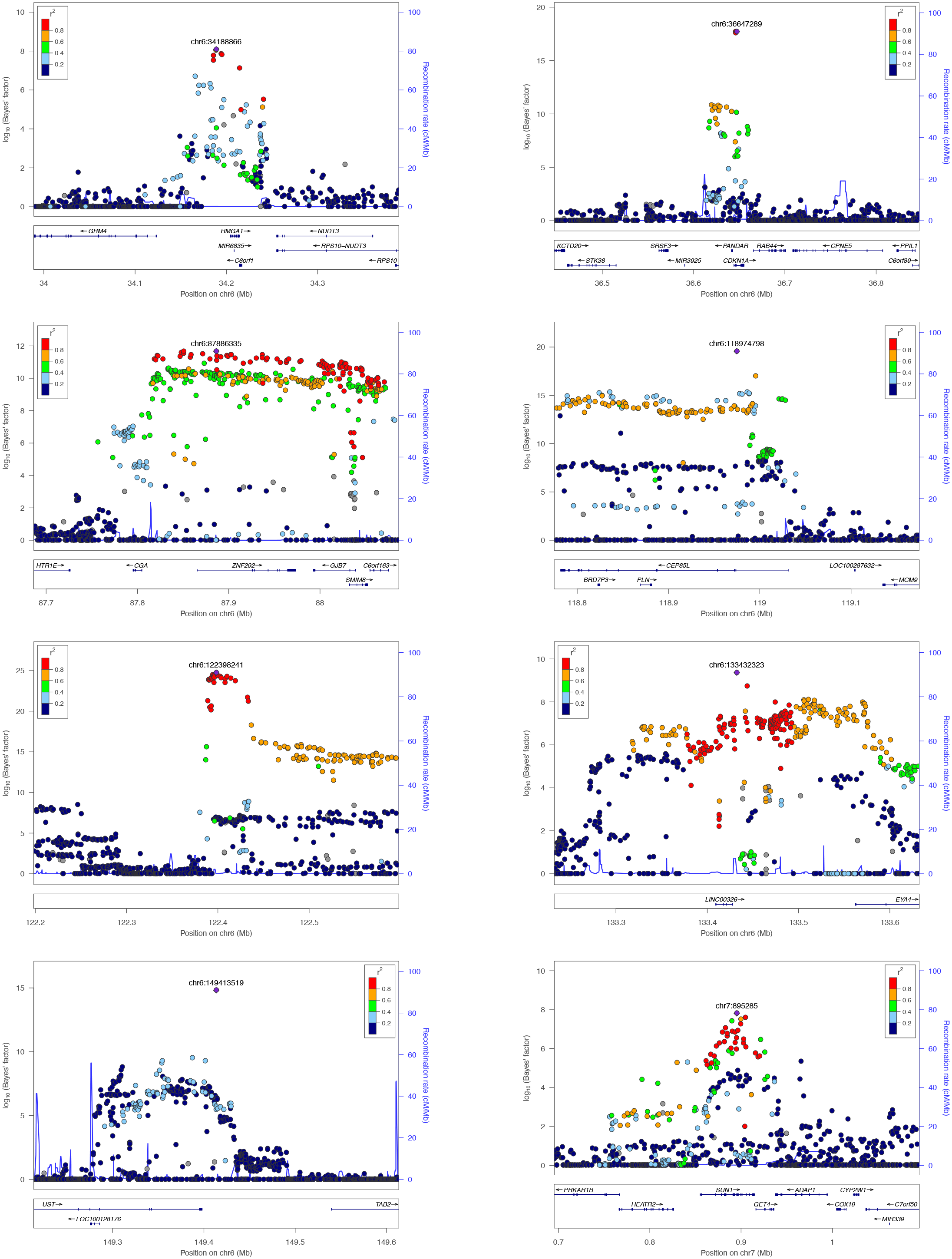
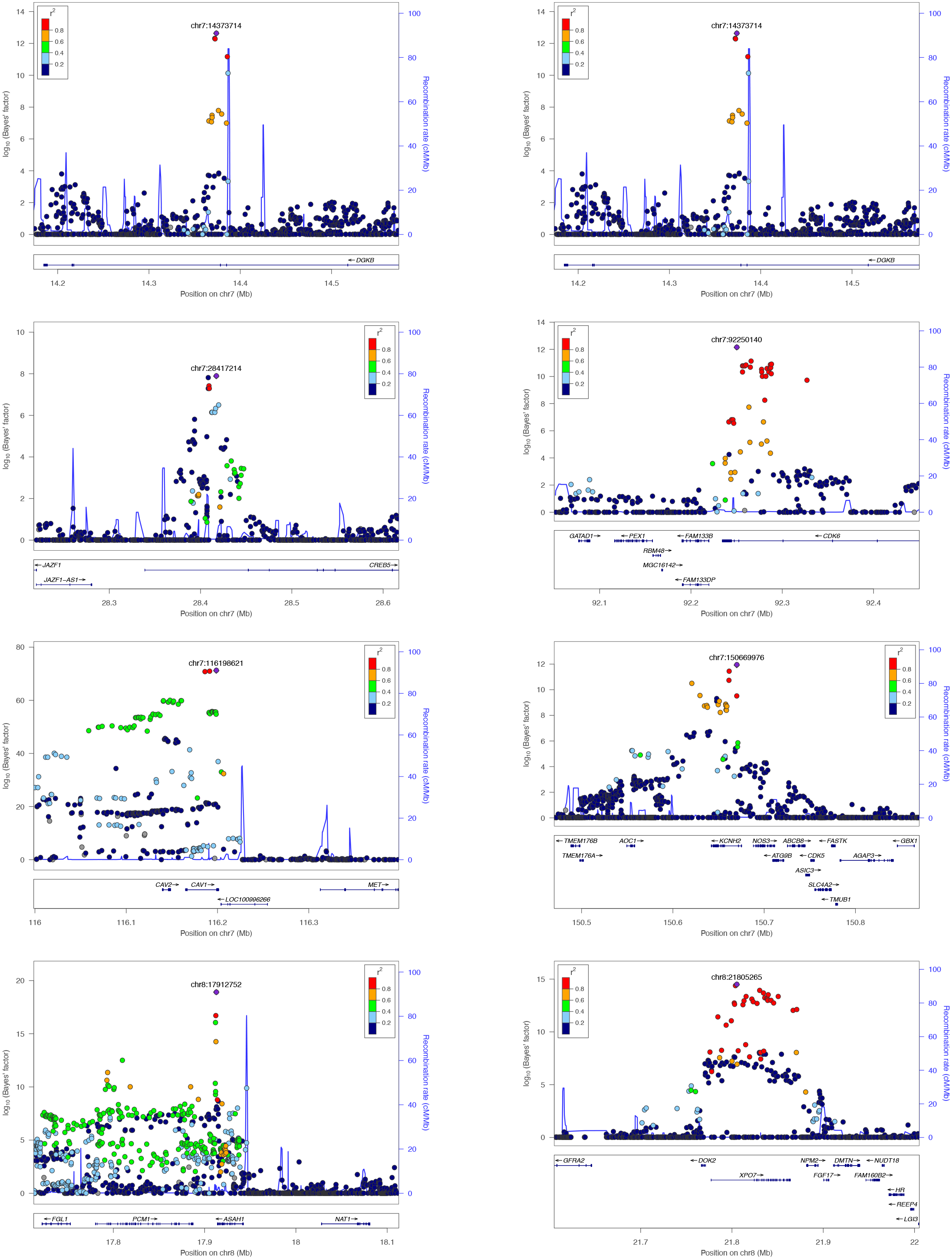
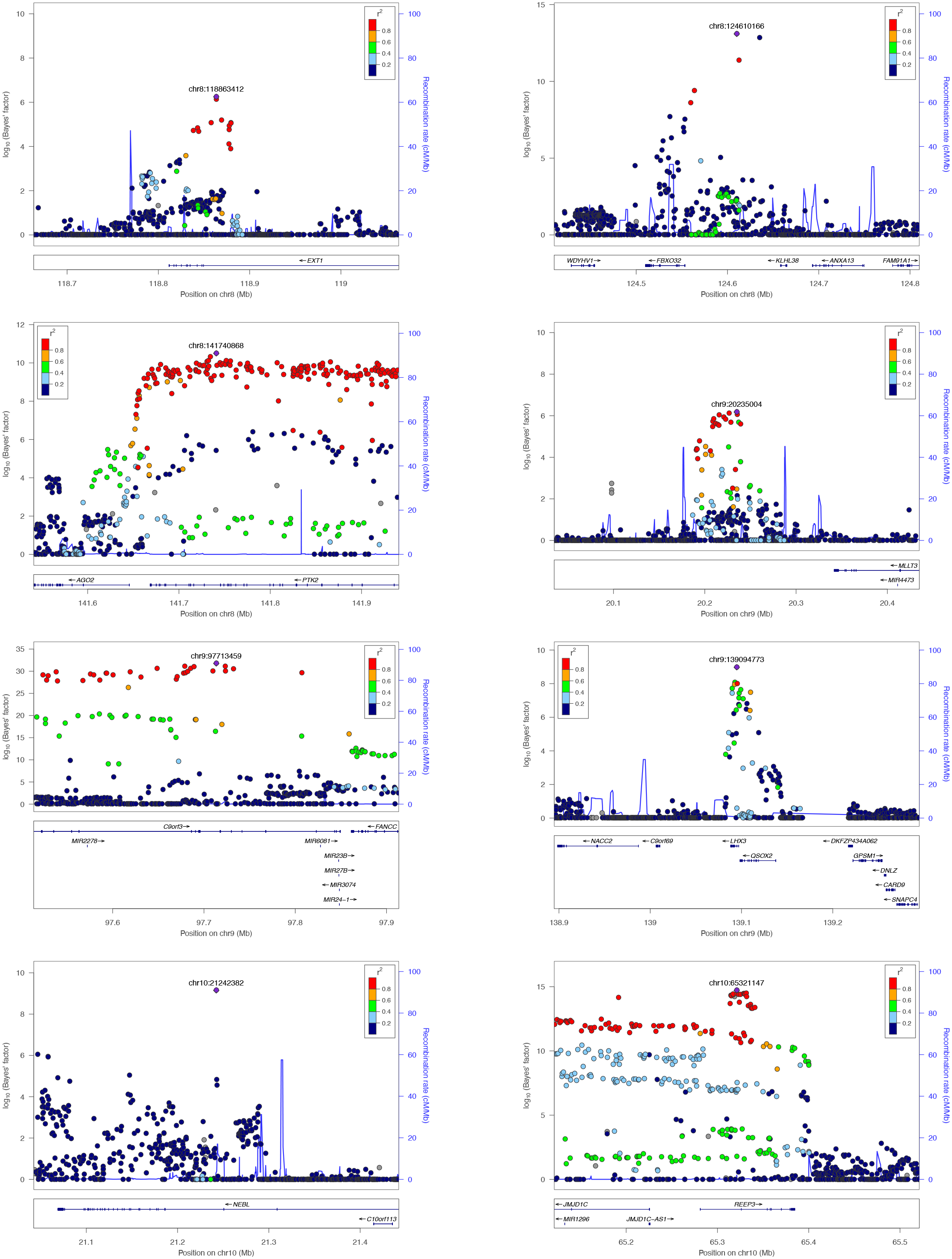
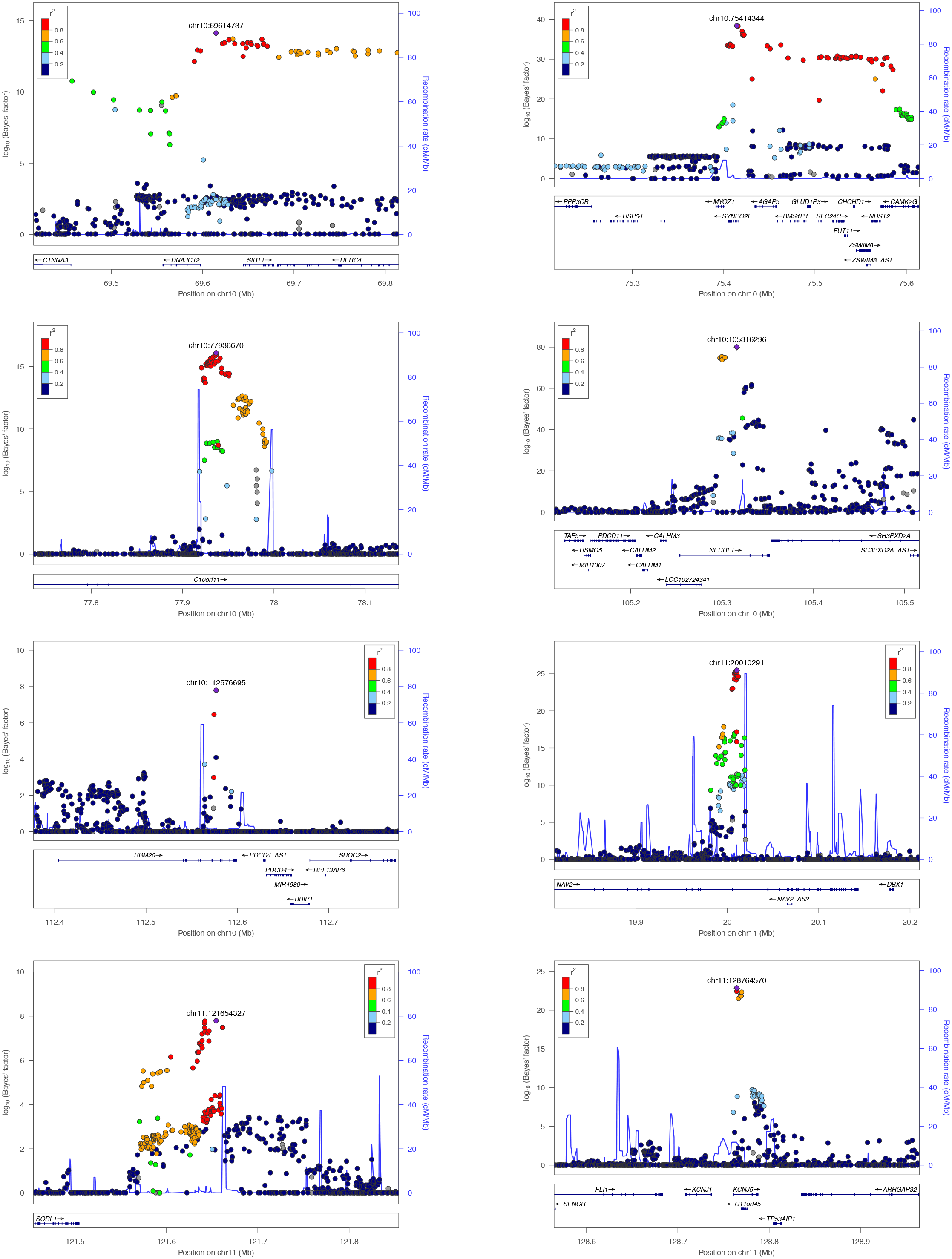
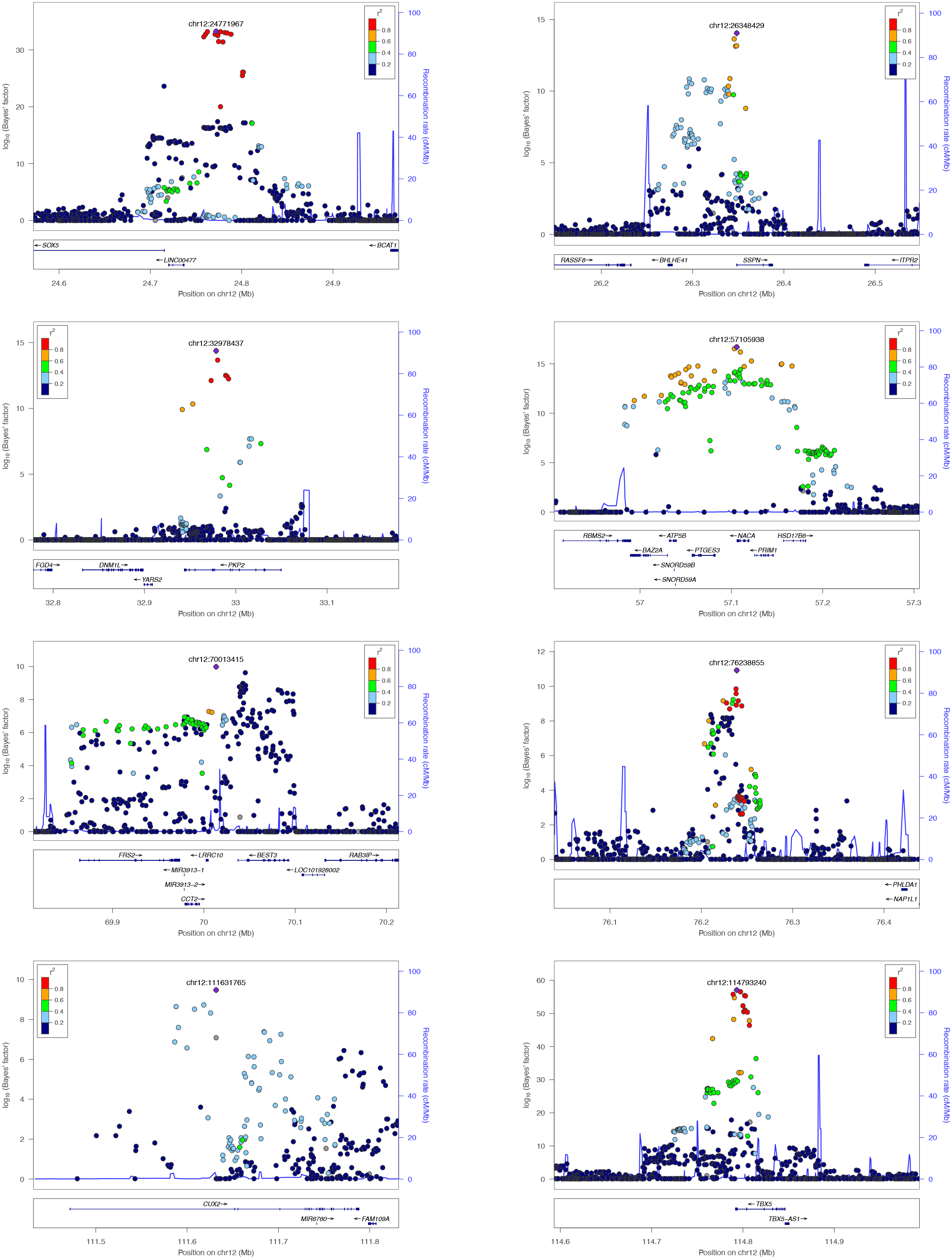
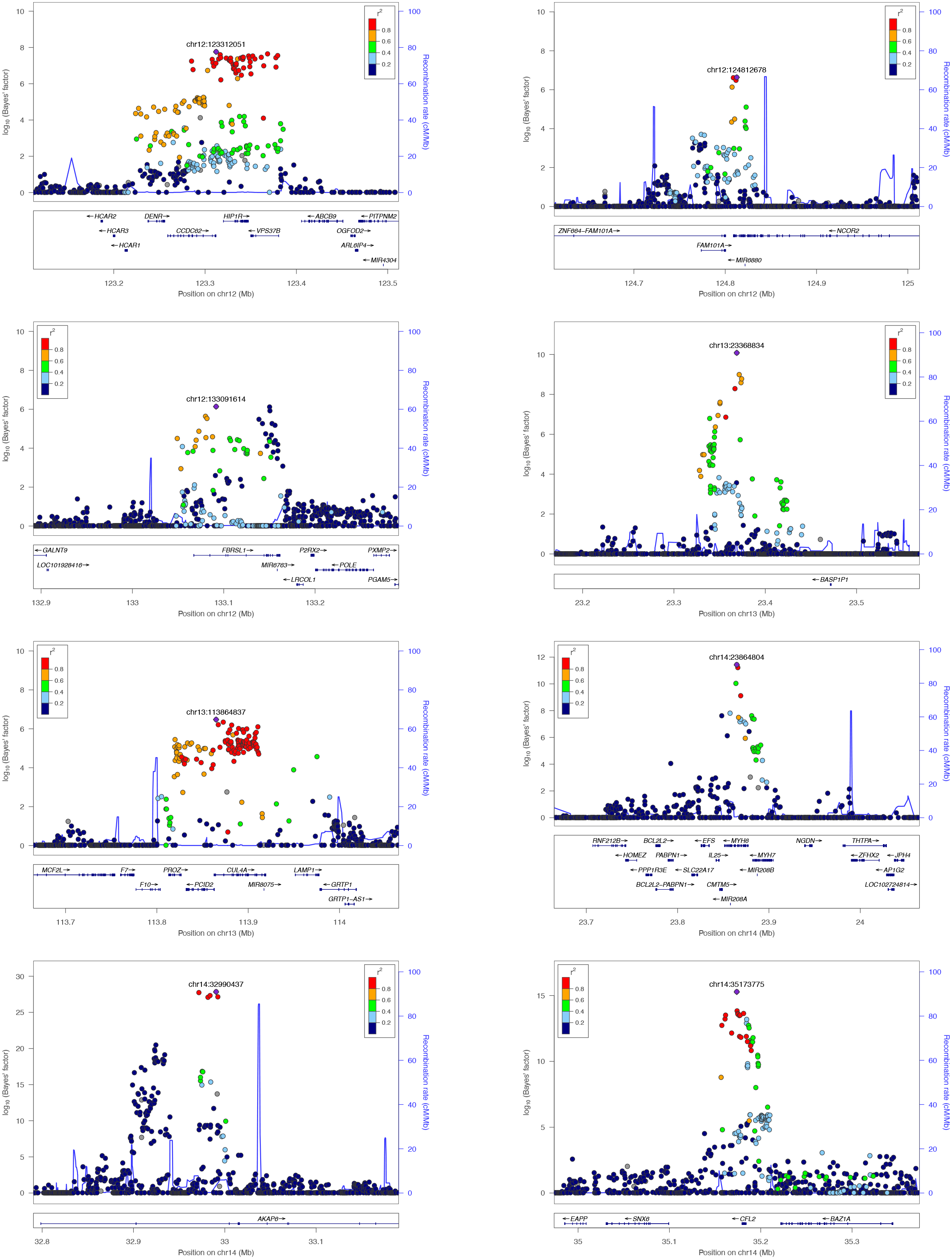
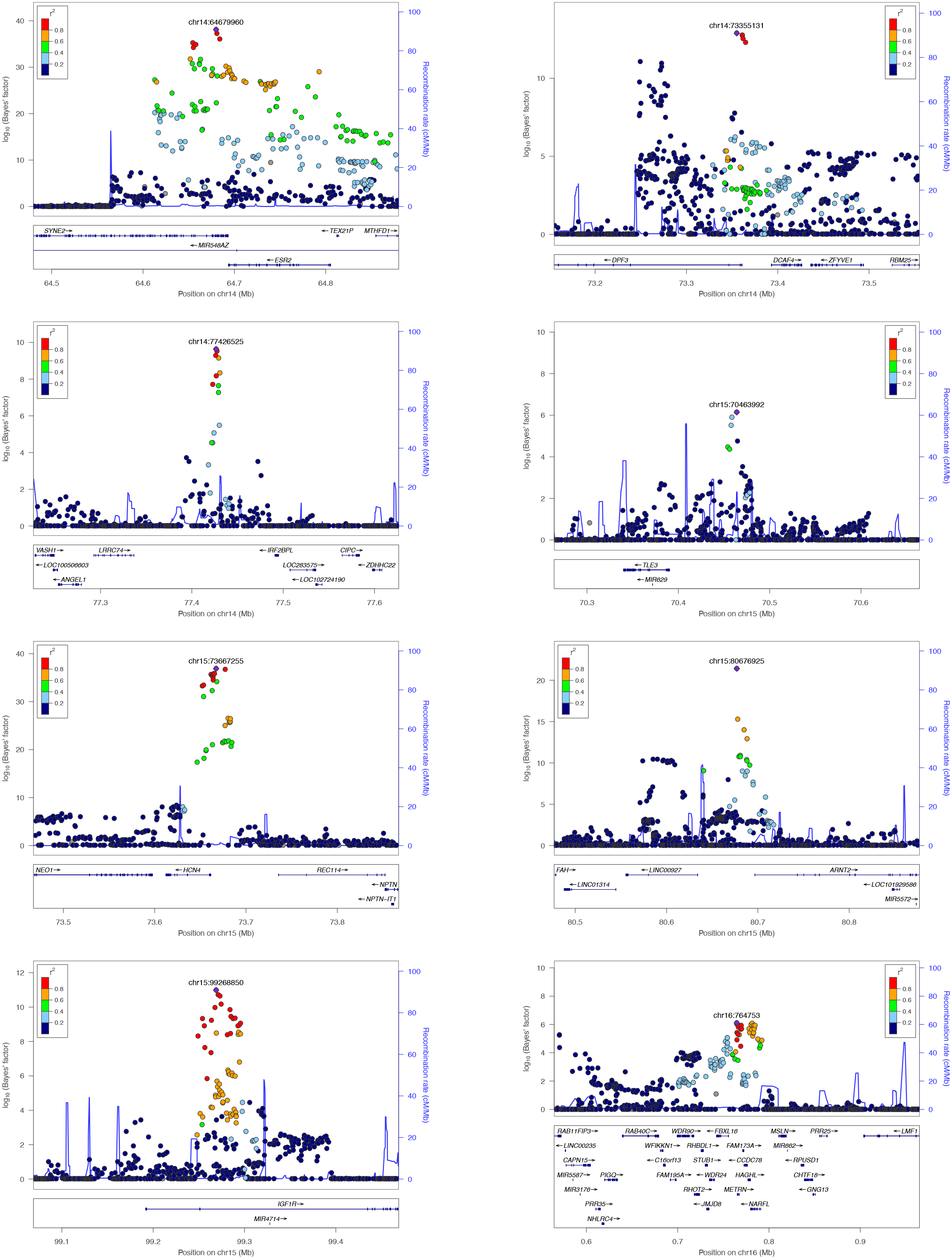
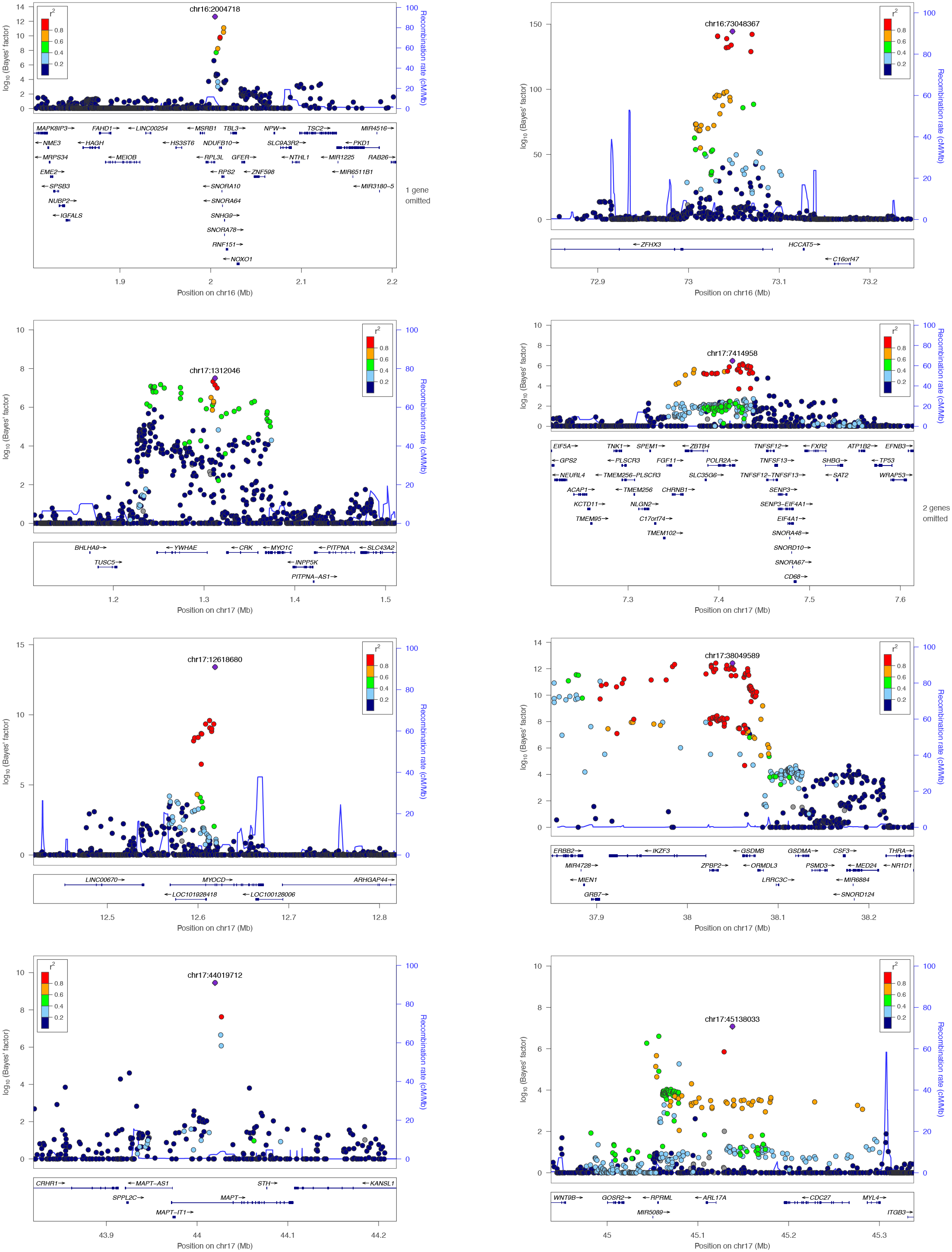
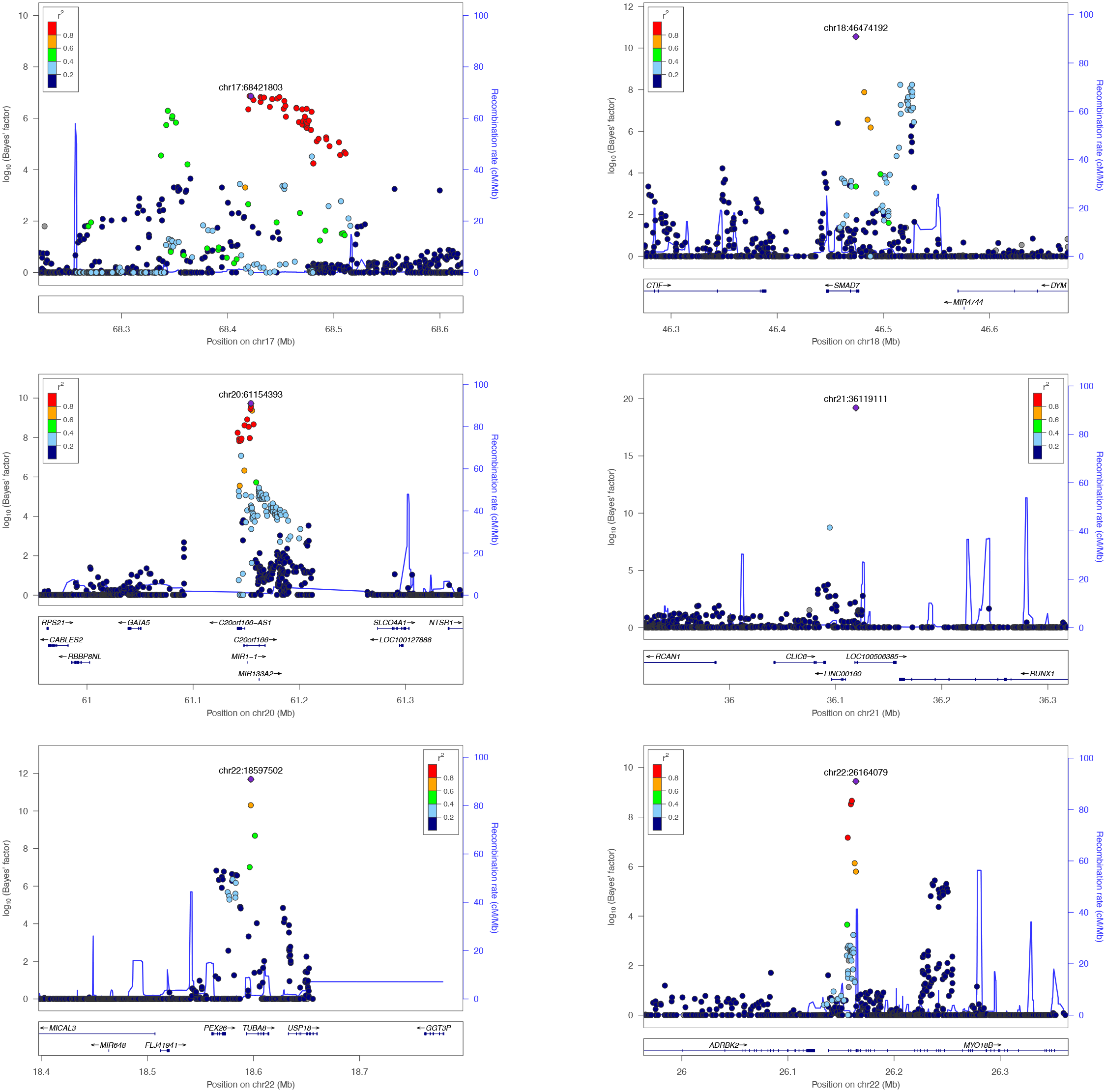

